# Early oral switch in low-risk *Staphylococcus aureus* bloodstream infection

**DOI:** 10.1101/2023.07.03.23291932

**Authors:** Achim J. Kaasch, Luis Eduard López-Cortés, Jesús Rodríguez-Baño, José Miguel Cisneros, M. Dolores Navarro, Gerd Fätkenheuer, Norma Jung, Siegbert Rieg, Raphaël Lepeule, Laetitia Coutte, Louis Bernard, Adrien Lemaignen, Katrin Kösters, Colin R. MacKenzie, Alex Soriano, Stefan Hagel, Bruno Fantin, Matthieu Lafaurie, Jean-Philippe Talarmin, Aurélien Dinh, Thomas Guimard, David Boutoille, Tobias Welte, Stefan Reuter, Jan Kluytmans, Maria Luisa Martin, Emmanuel Forestier, Hartmut Stocker, Virginie Vitrat, Pierre Tattevin, Anna Rommerskirchen, Marion Noret, Anne Adams, Winfried V. Kern, Martin Hellmich, Harald Seifert, SABATO study group (members and affiliations listed in Acknowledgement record)

## Abstract

**Background:** *Staphylococcus aureus* bloodstream infection (SAB) is treated with at least 14 days of intravenously administered antimicrobials. We assessed the efficacy and safety of an early oral switch therapy in patients at low risk for SAB-related complications.

**Methods:** In an international non-inferiority trial, we randomized patients with SAB after 5 to 7 days of intravenous antimicrobial therapy to either switch to an oral antimicrobial or to continue with intravenous standard therapy. Main exclusion criteria were signs and symptoms of complicated SAB, non-removable foreign devices, and severe comorbidity. Composite primary endpoint was the occurrence of any SAB-related complication (relapsing SAB, deep-seated infection, and mortality attributable to SAB) within 90 days.

**Results:** 213 patients were randomized into the intention-to-treat population. In the oral switch group, 14/108 (13%) participants reached the primary endpoint versus 13/105 (12%) in the standard therapy group (adjusted difference 0.7%, 95% confidence interval [CI] -7.8% to 9.1%). Participants in the oral switch group were discharged earlier (median hospital stay from SAB onset of 12 days versus 16 days; adjusted difference -3.1 days [95% CI -7.5 to 1.4]). There was no statistical difference in 30-day survival and complications of intravenous administration. More participants in the oral group experienced at least one serious adverse event (34% versus 26%, p=0.292).

**Conclusion:** Oral switch was non-inferior to intravenous standard therapy in participants with low-risk SAB. However, a careful assessment of patients for signs and symptoms of complicated SAB at time of presentation and thereafter is necessary before considering early oral switch therapy.

The trial was registered as NCT01792804 in ClinicalTrials.gov, as DRKS00004741 in the German Clinical trials register, and as EudraCT 2013-000577-77.

## Introduction

*Staphylococcus aureus* bloodstream infection (SAB) affects 20 to 30 persons per 100.000 population annually and has a 3 month mortality between 20% and 30%.^1, 2^ Metastatic foci, late metastatic complications, and relapse are common.^3^ Low-risk SAB defines a subset of patients where risk factors and the clinical course do not point towards an elevated risk for the presence or development of complicated SAB (such as deep-seated infection, prolonged bacteremia, and metastatic foci).^4–6^

The optimal antimicrobial therapy in patients with SAB has been a matter of considerable debate. Guidelines recommend at least 14 days of intravenous antimicrobial therapy in patients with low-risk SAB.^7–9^ An early oral switch antimicrobial therapy has the potential benefit of an abbreviated hospital stay and a reduction of infusion-related complications. However, there is the potential risk of clinical failure if adequate serum concentrations are not reached by oral medication. Furthermore, an earlier outpatient management could potentially lead to a lower compliance with the prescribed drug regimen and to a delayed identification of complications.

Randomized controlled trials directly addressing an early oral switch in SAB are lacking and previous trials have included few participants with SAB receiving oral therapy.^10^ Two recent trials demonstrated non-inferiority of an oral switch in patients with endocarditis^11^ and bone and joint infection.^12^ However, the first trial mainly included participants with other organisms and in the second trial, patients with SAB were excluded. Recent retrospective, observational studies found similar outcomes for oral switch therapy in patients with SAB.^5, 6, 13–19^ However, retrospective studies have a high risk of confounding by indication. Consequently, less than 20% of infectious disease physicians from the US and Canada felt comfortable with switching to oral medication in SAB as two recent surveys demonstrated.^20, 21^

Here we report results of the SABATO trial, a randomized non-inferiority trial to evaluate the efficacy and safety of early oral switch antimicrobial therapy in patients with low-risk SAB as compared to standard intravenous antimicrobial therapy.

## Methods

### TRIAL DESIGN AND OVERSIGHT

We conducted a randomized, parallel-group, open-label, non-inferiority trial. In-patients with low-risk SAB were recruited in 31 sites in four European countries from 20 December 2013 to 22 December 2019 (**Figure 1** and **Supplementary Table S1**). The study protocol was published and approved by the Ethics Committees of all participating centers and the respective competent authorities.^22, 23^ All participants gave written informed consent before enrolment. An independent data monitoring committee periodically assessed the safety of the trial participants. The trial was registered as NCT01792804 in ClinicalTrials.gov, as DRKS00004741 in the German clinical trials register, and as EudraCT 2013-000577-77.

**Figure 1:**
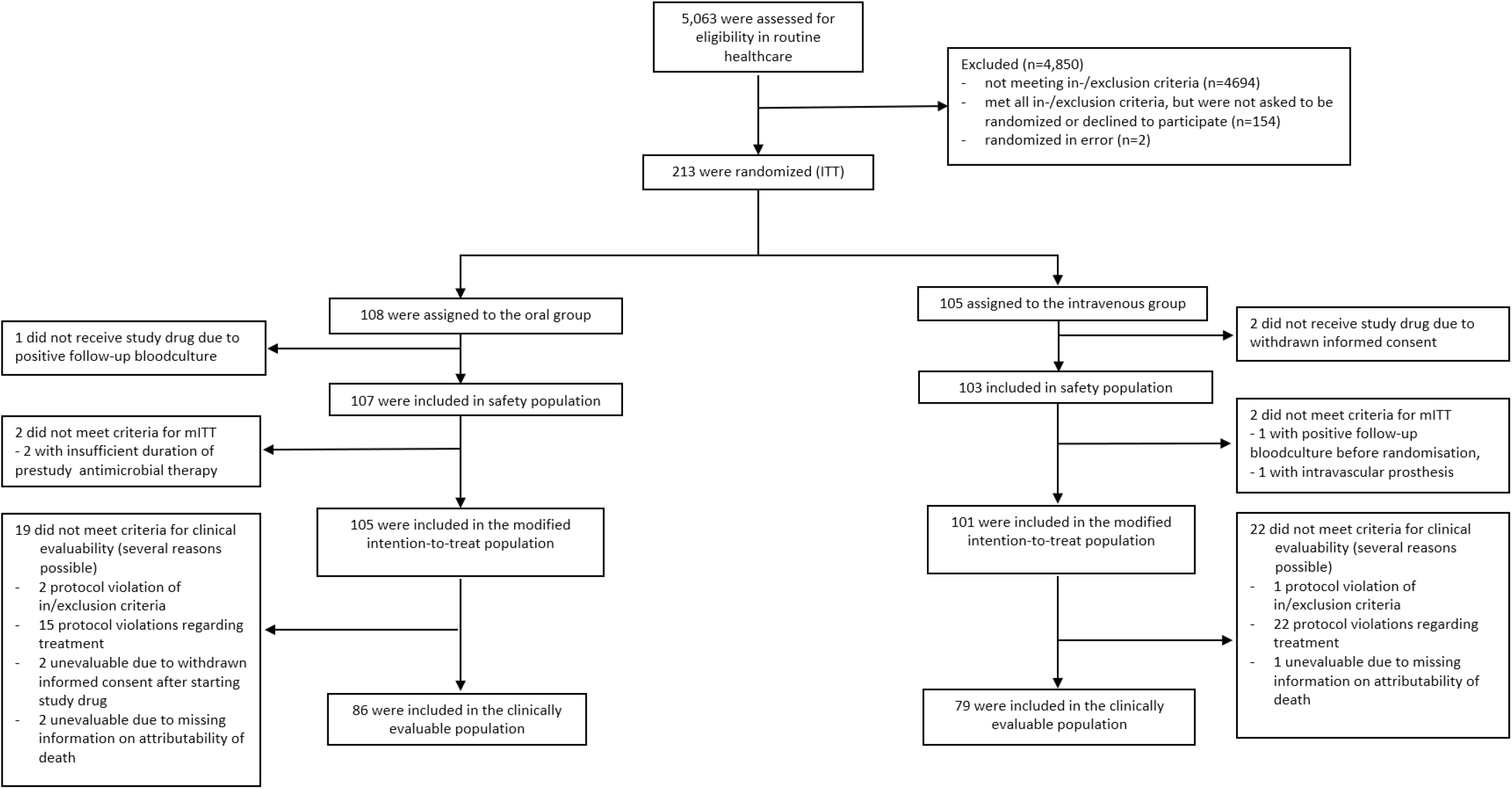
Study flow chart

### PARTICIPANTS

Adult patients (≥18 years) with *S. aureus* isolated from at least one blood culture were eligible, if they had received five to seven days of appropriate intravenous antimicrobial therapy, initiated within 72h after the first positive blood culture was drawn; and follow-up blood cultures obtained within 24-96 hours after the start of appropriate antimicrobial therapy were persistently negative.

Patients were excluded if signs and symptoms of complicated *S. aureus* infection were present prior to enrolment, *i.e.*, deep-seated focus (*e.g.*, endocarditis, pneumonia, infected implant, undrained abscess, empyema, and osteomyelitis), septic shock within 4 days before randomization, prolonged bacteremia (defined as a positive blood culture obtained more than 72 hours after start of adequate antimicrobial therapy), and body temperature >38°C on two separate days within 48 hours before randomization. Patients were excluded, if intravascular catheters were not removed within 4 days after the first positive blood culture was drawn. Patients with a higher *a priori* risk for SAB-related complications were also excluded, namely patients with a recent history of SAB within the preceding 3 months, injection drug use, severe immunodeficiency or immunosuppression, presence of a prosthetic heart valve or deep-seated vascular graft.

Patients with end-stage renal disease, severe liver disease, prosthetic joint, or pacemaker (if implanted more than 6 months prior) were eligible if the infective focus was either a removable intravascular catheter or a skin or soft-tissue infection. In patients with end-stage renal disease or a pacemaker, echocardiography was needed to rule out endocarditis. For details see **Supplementary Table S7**.

### RANDOMIZATION AND MASKING

Participants were randomly allocated to treatment groups (1:1) not earlier than one day before starting study drug through a central internet randomization service TENALEA (stratified by study center, permuted blocks of varying length). Participants and investigators were unmasked regarding treatment. The clinical review committee (CRC), who made the final adjudication, was masked to group assignment.

### PROCEDURES

Participants either received oral switch therapy or intravenous standard therapy. The duration of study therapy was 5 to 7 days to yield a total duration of 14 days of pre-randomization antimicrobial therapy plus study medication.

Antimicrobials were selected by the study physician according to susceptibility results, suspected allergy or intolerance in the following order: oral trimethoprim-sulfamethoxazole 160/800 mg q12h for methicillin-susceptible (MSSA) or methicillin-resistant *S. aureus* (MRSA), clindamycin 600mg q8h for MSSA, and oral linezolid 600mg q12h for MRSA in the oral switch group; intravenous flucloxacillin 2g q6h (cloxacillin 2g q6h in Spain and France), cefazolin 2g q8h, or vancomycin 1g q12h for MSSA, vancomycin 1g q12h or daptomycin 6-10mg/kg q24h for MRSA in the intravenous standard therapy group. The local hospital pharmacy provided the study drug as marketed. Individual adjustments of dosing were possible.

### OUTCOMES

The composite primary endpoint was SAB-related complications within 90 days, defined as relapsing SAB, deep-seated infection with *S. aureus*, or death attributable to SAB. Secondary outcomes were length of hospital stay from the date the first positive blood culture was drawn to hospital discharge, complications of intravenous therapy from start of study drug to end of study, *C. difficile* infection, and 14-, 30-, and 90-day survival. The median length of stay was measured from the first positive blood culture until discharge or death in hospital.

Safety analysis assessed adverse events and serious adverse events. Events were graded according to the Common Terminology Criteria for Adverse Events version 4.0.^24^ Events graded 3 or above were recorded from start of study drug to end of study.

### OUTCOME ASSESSMENT AND STATISTICAL ANALYSIS

The sample size of and the non-inferiority margin were derived as described in the **Supplementary Material**. Due to slow recruitment, the final analysis was performed at half of the initial target sample size, which still accommodated a 10% non-inferiority margin (one-sided α = 0.05, β = 0.2, expected event rate 2.5%, hierarchical testing with Zhao’s test).^25^ According to recommendations from the European Medicines Agency, we initially selected the clinical evaluable population (termed per protocol population in the statistical analysis plan) as primary analysis and the intention-to-treat population as coprimary.^26, 27^ However, recently updated guidance recommends the intention-to-treat population for the primary analysis,^28^ and we therefore report both populations side-by-side, with a focus on the intention-to-treat population.

The intention-to-treat population included all randomly assigned participants. The modified intention-to-treat population comprised participants that were randomized AND received any study drug, excluding participants in whom an inclusion criterion incompatible with low-risk SAB was violated before starting study drug (e.g., persistent positive blood cultures). In both intention-to-treat populations, data were analyzed as assigned with indeterminate and missing outcomes counted as failures.

The clinically evaluable population included all study subjects who were treated according to protocol with study drug, were observed until end-of-follow-up or reached the primary endpoint and did not receive antimicrobial therapy during follow-up that could have masked the primary endpoint. Study drug was considered according to protocol when the total duration of antimicrobial therapy was between 12 and 16 days with at least 5 days of initial intravenous therapy. A masked, independent Clinical Review Committee carefully evaluated study outcomes and treatment.

The safety population included all study subjects who received any study drug. Specifically, participants who ever received an oral antimicrobial agent were compared to participants who never received an oral antimicrobial agent.

A Kaplan-Meier analysis was performed to assess differences in time to SAB-related complications. Hazard ratios were estimated from Cox regression and compared with an unstratified log-rank test. Competing risks were calculated according to Fine *et al* considering non-attributable mortality as competing event.^29^ Statistical calculations were done with the software SAS 9.4 (SAS Inc., Cary, NC, USA) and Stata 17.0 (Stata/SE, College Station, TX, USA).

## Results

### PARTICIPANTS

From December 2013 through December 2019, 213 participants were enrolled into the intention-to-treat population at 31 sites in four countries (**Figure 1** and **Supplementary Table S1 to S3**). The clinically evaluable population consisted of 165 participants, with 86 participants in the oral switch group and 79 participants in the intravenous standard therapy group. The safety population comprised 210 participants who received at least one dose of study drug.

In the intention-to-treat population, baseline characteristics were largely balanced between groups and participants were overall representative of low-risk SAB (**Table 1**). Catheter-related infection and skin-soft-tissue infection were the most prevalent foci, and 16 (7.5%) participants had a MRSA bloodstream infection. There were more participants in the oral switch group with moderate or severe liver disease (10.2% versus 3.8%) and diabetes mellitus (40.7% versus 26.6%). Before starting study medication, participants received a median of 6 days of intravenous antimicrobials in both groups.

**Table 1:**
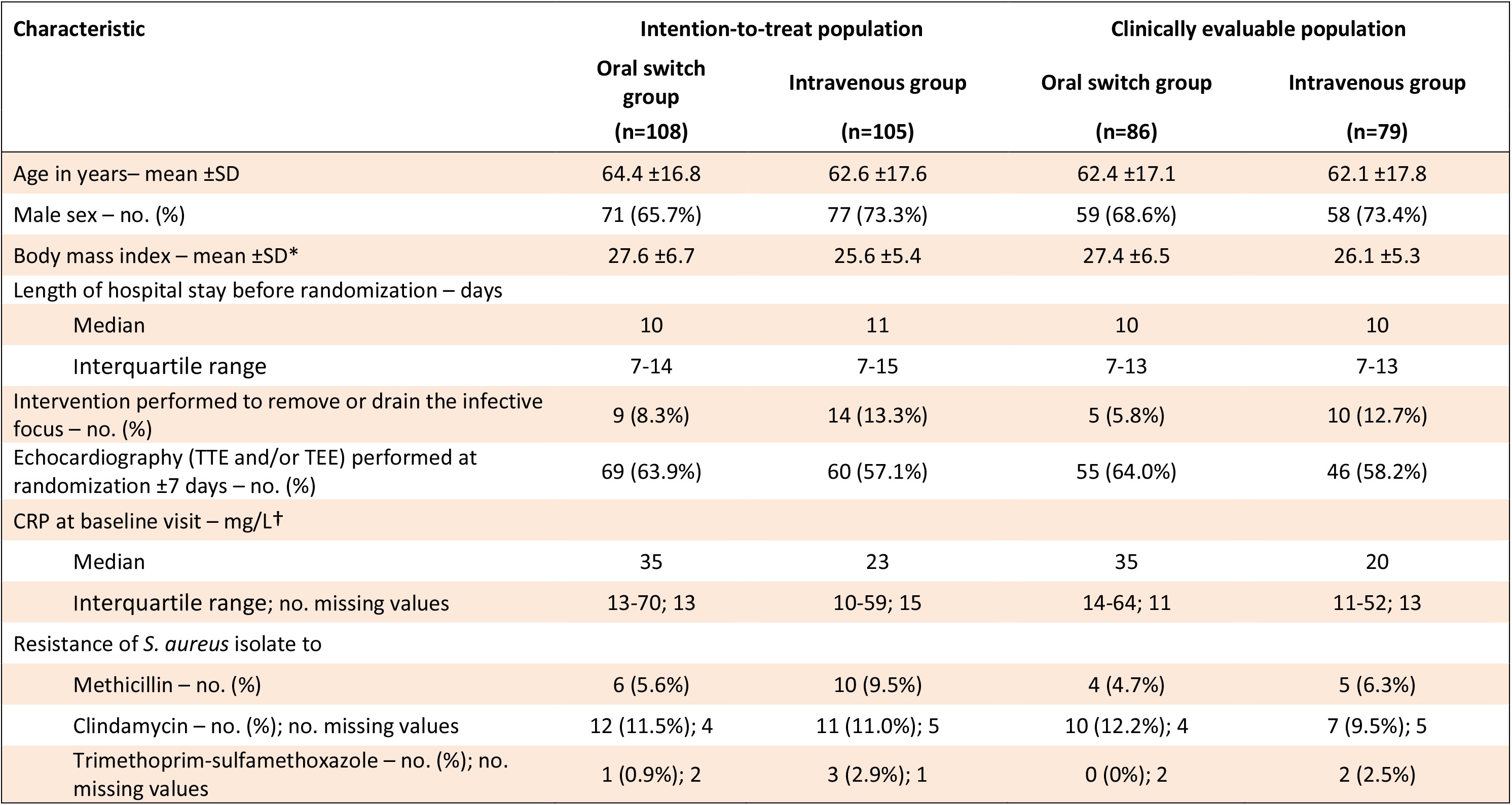
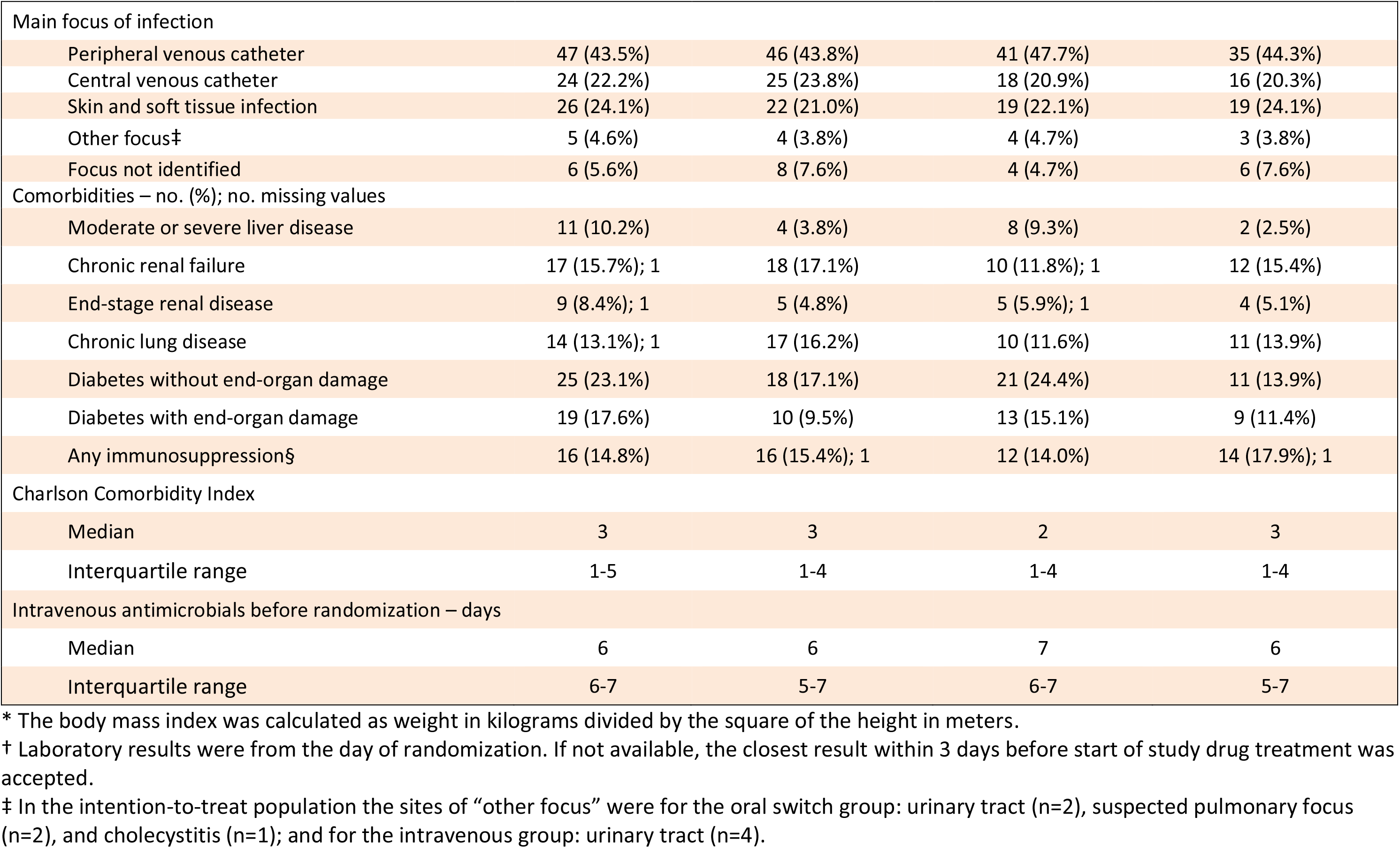
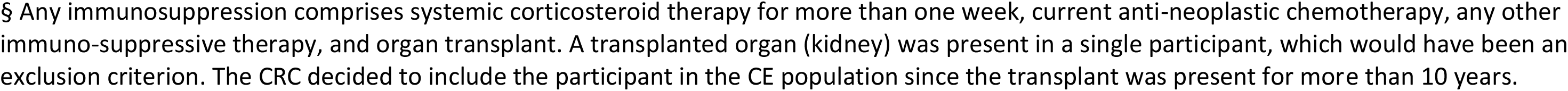
Baseline characteristics of the intention-to-treat (ITT) and the clinically evaluable (CE) population. The number of missing values is given when applicable.

| Characteristic | Intention-to-treat population |  | Clinically evaluable population |  |
| --- | --- | --- | --- | --- |
|  | Oral switch group<br>(n=108) | Intravenous group<br>(n=105) | Oral switch group<br>(n=86) | Intravenous group<br>(n=79) |
| Age in years– mean ±SD | 64.4 ±16.8 | 62.6 ±17.6 | 62.4 ±17.1 | 62.1 ±17.8 |
| Male sex – no. (%) | 71 (65.7%) | 77 (73.3%) | 59 (68.6%) | 58 (73.4%) |
| Body mass index – mean ±SD* | 27.6 ±6.7 | 25.6 ±5.4 | 27.4 ±6.5 | 26.1 ±5.3 |
| Length of hospital stay before randomization – days |  |  |  |  |
| Median | 10 | 11 | 10 | 10 |
| Interquartile range | 7-14 | 7-15 | 7-13 | 7-13 |
| Intervention performed to remove or drain the infective focus – no. (%) | 9 (8.3%) | 14 (13.3%) | 5 (5.8%) | 10 (12.7%) |
| Echocardiography (TTE and/or TEE) performed at randomization ±7 days – no. (%) | 69 (63.9%) | 60 (57.1%) | 55 (64.0%) | 46 (58.2%) |
| CRP at baseline visit – mg/L† |  |  |  |  |
| Median | 35 | 23 | 35 | 20 |
| Interquartile range; no. missing values | 13-70; 13 | 10-59; 15 | 14-64; 11 | 11-52; 13 |
| Resistance of *S. aureus* isolate to |  |  |  |  |
| Methicillin – no. (%) | 6 (5.6%) | 10 (9.5%) | 4 (4.7%) | 5 (6.3%) |
| Clindamycin – no. (%); no. missing values | 12 (11.5%); 4 | 11 (11.0%); 5 | 10 (12.2%); 4 | 7 (9.5%); 5 |
| Trimethoprim-sulfamethoxazole – no. (%); no. missing values | 1 (0.9%); 2 | 3 (2.9%); 1 | 0 (0%); 2 | 2 (2.5%) |
| Main focus of infection |  |  |  |  |
| Peripheral venous catheter | 47 (43.5%) | 46 (43.8%) | 41 (47.7%) | 35 (44.3%) |
| Central venous catheter | 24 (22.2%) | 25 (23.8%) | 18 (20.9%) | 16 (20.3%) |
| Skin and soft tissue infection | 26 (24.1%) | 22 (21.0%) | 19 (22.1%) | 19 (24.1%) |
| Other focus† | 5 (4.6%) | 4 (3.8%) | 4 (4.7%) | 3 (3.8%) |
| Focus not identified | 6 (5.6%) | 8 (7.6%) | 4 (4.7%) | 6 (7.6%) |
| Comorbidities – no. (%); no. missing values |  |  |  |  |
| Moderate or severe liver disease | 11 (10.2%) | 4 (3.8%) | 8 (9.3%) | 2 (2.5%) |
| Chronic renal failure | 17 (15.7%); 1 | 18 (17.1%) | 10 (11.8%); 1 | 12 (15.4%) |
| End-stage renal disease | 9 (8.4%); 1 | 5 (4.8%) | 5 (5.9%); 1 | 4 (5.1%) |
| Chronic lung disease | 14 (13.1%); 1 | 17 (16.2%) | 10 (11.6%) | 11 (13.9%) |
| Diabetes without end-organ damage | 25 (23.1%) | 18 (17.1%) | 21 (24.4%) | 11 (13.9%) |
| Diabetes with end-organ damage | 19 (17.6%) | 10 (9.5%) | 13 (15.1%) | 9 (11.4%) |
| Any immunosuppression§ | 16 (14.8%) | 16 (15.4%); 1 | 12 (14.0%) | 14 (17.9%); 1 |
| Charlson Comorbidity Index |  |  |  |  |
| Median | 3 | 3 | 2 | 3 |
| Interquartile range | 1-5 | 1-4 | 1-4 | 1-4 |
| Intravenous antimicrobials before randomization – days |  |  |  |  |
| Median | 6 | 6 | 7 | 6 |
| Interquartile range | 6-7 | 5-7 | 6-7 | 5-7 |

\* The body mass index was calculated as weight in kilograms divided by the square of the height in meters.
† Laboratory results were from the day of randomization. If not available, the closest result within 3 days before start of study drug treatment was accepted.
‡ In the intention-to-treat population the sites of “other focus” were for the oral switch group: urinary tract (n=2), suspected pulmonary focus (n=2), and cholecystitis (n=1); and for the intravenous group: urinary tract (n=4).
§ Any immunosuppression comprises systemic corticosteroid therapy for more than one week, current anti-neoplastic chemotherapy, any other immuno-suppressive therapy, and organ transplant. A transplanted organ (kidney) was present in a single participant, which would have been an exclusion criterion. The CRC decided to include the participant in the CE population since the transplant was present for more than 10 years.

### ANTIMICROBIAL TREATMENT

Participants were treated with study medication for a median duration of 8 days in both groups, resulting in a full treatment course of antimicrobial medication with a median of 14-days (**Supplementary Table S4**). The most frequently chosen study medications in the oral switch group were trimethoprim-sulfamethoxazole in 63 (58.3%) participants and clindamycin in 35 (32.4%) participants. In the intravenous group, 46 (43.8%) participants received cefazolin and 45 (42.9%) participants received intravenous flucloxacillin or cloxacillin. In 9 (4.2%) participants, initially started study medication was switched to the alternative study medication; 3 (1.4%) participants did not receive study medication.

### OUTCOMES

In the ITT population, the primary composite outcome, *i.e.*, SAB-related complication or a missing outcome within 90 days, occurred in 14/108 participants in the oral switch group and 13/105 in the intravenous standard therapy group (**Table 2**). The treatment-control difference was 0.7 percentage points (95% confidence interval [CI], -7.8% to 9.1%). In the clinical evaluable population, SAB-related complications occurred in 3/86 participants in the oral group versus 4/79 participants in the intravenous group (treatment-control difference -2.9 percentage points; 95% CI, -9.6% to 3.9%). Non-inferiority was achieved in both populations. The results proved robust in a number of sensitivity analyses, which addressed the influence of non-attributable mortality, missing outcome measures, the contribution of SAB-related complications occurring more than 7 days after randomization, and whether SAB-related complications were microbiologically confirmed (**Supplementary Figure S2**). Cumulative incidence plots for the primary outcome with and without non-attributable death as competing risk are shown in **Figure 2**. Details of all participants with SAB-related complications are presented in **Supplementary Table S5.**

**Figure 2:**
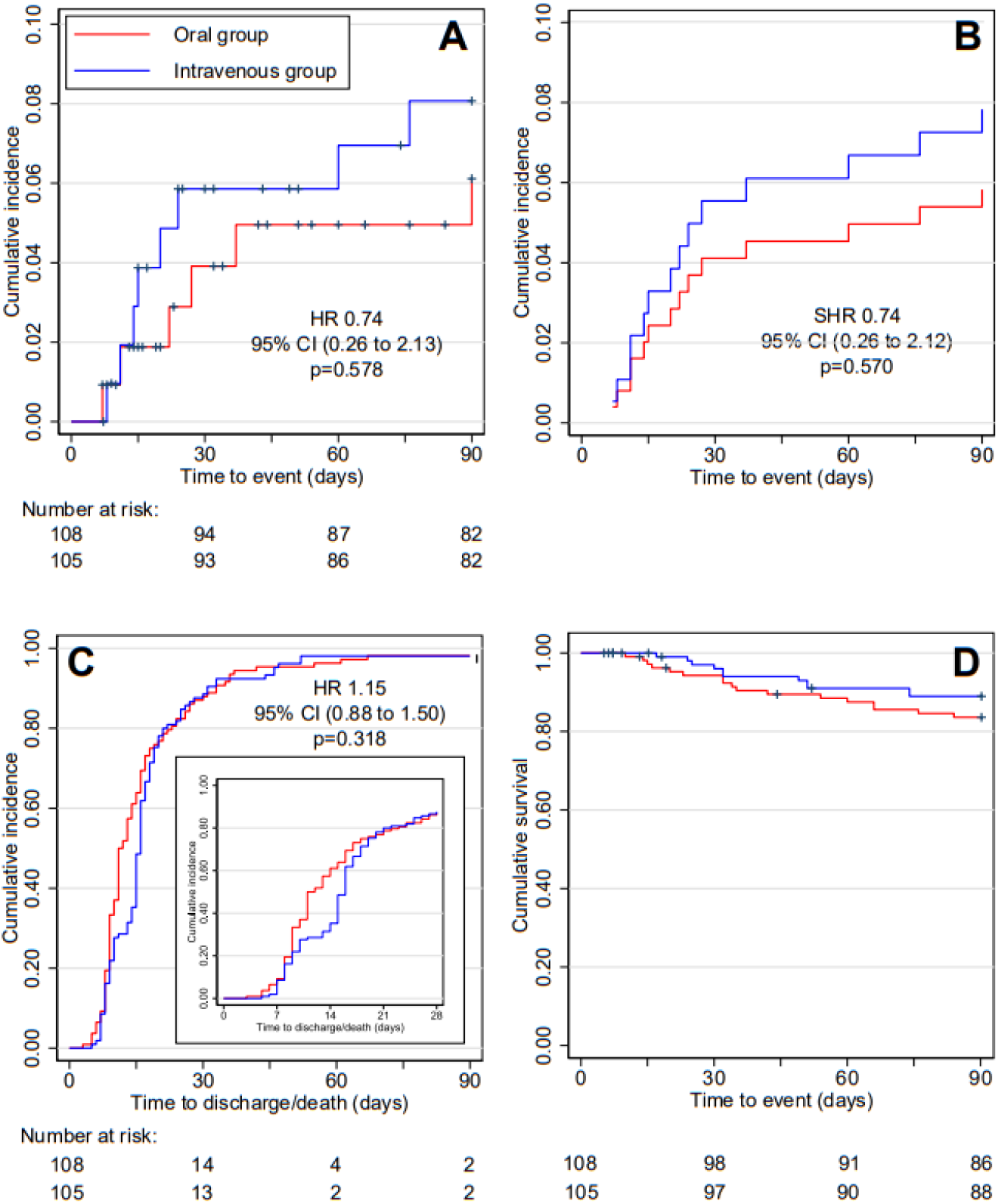
Time course of the primary and secondary outcomes in the intention-to-treat population from onset of SAB. (A) Cumulative incidence of the primary endpoint (SAB-related complications within 90 days) with hazard ratio (HR) estimated from Cox regression. Participants who died from non-attributable death were censored; (B) cumulative incidence of the primary endpoint accounting for non-attributable death as competing event with subdistribution hazard ratio (SHR) estimated from the Fine and Gray model for competing risks; (C) Kaplan-Meier estimates for the length of hospital stay from onset of SAB; discharged participants and participants who died in hospital were counted as events, with hazard ratio estimated from Cox regression; inset: enlarged part that illustrates differences in discharge between study arms. (D) Cumulative survival with hazard ratio (HR) estimated from Cox regression.

**Table 2:**
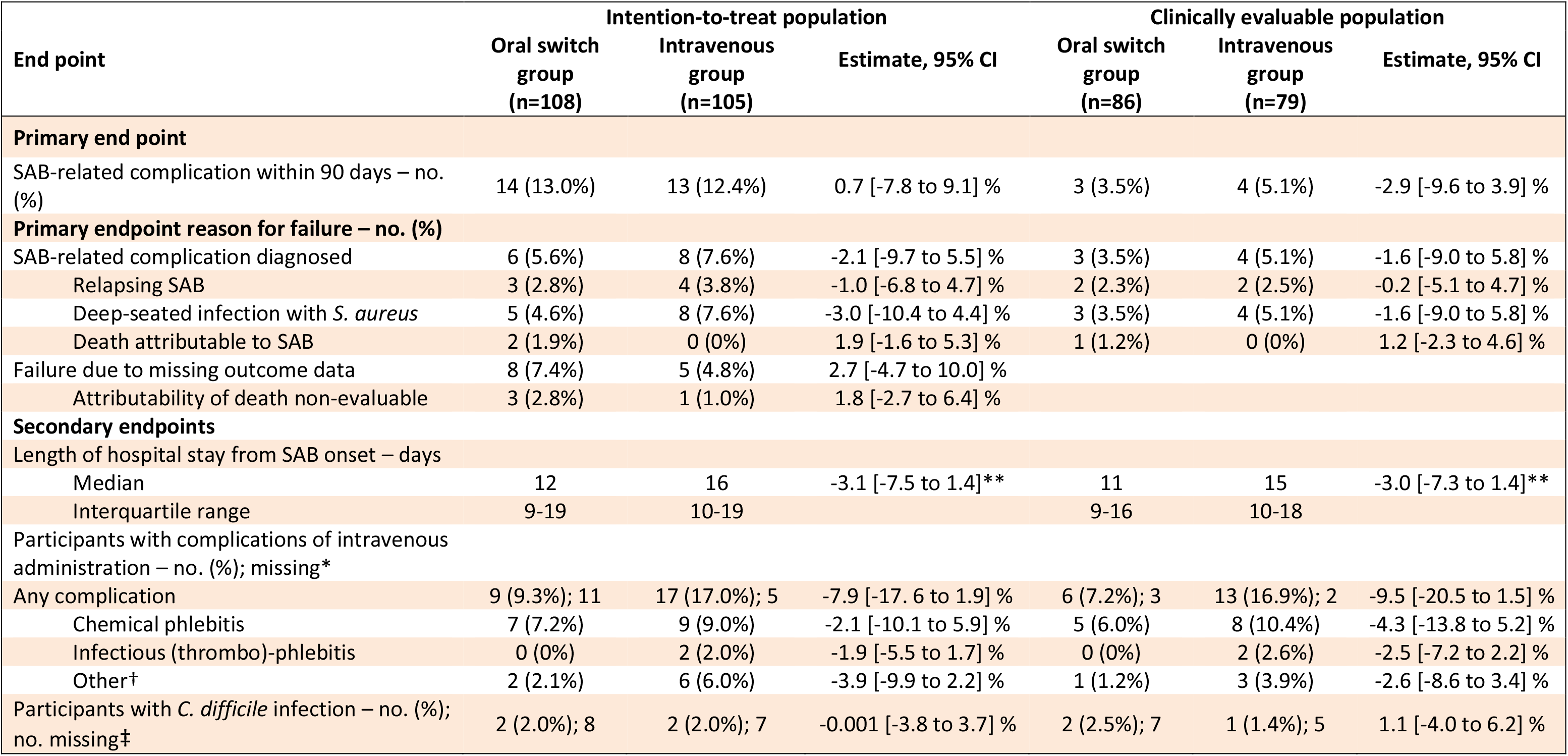
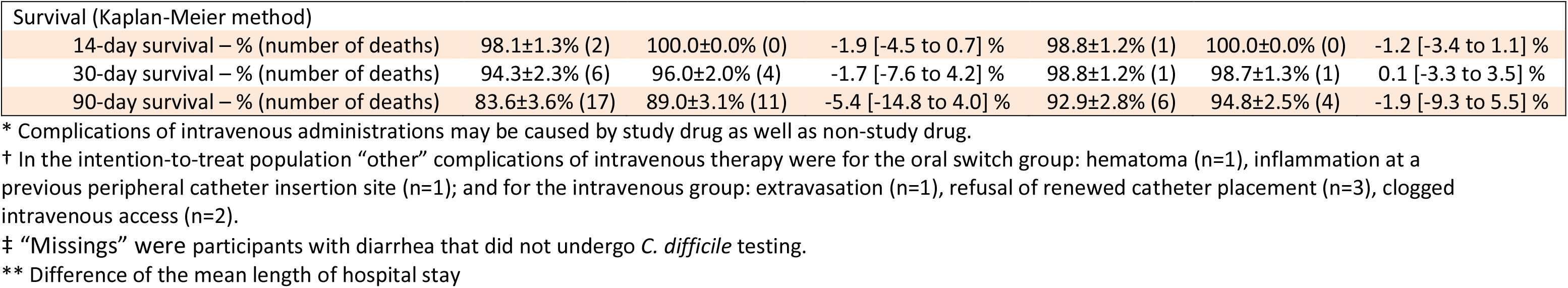
Primary and secondary outcome variables for the intention-to-treat and the clinically evaluable population. Estimates denote the treatment difference between the oral and intravenous group; negative values favour oral treatment (except for survival). SAB, *Staphylococcus aureus* bloodstream infection

| End point | Intention-to-treat population |  |  | Clinically evaluable population |  |  |
| --- | --- | --- | --- | --- | --- | --- |
|  | Oral switch group (n=108) | Intravenous group (n=105) | Estimate, 95% CI | Oral switch group (n=86) | Intravenous group (n=79) | Estimate, 95% CI |
| **Primary end point** |  |  |  |  |  |  |
| SAB-related complication within 90 days – no. (%) | 14 (13.0%) | 13 (12.4%) | 0.7 [-7.8 to 9.1] % | 3 (3.5%) | 4 (5.1%) | -2.9 [-9.6 to 3.9] % |
| **Primary endpoint reason for failure – no. (%)** |  |  |  |  |  |  |
| SAB-related complication diagnosed | 6 (5.6%) | 8 (7.6%) | -2.1 [-9.7 to 5.5] % | 3 (3.5%) | 4 (5.1%) | -1.6 [-9.0 to 5.8] % |
| Relapsing SAB | 3 (2.8%) | 4 (3.8%) | -1.0 [-6.8 to 4.7] % | 2 (2.3%) | 2 (2.5%) | -0.2 [-5.1 to 4.7] % |
| Deep-seated infection with *S. aureus* | 5 (4.6%) | 8 (7.6%) | -3.0 [-10.4 to 4.4] % | 3 (3.5%) | 4 (5.1%) | -1.6 [-9.0 to 5.8] % |
| Death attributable to SAB | 2 (1.9%) | 0 (0%) | 1.9 [-1.6 to 5.3] % | 1 (1.2%) | 0 (0%) | 1.2 [-2.3 to 4.6] % |
| Failure due to missing outcome data | 8 (7.4%) | 5 (4.8%) | 2.7 [-4.7 to 10.0] % |  |  |  |
| Attributability of death non-evaluable | 3 (2.8%) | 1 (1.0%) | 1.8 [-2.7 to 6.4] % |  |  |  |
| **Secondary endpoints** |  |  |  |  |  |  |
| Length of hospital stay from SAB onset – days |  |  |  |  |  |  |
| Median | 12 | 16 | -3.1 [-7.5 to 1.4]** | 11 | 15 | -3.0 [-7.3 to 1.4]** |
| Interquartile range | 9-19 | 10-19 |  | 9-16 | 10-18 |  |
| Participants with complications of intravenous administration – no. (%); missing* |  |  |  |  |  |  |
| Any complication | 9 (9.3%); 11 | 17 (17.0%); 5 | -7.9 [-17.6 to 1.9] % | 6 (7.2%); 3 | 13 (16.9%); 2 | -9.5 [-20.5 to 1.5] % |
| Chemical phlebitis | 7 (7.2%) | 9 (9.0%) | -2.1 [-10.1 to 5.9] % | 5 (6.0%) | 8 (10.4%) | -4.3 [-13.8 to 5.2] % |
| Infectious (thrombo)-phlebitis | 0 (0%) | 2 (2.0%) | -1.9 [-5.5 to 1.7] % | 0 (0%) | 2 (2.6%) | -2.5 [-7.2 to 2.2] % |
| Other† | 2 (2.1%) | 6 (6.0%) | -3.9 [-9.9 to 2.2] % | 1 (1.2%) | 3 (3.9%) | -2.6 [-8.6 to 3.4] % |
| Participants with *C. difficile* infection – no. (%); no. missing‡ | 2 (2.0%); 8 | 2 (2.0%); 7 | -0.001 [-3.8 to 3.7] % | 2 (2.5%); 7 | 1 (1.4%); 5 | 1.1 [-4.0 to 6.2] % |

| Survival (Kaplan-Meier method) |  |  |  |  |  |  |
| --- | --- | --- | --- | --- | --- | --- |
| 14-day survival – % (number of deaths) | 98.1±1.3% (2) | 100.0±0.0% (0) | -1.9 [-4.5 to 0.7] % | 98.8±1.2% (1) | 100.0±0.0% (0) | -1.2 [-3.4 to 1.1] % |
| 30-day survival – % (number of deaths) | 94.3±2.3% (6) | 96.0±2.0% (4) | -1.7 [-7.6 to 4.2] % | 98.8±1.2% (1) | 98.7±1.3% (1) | 0.1 [-3.3 to 3.5] % |
| 90-day survival – % (number of deaths) | 83.6±3.6% (17) | 89.0±3.1% (11) | -5.4 [-14.8 to 4.0] % | 92.9±2.8% (6) | 94.8±2.5% (4) | -1.9 [-9.3 to 5.5] % |

\* Complications of intravenous administrations may be caused by study drug as well as non-study drug.
† In the intention-to-treat population “other” complications of intravenous therapy were for the oral switch group: hematoma (n=1), inflammation at a previous peripheral catheter insertion site (n=1); and for the intravenous group: extravasation (n=1), refusal of renewed catheter placement (n=3), clogged intravenous access (n=2).
‡ “Missings” were participants with diarrhea that did not undergo *C. difficile* testing.
\*\* Difference of the mean length of hospital stay

The length of stay after the first positive blood culture was shorter in the oral switch group with a median stay of 12 days versus 16 days (mean difference -3.1 days [95% CI, -7.5 to 1.4], **Table 2**). Orally treated participants had fewer complications of intravenous therapy (treatment difference - 7.9% [95% CI, -17.6% to 1.9%]). The incidence of *C. difficile* infection was similar in both groups. Survival was lower in the oral switch group than in the intravenous group with a treatment difference of-1.73% [95% CI, -7.61 to 4.15] at 30 days, and -5.36% [95% CI, -14.75 to 4.03] at 90 days.

### SUBGROUP ANALYSIS

Analyses of the primary endpoint were similar, regardless of whether the bacterial isolate was methicillin-susceptible or resistant, or whether the participants had different foci at baseline, a Charlson comorbidity index <3 or ≥3, had echocardiography performed or not, or were enrolled in different countries (**Supplementary Figure S1**).

### SAFETY OUTCOMES

The safety population consisted of 210 participants who received at least one dose of study drug. Overall, 94 (44.8%) participants reported an adverse event; 63 (30.0%) were serious adverse events (**Table 3, Supplementary Table S6**). The most common adverse events were infections with 45 events recorded in 40 (19.0%) participants. Participants in the oral group reported more adverse events and serious adverse events than participants in the intravenous group. Serious adverse events occurred with a rate of 0.21 per person-month (95% CI, 0.15 to 0.29) in the oral group and 0.14 (95% CI, 0.10 to 0.20) in the intravenous group, with a risk ratio of 0.65 (95% CI, 0.40 to 1.05; p=0.079).

**Table 3:** Adverse events and serious adverse events in the safety population occurring within 90 days. Common Terminology Criteria for Adverse Events (CTCAE) were recorded from grade 3 and above and listed by system organ class and preferred term. System organ class terms for serious adverse events occurring in more than 5% of participants and preferred terms with at least two serious adverse events are shown. Preferred terms were aggregated as denoted. P: Fisher’s exact test comparing the number of participants with at least one event.

| Event | Oral switch group<br>(n=107) |  | Intravenous group<br>(n=103) |  | p |
| --- | --- | --- | --- | --- | --- |
|  | Participants<br>with events –<br>no. (%) | Total<br>events<br>– no. | Participants<br>with events –<br>no. (%) | Total<br>events<br>– no. |  |
| **Any adverse event** | 52 (48.6%) | 89 | 42 (40.8%) | 61 | 0.270 |
| **Any serious adverse event** | 36 (33.6%) | 60 | 27 (26.2%) | 40 | 0.292 |
| Drug-related* | 3 (2.8%) | 3 | 0 | 0 |  |
| **Infections and infestations** | 23 (21.5%) | 27 | 17 (16.5%) | 18 | 0.384 |
| Sepsis, septic shock,<br>bacteraemia, endocarditis |  | 7 |  | 5 |  |
| Urinary tract infection,<br>cystitis |  | 5 |  | 3 |  |
| Respiratory tract infection,<br>pneumonia, bronchitis |  | 4 |  | 2 |  |
| Wound infection, skin<br>infection |  | 4 |  | 2 |  |
| Arthritis, osteomyelitis |  | 2 |  | 2 |  |
| *C. difficile* infection |  | 1 |  | 1 |  |
| **General disorders and**<br>**administration site conditions** | 11 (10.3%) | 13 | 4 (3.9%) | 4 | 0.106 |
| Fever |  | 5 |  | 1 |  |
| Oedema |  | 3 |  | 0 |  |
| Asthenia, general physical<br>deterioration |  | 2 |  | 1 |  |
| Severe Pain |  | 2 |  | 0 |  |
| **Cardiac disorders** | 9 (8.4%) | 9 | 5 (4.9%) | 5 | 0.409 |
| Cardio-(respiratory) arrest |  | 4 |  | 1 |  |
| Cardiac failure |  | 1 |  | 3 |  |
| Myocardial ischemia,<br>myocardial infarction |  | 2 |  | 0 |  |
| **Respiratory, thoracic and**<br>**mediastinal disorders** | 4 (3.7%) | 4 | 10 (9.7%) | 10 | 0.101 |
| Pulmonary embolism |  | 1 |  | 2 |  |
| Dyspnoea |  | 1 |  | 1 |  |
| Exacerbated chronic<br>obstructive pulmonary<br>disease |  | 1 |  | 1 |  |

\* Drug related serious adverse events (classified as certain or probable/likely) were cardiac failure, *C. difficile* colitis, and fever.

## Discussion

In this multicenter randomized trial of patients with low-risk SAB, early oral switch antimicrobial therapy was non-inferior to intravenous standard therapy with regard to the primary endpoint. Participants on oral medication were discharged from hospital earlier than participants on intravenous medication, as expected from previous trials on oral switch antimicrobial therapy in patients with various infections.^11, 30^ We further found that a switch to oral medication reduced the rate of complications of intravenous administration by 7.9 percentage points (95% CI, -17.6 to 1.9).

Survival at 90 days was slightly lower in the oral switch group across all analyses (**Table 2**), albeit not statistically significant. The two participants with death attributable to SAB were in the oral group. Both had catheter-related SAB and developed disseminated infection within 3 weeks after starting study medication. One participant was discharged and declined readmission when her general condition deteriorated. This highlights the importance of monitoring patients with SAB closely for developing complications.

We further observed a higher incidence of adverse events and serious adverse events in the oral switch group, with the overall incidence being in the range of previous trials.^31, 32^ A large proportion of adverse events were infections not caused by *S. aureus*, which underscores that the participant population is generally at risk for infections.

Assigning the low-risk category to patients with SAB can be challenging. In a RCT in patients with uncomplicated staphylococcal bacteremia, Holland *et al.* found that one-third of patients without suspected metastatic infection at enrollment were ultimately diagnosed with complicated SAB.^32^ With our criteria for low-risk SAB, we observed 14 (7%) SAB-related complications, of which more than half became apparent during the intervention phase, i.e. during the second week of illness. These early complications may have been present at baseline and – unlike in our study – it has been proposed not to consider them as treatment failures in RCTs.^33^ When we excluded early complications from the primary endpoint, we confirmed non-inferiority (**Supplementary Table S2**).

The strengths of the trial were the pragmatic approach with globally available oral medication as first choice, the strict definition of low-risk SAB, and the choice of a composite primary endpoint specific to SAB.

The trial has several limitations. It was open-label, since intravenous administration of placebo in the oral switch group would have interfered with the aim of facilitating early discharge. The large number of patients screened raises the question, whether low-risk SAB is a clinically relevant entity. However, screening-to-enrolment ratios above 1:25 are typical for trials on uncomplicated SAB,^33^ and upcoming platform trials may allow more efficient recruitment.^34^ Further, the trial was terminated due to slow recruitment and did not reach the initially planned sample size. Nevertheless, results were sufficiently precise to draw robust conclusions.

Possibly, a more aggressive diagnostic approach, *e.g.* involving FDG-PET/CT,^35^ a more stringent evaluation of the need for echocardiography such as the VIRSTA score,^36^ or novel biomarkers^37^ may allow to more accurately identify and predict deep-seated infections. Nevertheless, careful assessment of the individual patient, preferably by consulting an infectious diseases expert, is necessary to correctly assign a patient to the low-risk group and close monitoring for early detection of complications and potential prolongation of antimicrobial therapy is advised.

In summary, we found that an early oral switch was non-inferior to standard intravenous antimicrobial therapy in low-risk SAB. Provided a rigorous clinical assessment and close monitoring for complications, this study supports a change in treatment guidelines to allow for the possibility of an early oral switch antimicrobial therapy in patients with thoroughly documented low-risk SAB.

## Contributors

AJK was the Coordinating Principal Investigator and sponsor’s representative. GF served as “Leiter klinische Prüfung” until July 2016. AJK, HS, WVK, SR, MH conceived and designed the study. AJK, AR, MN managed the trial. AJK, LELC, JRB, JMC, MDN, GF, NJ, WVK, SR, RL, LC, LB, AL, KK, CM, AS, SH, BF, ML, JPT, AD, TG, DB, TW, SR, JK, MLM, EF, HS, FM, and PT were site investigators, involved in participant recruitment and data collection. MH, AA, and AJK developed the statistical analysis plan. AJK, AR, AA, and MH performed data curation. MH and AA did the formal statistical analysis. AJK, MH, AA, and HS wrote the first draft of the manuscript, which was reviewed by all authors.

## Further Members of the SABATO trial group with linked authorship in pubmed

Adoración Valiente, Marina de Cueto (Clinical Unit of Infectious Diseases and Microbiology, Hospital Universitario Virgen Macarena, Department of Medicine, University of Sevilla, Biomedicine Institute of Sevilla (IBiS) / CSIC, CIBER de Enfermedades Infecciosas (CIBERINFEC), Spain), Ángel Rodríguez (Clinical Microbiology Department (UCEIMP), Virgen del Rocío University Hospital / Institute of Biomedicine of Seville (IBiS)/CSIC/University of Seville/CIBERINFEC, Seville, Spain), José Molina (Infectious Diseases Department (UCEIMP), Virgen del Rocío University Hospital / Institute of Biomedicine of Seville (IBiS) / CSIC / University of Seville /C IBERINFEC, Seville, Spain), Julia Fischer (Department I of Internal Medicine, Division of Infectious Diseases, University of Cologne, Kerpener Str. 62, 50937 Köln, Germany), Gregor Paul (Department I of Internal Medicine, Division of Infectious Diseases, University of Cologne, Kerpener Str. 62, 50937 Köln, Germany), Reinhild Prinz-Langenohl (Clinical Trials Centre Cologne (CTCC), University of Cologne, Gleueler Str. 269, 50924 Cologne, Germany), Gabriele Peyerl-Hoffmann, Daniel Hornuß (Division of Infectious Diseases, Department of Medicine II, Faculty of Medicine and University Medical Centre Freiburg, 79106 Freiburg, Germany), Sébastien Gallien (Infectious Diseases Department, Henri-Mondor University Hospital, AP-HP, 94000 Creteil, France), Vincent Fihman (Bacteriology and Infection Control Unit, Department of Prevention, Diagnosis, and Treatment of Infections, Henri-Mondor University Hospital, AP-HP, 94000 Creteil, France), Marion Lacasse (Service de Médecine Interne et Maladies Infectieuses, Centre Hospitalier Régional Universitaire de Tours, 37000 Tours, France), Francois Coustillères (Service de Médecine Interne et Maladies Infectieuses, Centre Hospitalier Régional Universitaire de Tours, 37000 Tours, France), Christian Becker (Helios Klinikum Krefeld, Lutherplatz 40, 47805 Krefeld, Germany), André Fuchs (Internal Medine III - Gastroenterology and Infectious Diseases, University Hospital of Augsburg, Germany), Laura Morata (Department of Infectious Diseases, Hospital Clínic, IDIBAPS, Barcelona, Spain), Sebastian Weis (Institute for Infectious Diseases and Infection Control, Jena University Hospital – Friedrich Schiller University Jena, Am Klinikum 1, 07747 Jena, Germany), Diane Ponscarme (Infectious Diseases Department, Saint-Louis hospital, 1 avenue Claude Vellefaux 75010, Paris, France), Lydie Khatchatourian (Department of Infectious Diseases, Centre Hospitalier de Cornouaille, Quimper, France), Elisabeth Rouveix (Internal Medicine Department, Ambroise Paré University Hospital, 92100 Boulogne-Billancourt, France), Dominique Merrien (Infectious diseases department, CHD Vendée, Boulevard Stéphane Moreau, 85925 La Roche-sur-Yon, France), Raphaël Lecomte (Unité maladies infectieuses et tropicales, CHU de Nantes, 1, Place Alexis Ricordeau, Nantes, France), Jacobien Veenemans (Department of Medical Microbiology, Amphia Hospital Breda, Molengracht 21, 4818 CK Breda, The Netherlands), Helem H. Vilchez (Infectious Diseases Unit, Internal Medicine Department, Hospital Universitari Son Espases, Fundació Institut d’Investigació Sanitària Illes Balears (IdISBa), 07120 Palma de Mallorca, Spain), Violaine Tolsma (Service de maladies infectieuses, Centre Hospitalier d’Annecy Genevois, 1 Avenue de l’Hôpital, 74370 Epagny Metz-Tessy France), Johanna Kessel (Medical Clinic: Haematology, Oncology, Haemostaseology, Rheumatology, Infectious Diseases, University Hospital Frankfurt, Theodor-Stern-Kai 7, 60590 Frankfurt, Germany), Marc J.M. Bonten (Julius Center for Health Sciences and Primary Care, UMC Utrecht, Utrecht University, Utrecht, The Netherlands), Jan Rupp (Clinic for Infectious Diseases and Microbiology, University Hospital Schleswig-Holstein, Campus Lübeck, Ratzeburger Allee 160, 23538 Lübeck, Germany), Laurent Hocqueloux (Service des Maladies Infectieuses, Centre Hospitalier Régional, Orléans, France), Frederic Lucht (Clinical investigation center-Inserm-CIC1408, hôpital Nord, CHU de Saint-Étienne, 42055 Saint-Étienne cedex 02, France), Jean-Paul Stahl (Infectious diseases, CHU Grenoble, 38043 Grenoble, France), Anne L.M. Vlek (Department of Medical Microbiology and Immunology, Diakonessenhuis, Utrecht, The Netherlands)

## Data sharing

Data collected for the study will be available (for Details see Supplementary Material).

## Supporting information

Study Protocol

Statistical Analysis Plan

## Data Availability

All data produced in the present study are available upon reasonable request to the authors as detailed in the supplementary material.

## Acknowledgements

The trial was funded by the Deutsche Forschungsgemeinschaft (project number 217917502 to A.J.K.). We thank participants and staff from all sites who contributed to the trial (listed in the **Supplementary Material**). We thank Vance G. Fowler, Stephan Harbarth, and Guy Thwaites for expert opinion in the Trial Steering Committee, Vincent LeMoing, Miquel Pujol Rojo, and Estée Török for participation in the Clinical Review Committee, and Walter E. Häfeli, Alexandra Heininger, Geraldine Rauch for participation in the Data Monitoring Committee.

## Supplement

Supplementary Material

Study Protocol

Statistical Analysis Plan

