## Supplementary Material for "Early oral switch in low-risk *Staphylococcus aureus* bloodstream infection"

Supplement to:

**Early oral switch in low-risk *Staphylococcus aureus* bloodstream infection: a randomized, open-label, phase 3, non-inferiority trial (SABATO)**

Provided by the authors for additional information about the work. More details on the study methodology are available in the trial protocol and the statistical analysis plan.

#### Section 1: SABATO Investigators

__Participating centers__

**Germany**

**Cologne University Clinics:** G. Fätkenheuer, N. Jung, H. Seifert, J. Fischer, G. Paul, C. Bernasch, S. Margane, K. Fiddike, A. Kaasch
**Freiburg University Hospital:** W.V. Kern, S. Rieg, D. Hornuß, *I. Joost, G. Peyerl-Hoffmann*
**Vivantes Hospital, Berlin:** H. Stocker, K. Arastéh, *B. Krondorfer*
**Helios Clinic Krefeld:** K. Kösters, C. Becker, *T. Frieling*
**Hannover Medical School:** T. Welte, *J. Freise, C. Mölgen, A. Bergner, N. Scharf*
**Jena University Hospital:** M. Pletz, S. Beier, S. Hagel, S. Weis, *J. Schmidt*
**University of Schleswig-Holstein, Lübeck:** J. Rupp, *K. Dahlhoff, D. Gadji, D. Lenke, N. Käding*
**Leverkusen Hospital:** S. Reuter, *H. Faber, F. Mandraka, P.-I. Scharrenbroich, D. Demircan*
**University Hospital Frankfurt:** C. Stephan, J. Kessel, *T. Wolf, F. Ebeling, C. Wengenroth*
**University Hospital Düsseldorf:** A. Kaasch, A. Rommerskirchen, C. Mackenzie, A. Fuchs, S. Janetzki

**The Netherlands**

**University Medical Center Utrecht:** M. Bonten, *J. Oosterheert, M. Ekkelenkamp, V. Schweitzer*
**Amphia Hospital, Breda**: J. Kluytmans, J. Veenemans
**Diakonessenhuis Utrecht:** A. Vlek, *A. van der Bij, S. Sankatsing*

**Spain**

**Hospital Clínic of Barcelona:** A. Soriano, *L. Morata, M. Solé*
**Hospital Universitario Virgen Macarena Sevilla:** J. Rodríguez-Baño, L. E. López-Cortés, M. Nuñez, E. Moreno, M. de Cueto, A. Valiente, M. Macías
**Hospital Universitario Virgen del Rocío Sevilla:** J. Cisneros, M. J. Gómez-Gómez, R. Alvarez-Marín, J. A. Lepe, J. Molina
**Hospital Universitario Son Espases Palma de Mallorca:** M. Riera, E. Ruiz de Gopegui Bordes, M. L. Martin, H. H. Vilchez, J.L. Perez

**France**

**APHP Ambroise Paré, Boulogne-Billancourt:** E. Rouveix-Nordon, C. Duran, A. Dinh, *M. Lachâtre*, *D. Armougom*
**APHP Beaujon, Clichy:** B. Fantin, *V. De, N. Gamay, S. Zaher*
**APHP Henri Mondor, Créteil:** R. Lepeule, A. Moussafeur, L. Coutte, S. Gallien, V. Fihman
**APHP St. Louis, Paris:** J. Molina, N. de Castro, A.-L. Munier, M. Noret, D. Ponscarme, M. Lafaurie, *B. Denis, M. Siguier, T. Delory, M. Alice, D. Sardou*
**Centre Hospitalier Annecy Genevois:** J. Gaillat, V. Vitrat, G. Clavere, J.P. Bru, C. Janssen, E. Piet, V. Tolsma, A.L. Destrem, M. Maillet
**CH Métropole Savoie, Chambéry:** E. Forestier, O. Rogeaux, *A. Bosch, C. Descotes-Genon, MC Carret, T. Habet*
**CHU St. Etienne:** F. Lucht, *E. Botelho-Nevers, C. Cazorla, A. Fresard, M. Lutz, A. Gagneux-Brunon*
**CHU Grenoble Alpes:** J.-P. Stahl, *O. Epaulard, S. Touati, M. Daoukhi, P. Pavese, J. Brion, A. Mounayar*
**CH Orléans:** L. Hocqueloux, *T. Prazuck, J. Buret, C. Mille, C. Gubavu, A. Seve, B. De Dieulevelt, C. Boulard, G. Thomas*
**CHRU Tours:** L. Bernard, A. Lemaignen, M. Lacasse, F. Coustillères, *M. Hallouin-Bernard, C. Carvhalo, F. Bastides, Z. Maakaroun, G. Gras, B. Lioger*
**CHU Nantes:** D. Boutoille, A.-S. Lecompte, R. Lecomte, C. Deschanvres, *C. Biron, M Lefebvre, J. Brochard, B. Gaborit, S. Delaure, J. Orain*
**CH De Cornouaille, Quimper:** J.-P. Talarmin, L. Khatchatourian, N. Saïdani, F. Le Gall, M.-S. Fangous, *M. Brière, P. Rameau, S. Jolas, M. Le Donge*

**CHU Pontchaillou, Rennes:** P. Tattevin, E. Ouamara-Digue, S. Limonta*, M. Revest, M. Baldeyrou, E. Thébault*
**CH Départemental de Vendée, La Roche sur Yon:** T. Guimard, H. Pelerin, M. Morrier, E. Migne, D. Merrien, *H. Durand, A. Pegeot, S. Leautez-Nainville*

**Trial coordination and oversight**
**Cologne Clinical Trial Center**: R. Prinz-Langenohl, P. Schmalz, U. Paulus, L. Scharfenberger, H. Haddadi, K. Eggers, A. Krema, A. Pfeiffer, F. Schwartzkopf
**Heinrich-Heine-University Düsseldorf:** A. Rommerskirchen, A.J. Kaasch, S. Janetzki
**Utrecht Medical Center:** W. Pujik
**AMC Amsterdam:** J. Dalen, M. Hoefkens
**Hospital Clinic, Barcelona**: J Arnaiz, S. Varea, L. Caballero, L. Burunat
**RENARCI, Annecy:** M. Noret
**Statistical Support, University of Cologne:** M. Hellmich, A. Adams, V. Weiß
**Scientific Advisory Committee/Trial Steering Committee:** V. G. Fowler, S. Harbarth, and G. Thwaites (independent); A. J. Kaasch, H. Seifert, W. V. Kern, S. Rieg, G. Fätkenheuer, M. Hellmich, A. Soriano, M. Bonten, and P. Tattevin (investigators)
**Independent Data Monitoring Committee:** Walter E. Haefeli, Alexandra Heininger, Geraldine Rauch
**Independent Clinical Review Committee:** Vincent LeMoing, Miquel Pujol Rojo, and Estée Török

### Section 2: Methods

#### Study Overview

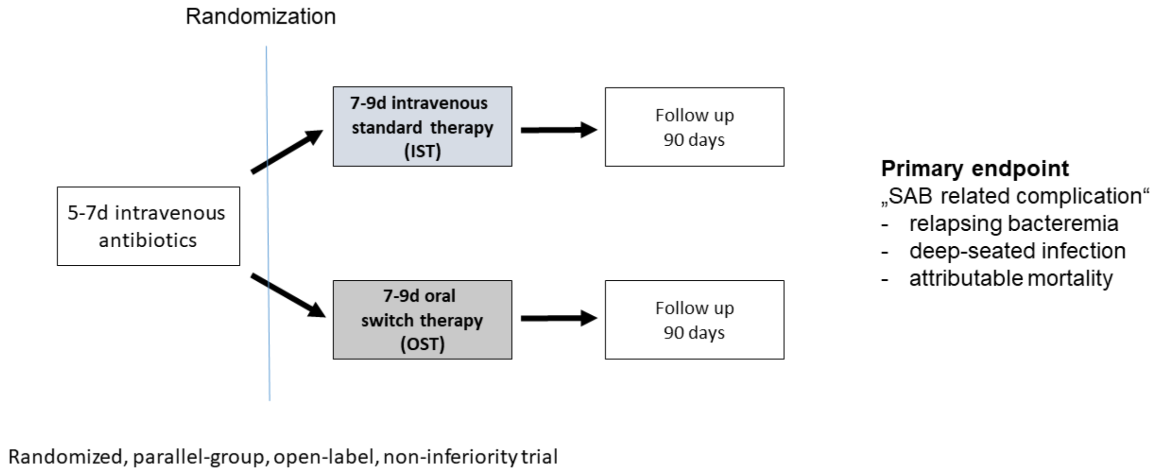

#### Non-inferiority margin selection and sample size

The trial was originally designed with a 5% non-inferiority margin, which yielded a total sample size of 430 participants (one-sided $\alpha = 0.05$, $\beta = 0.2$, one interim analysis at information fraction 0.5 (i.e. 215 participants) using the O'Brien-Fleming bound 2.373, 2.5% of participants reaching the endpoint).

To accommodate conflicting views about the size of the non-inferiority margin (*i.e.* 5% or 10%) we planned a sequentially rejective testing procedure (hierarchical testing, stratified by centre) based on Zhao's test,[1] from weaker to stronger assertions. This means, that first the null-hypothesis is tested with a 15% non-inferiority margin and a 90% confidence interval. If the null-hypothesis is rejected, then the test is repeated with a 15% non-inferiority margin and a 95% confidence interval; if rejected again, then with a 10% non-inferiority margin and a 90% confidence interval, and so on, until finally a 5% non-inferiority margin and a 95% confidence interval is tested.

Slow enrolment necessitated termination of the trial before reaching the sample size of 430 participants. Since it seemed impossible to complete the full sample size without massively increasing the number of centers and there were no funds and infrastructure to do so, the steering committee and scientific advisory board decided to omit the planned interim analysis and to replace it by the final analysis (amendment III, 20 March 2018). This decision took place without knowledge of the data collected at this point and enrolment continued until December 2019.

The adapted sample size of 215 participants was expected to be sufficient to demonstrate non-inferiority with non-inferiority margin of 10% and a power of 80% (one-sided $\alpha = 0.05$, 2.5% of participants reaching the endpoint).[2] We, therefore, adapted a 10% non-inferiority margin, which is in good accordance with suggestions by CPMP/EWP/558/95 rev 3 for non-inferiority trials of infection-site-specific indications, such as acute bacterial skin and skin structure infections.[3]

All decisions regarding sample size and the choice of the non-inferiority margin were reported in the manuscripts describing the trial protocol.[2,4]

**Selection of trial population**

SAB stands out among severe bacterial infections as it is a notoriously difficult-to-treat infection associated with the risk of serious complications and a 90-day mortality of approximately 25% [5]. In the SABATO trial, we used stringent in- and exclusion criteria to focus on patients with a low-risk for SAB-related complications.

Patients entering the study will have already received five to seven days of appropriate intravenous antimicrobial therapy and have no signs and symptoms of complicated S. aureus infection prior to enrolment. Patients with a higher a- priori risk for SAB-related complications are excluded (e.g. severe immunosuppression), or additional diagnostic steps to rule out deep-seated infection are required as defined below.

SAB is more frequent in male patients (m:f = 2:1). The reason for this phenomenon is not known. However, since most infections arise from the skin and nasal flora of the patient, this may reflect a higher nasal colonization rate in men. However, mortality does not vary between male and female gender. Gender-specific differences in efficacy and safety of the used antimicrobial therapy are not expected.

**Inclusion criteria**

The following list includes all changes from protocol amendments

- Age at least 18 years
- Not legally incapacitated
- Written informed consent from the trial subject has been obtained
- Blood culture positive for S. aureus not considered to represent contamination
- At least one negative follow-up blood culture obtained within 24-96 hours (changed in Amendment 1 from 72 hours to 96 hours) after the start of appropriate antimicrobial therapy to rule out persistent bacteremia and absence of a blood culture positive for *S. aureus* at the same time or thereafter.
- Five to seven full days of appropriate intravenous antimicrobial therapy administered prior to randomization documented in the patient chart. Appropriate therapy has all of the following characteristics:
    - Antimicrobial therapy has to be initiated within 72h after the first positive blood culture was drawn.
    - Provided in-vitro susceptibility and adequate dosing (as judged by the principle investigator) the following parenteral antimicrobials are allowed:
        - MSSA: penicillinase-resistant penicillins (e.g. flucloxacillin, cloxacillin), β-lactam plus β-lactamase-inhibitors (e.g. ampicillin+sulbactam, piperacillin+tazobactam), cephalosporins (except ceftazidime), carbapenems, clindamycin, fluoroquinolones, trimethoprim-sulfamethoxazole, doxycycline, tigecycline, vancomycin, teicoplanin, telavancin, linezolid, daptomycin, ceftaroline, ceftobiprole, and macrolides.
        - MRSA: vancomycin, teicoplanin, telavancin, fluoroquinolones, clindamycin, trimethoprim-sulfamethoxazole, doxycycline, tigecycline, linezolid, daptomycin, macrolides, ceftaroline, and ceftobiprole.

**Exclusion criteria**

The following list includes all changes from protocol amendments

- Polymicrobial bloodstream infection, defined as isolation of pathogens other than S. aureus from a blood culture obtained in the time from two days prior to the first positive blood culture with S. aureus until randomization. Common skin contaminants (coagulase-negative staphylococci, diphteroids, Bacillus spp., and Propionibacterium spp.) detected in one of several blood cultures will not be considered to represent polymicrobial infection
- Recent history (within 3 months) of prior S. aureus bloodstream infection
- Contraindications in reference documents or in vitro resistance of S. aureus for all oral or all i.v. study drugs (i.e. there is no therapeutic option available in each study arm)
- Previously planned treatment with active drug against S. aureus during intervention phase (e.g., cotrimoxazole prophylaxis)
- Signs and symptoms of complicated SAB as judged by an ID physician. Complicated infection is defined as at least one of the following:
    - deep-seated focus: e.g. endocarditis, pneumonia, undrained abscess, empyema, and osteomyelitis
    - septic shock, as defined by the AACP criteria, within 4 days before randomization
    - prolonged bacteraemia: positive follow-up blood culture more than 72h after the start of adequate antimicrobial therapy
    - body temperature >38 °C on two separate days within 48h before randomization
- Presence of the following non-removable foreign bodies (if not removed 2 days or more before randomization):
    - prosthetic heart valve
    - deep-seated vascular graft with foreign material (e.g. PTFE or dacron graft). Hemodialysis shunts are not considered deep-seated vascular grafts (s. below).
    - ventriculo-atrial shunt
- Presence of a prosthetic joint (if not removed 2 days or more before randomization). However, this is NOT an exclusion criterion, if all of the following conditions are fulfilled:
    - prosthetic joint was implanted at least 6 months prior, and
    - catheter-related infection, skin and soft tissue infection, or surgical wound infection is present (as defined below), and
    - joint infection unlikely (no clinical or imaging signs)
- Presence of a pacemaker or an automated implantable cardioverter defibrillator (AICD) device (if not removed 2 days or more before randomization). However, this is NOT an exclusion criterion, if all of the following conditions are fulfilled:
    - pacemaker or AICD was implanted at least 6 months prior, and
    - catheter-related infection, skin and soft tissue infection, or surgical wound infection is present (as defined below), and
    - no clinical signs of infective endocarditis, and
    - infective endocarditis unlikely by echocardiography (preferably TEE), and
    - pocket infection unlikely (no clinical or imaging signs)
- Failure to remove within 4 days of the first positive blood culture any intravascular catheter which was present when first positive blood culture was drawn
- Severe liver disease. However, this is NOT an exclusion criterion, if the following condition is fulfilled:
    - catheter-related infection, skin and soft tissue infection, or surgical wound infection is present (as defined below)
- End-stage renal disease. However, this is NOT an exclusion criterion, if all of the following conditions are fulfilled:
    - catheter-related infection, skin and soft tissue infection, or surgical wound infection is present (as defined below), and
    - no clinical signs of infective endocarditis, and
    - infective endocarditis unlikely by echocardiography (preferably TEE), and

    - in patients with a hemodialysis shunt with a non-removable foreign body (e.g. synthetic PTFE loop): no clinical signs of a shunt infection
- Severe immunodeficiency
    - primary immunodeficiency disorders
    - neutropenia (<500 neutrophils/µl) at randomization or neutropenia expected during intervention phase due to immunosuppressive treatment
    - uncontrolled disease in HIV-positive patients
    - high-dose steroid therapy (>1 mg/kg prednisone or equivalent doses given for >4 weeks or planned during intervention)
    - immunosuppressive combination therapy with two or more drugs with different mode of action
    - hematopoietic stem cell transplantation within the past 6 months or planned during treatment period
    - solid organ transplant
    - treatment with biologicals within the previous year
- Life expectancy < 3 months
- Inability to take oral drugs
- Injection drug user
- Expected low compliance with drug regimen
- Participation in other interventional trials within the previous three months or ongoing
- Pregnant women and nursing mothers
- For premenopausal women: failure to use highly-effective contraceptive methods for 1 month after receiving study drug. The following contraceptive methods with a Pearl Index lower than 1% are regarded as highly-effective:
    - oral hormonal contraception ('pill')
    - dermal hormonal contraception
    - vaginal hormonal contraception (e.g., NuvaRing®)
    - contraceptive plaster
    - long-acting injectable contraceptives
    - implants that release progesterone (e.g., Implanon®)
    - tubal ligation (female sterilisation)
    - intrauterine devices that release hormones (hormone spiral)
    - double barrier methods
    - This means that the following are not regarded as safe: condom plus spermicide, simple barrier methods (vaginal pessaries, condom, female condoms), copper spirals, the rhythm method, basal temperature method, and the withdrawal method (coitus interruptus). Due to possible interactions and side effects of the study medication (applies to cotrimoxazole, clindamycin, and flucloxacillin), hormonal contraception may not be safe and another highly-effective contraceptive method needs to be employed.
- Persons with any kind of dependency on the investigator or employed by the sponsor or investigator
- Persons held in an institution by legal or official order

**Definitions for use in exclusion criteria**

<u>Catheter-related infection</u>

- The same S. aureus isolate (based on antimicrobial susceptibility testing result) is present in the positive blood culture and in the catheter tip culture, or
- The same S. aureus isolate is present in the positive blood culture and in pus or skin swab from the catheter exit site, or
- Two initial blood cultures positive for S. aureus exhibit a positive differential time to positivity and there is no other plausible source of infection, or

- Clinically strongly expected catheter-related infection: e.g., pus/reddening/pain at exit-site, or shivers during infusion and no other plausible cause of infection.

Skin and soft tissue infection

- S. aureus cultured from wound swab, or
- Clinical signs of skin and soft tissue infection (abscess, thrombophlebitis, furuncle, etc.) and no signs of any other infective focus

Surgical wound infection

- S. aureus cultured from wound swab, or
- Clinical signs of an infected wound and no signs of any other infective focus

**Relevant changes to inclusion and exclusion criteria in Amendment 1**

The following changes were implemented in Amendment 1 to the study protocol and were approved by the leading Ethics Committee on 29 December 2014.

- The time window for the required negative follow-up blood culture was extended to 96h to account for variations in daily clinical practice.
- The presence of the following foreign bodies was not an exclusion criterion any longer but was allowed under certain circumstances (as listed above): prosthetic joint, prosthetic heart valve, pacemaker, automated implantable cardioverter (AICD).
- Severe liver disease and end-stage renal disease were not exclusion criteria any longer but were allowed under certain circumstances (as listed above).

**Outcome Assessment**

All data were obtained at the study visits or through telephone contacts and are based on the assessment of the study physician, participant interviews, laboratory reports, and chart data. At a baseline visit, participant data (physical examination, vital signs, current medication, imaging studies, and laboratory parameters) were collected. An end-of-treatment visit, within two days of the last dose of study drug, served to assess the current medication and outcome measures. Discharged participants were contacted every two to three days by telephone during study drug administration. Two follow-up visits to assess outcome measures were either performed in-person or by telephone 25 to 39 days and 85 to 99 days after start of study drug. Participants were asked to contact the study staff at any time to report adverse events.

**Definitions used in outcome assessment**

Relapsing SAB and deep-seated infection

Relapsing SAB is defined as positive blood culture for *S. aureus* within the intervention or follow-up period. Deep-seated infection is any deep-seated focus of S. aureus infection resulting from hematogenous dissemination. Diagnosis requires either a positive culture from the respective site, or a blood culture positive with *S. aureus* plus imaging studies showing the presumed focus.

Deep-seated foci consist of, but are not limited to:

- Infective endocarditis, judged by modified Duke criteria
- Vertebral and non-vertebral osteomyelitis
- Suppurative arthritis
- Spinal empyema
- Muscle abscess (e.g. psoas abscess)
- Meningitis, brain abscess
- Lung abscess
- Visceral abscess (kidney, liver, spleen, etc.)

During the follow-up phase, blood cultures are taken when a bloodstream infection is clinically suspected, according to standard of care at the local study site. New onset catheter-related infections as well as superficial skin and soft-tissue or wound infections do not qualify as "deep-seated", since they are likely to result from a new infection. *S. aureus* bacteraemia during the follow-up period is not considered relapsing SAB, if a catheter infection is considered the source of infection with no other plausible source of infection.

To qualify for a "microbiologically confirmed" deep-seated infection or relapsing SAB, the *S. aureus* isolate needed to exhibit the same characteristics as the original infecting isolate, based on antimicrobial susceptibility and genotyping tests as appropriate, and judged not to represent a contaminant by the local investigator. If a suspected deep-seated infection had no corresponding bacterial isolate, it is classified as "clinically suspected".

Attributable death

Death was attributed to SAB when at least one of the following conditions was present:

- positive blood culture for *S. aureus* drawn within 72h before death
- persistent focus of deep-seated *S. aureus* infection at time of death
- persistent signs and symptoms of systemic infection at time of death as judged by study physician
- post-mortem analysis proving *S. aureus* related complication as cause of death.

All other deaths were classified as unrelated to SAB.

**Analysis populations**

Data were analyzed for three study populations: the intention-to-treat (ITT, termed ITT-1 in the statistical analysis plan) population, the modified intention-to-treat population (mITT, termed ITT-2 in the statistical analysis plan), and the clinically evaluable population (CE, also termed per protocol population or PP in the statistical analysis plan).

The intention-to-treat population and the modified intention-to-treat population were designed to explore the upper boundary of the treatment effect. This was done by (1) including all randomized participants in the ITT, regardless whether participants have received study medication or whether they belong to the low-risk group; and (2) by regarding indeterminate or missing outcome data as failures.

The mITT population reflected participants whose data were suitable to answer the trial question at the time when study drug was started. This means, it comprised participants who were randomized, received any study drug, did not withdraw informed consent AND who belonged to the low-risk group according to the inclusion criteria, judging from the data available at start of treatment. The latter excluded participants who had already reached the primary endpoint before the start of study drug but received study drug nonetheless (e.g. a persistently positive blood-culture with *S. aureus*) or who had a high likelihood of complicated infection due to risk factors (e.g. a remaining intravascular graft). As the ITT population, the mITT population was analyzed by regarding indeterminate or missing values as failures.

The clinically-evaluable (CE) population included participants whose data were suitable to answer the trial question at the end of follow-up. It included all participants who received study drug according to protocol, were observed until the end-of-follow-up or reached the primary endpoint, and did not receive antimicrobial therapy during follow-up, if this treatment could have masked the primary endpoint. Study drug was "according to the protocol", when the total duration of antimicrobial therapy (pre-study medication plus study medication) was between 12 and 16 days with at least 5 days of initial intravenous therapy. The Clinical Review Committee carefully evaluated treatment during the intervention phase and follow-up.

From the statistical analysis plan to the manuscript, we renamed the per protocol population and now call it clinical evaluable population. The reason for this name change was the realization, that the name “per protocol” was mostly taken as “study drug was applied as specified in the protocol”. With the name “clinical evaluable”, we wanted to emphasize that further criteria were employed to form a meaningful cohort.

**Primary analysis population**

The statistical analysis plan defined the clinically-evaluable population as the primary analysis population with equal importance to the intention-to-treat analysis, based on EMA guidance from 15 Dec 2012 for non-inferiority trials.[6,7] However, the updated version of the guideline CPMP/EWP/558/95 that came into effect on 19 May 2022 after completion of the SABATO trial states: “In trials that have a clinical primary endpoint, the primary analysis should be conducted in the all randomised (ITT) population.”[3]

The different study populations serve different purposes.[8] The intention-to-treat population follows a pragmatic approach and fully benefits from the randomized assignment of participants. It further reflects the upper-bound for the treatment differences, with the danger, that missing data (which is considered a failure) and treatment outside the protocol may mask treatment effects. On the other hand, the clinically-evaluable population may suffer from imbalances between groups. It more specifically addresses the question whether outcomes differ between groups, if the oral switch is performed correctly and participants do not receive antimicrobials outside the protocol that potentially interfere with outcome. Thus, we consider both analyses complementary and therefore decided to report primarily intention-to-treat data, but also provide data for the clinically evaluable population in the tables of the manuscript. Data of all analyses are included in the supplementary material.

#### Section 3: Results

##### Supplementary Figure S1 (Subgroup analyses)

**Figure S1:** Forest plot of subgroups in the intention-to-treat population. The treatment-control differences and 95%-confidence intervals are shown for the primary endpoint (SAB-related complications within 90-days). The non-inferiority boundary at 10% is denoted by a dashed vertical line. Prespecified subgroups with at least 10 events were formed according to baseline data of age, sex, methicillin-susceptibility, focus of infection, specific comorbidities (pacemaker, prosthetic joint, moderate or severe liver disease, end-stage renal disease, and immunosuppression), Charlson Comorbidity Index (CCI), country, and whether any echocardiography was performed. MSSA, methicillin-susceptible *Staphylococcus aureus*; MRSA, methicillin-resistent *Staphylococcus aureus*; PVC, peripheral venous catheter; CVC, central venous catheter; SSTI, skin and soft-tissue infection.

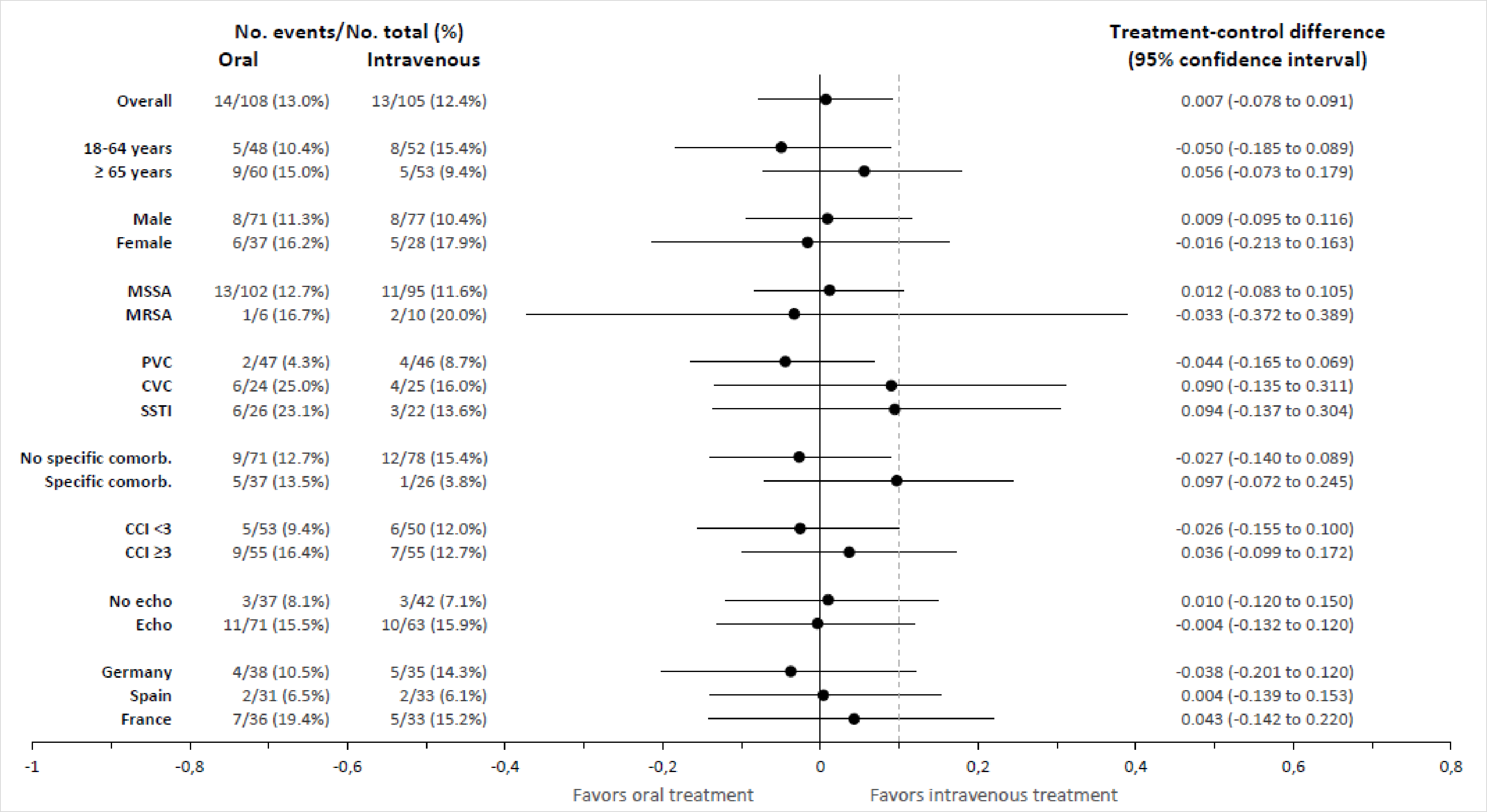
No. events/No. total (%)
Oral
Intravenous
Treatment-control difference
(95% confidence interval)
Overall 14/108 (13.0%) 13/105 (12.4%) 0.007 (-0.078 to 0.091)
18-64 years 5/48 (10.4%) 8/52 (15.4%) -0.050 (-0.185 to 0.089)
≥ 65 years 9/60 (15.0%) 5/53 (9.4%) 0.056 (-0.073 to 0.179)
Male 8/71 (11.3%) 8/77 (10.4%) 0.009 (-0.095 to 0.116)
Female 6/37 (16.2%) 5/28 (17.9%) -0.016 (-0.213 to 0.163)
MSSA 13/102 (12.7%) 11/95 (11.6%) 0.012 (-0.083 to 0.105)
MRSA 1/6 (16.7%) 2/10 (20.0%) -0.033 (-0.372 to 0.389)
PVC 2/47 (4.3%) 4/46 (8.7%) -0.044 (-0.165 to 0.069)
CVC 6/24 (25.0%) 4/25 (16.0%) 0.090 (-0.135 to 0.311)
SSTI 6/26 (23.1%) 3/22 (13.6%) 0.094 (-0.137 to 0.304)
No specific comorb. 9/71 (12.7%) 12/78 (15.4%) -0.027 (-0.140 to 0.089)
Specific comorb. 5/37 (13.5%) 1/26 (3.8%) 0.097 (-0.072 to 0.245)
CCI <3 5/53 (9.4%) 6/50 (12.0%) -0.026 (-0.155 to 0.100)
CCI ≥3 9/55 (16.4%) 7/55 (12.7%) 0.036 (-0.099 to 0.172)
No echo 3/37 (8.1%) 3/42 (7.1%) 0.010 (-0.120 to 0.150)
Echo 11/71 (15.5%) 10/63 (15.9%) -0.004 (-0.132 to 0.120)
Germany 4/38 (10.5%) 5/35 (14.3%) -0.038 (-0.201 to 0.120)
Spain 2/31 (6.5%) 2/33 (6.1%) 0.004 (-0.139 to 0.153)
France 7/36 (19.4%) 5/33 (15.2%) 0.043 (-0.142 to 0.220)
-1
-0,8
-0,6
-0,4
-0,2
0
0,2
0,4
0,6
0,8
Favors oral treatment
Favors intravenous treatment

**Supplementary Figure S2 (Sensitivity analyses)**

Sensitivity analysis with different definitions for alternative endpoints in a Forest plot, showing estimands for the treatment-control difference and 95% confidence intervals as calculated by Zhao's test. In the graph, the non-inferiority boundaries at 5% and 10% are denoted by vertical lines. The following definitions were applied: (I) main analysis: the primary composite endpoint, SAB-related complications within 90 days, was analyzed as planned with missing outcomes counted as failures in the ITT and mITT populations. Missing outcomes were disregarded in the CE population; (II) missing outcomes disregarded: missing outcomes were not considered to be failures; (III) complications after more than 7 days: only SAB-related complications occurring more than 7 days after the start of study drug were counted as failures and missing outcomes were disregarded. Thus, the analysis excludes complications that could have been identified with a more stringent workup at start of study drug; (IV) microbiologically proven: only SAB-related complications that were confirmed with a microbiological result were counted as events. Missing outcomes were disregarded; (V) non-attributable mortality included: the primary endpoint was modified to include all-cause mortality. ITT, intention-to-treat population; mITT, modified intention-to-treat population; CE, clinically evaluable population.

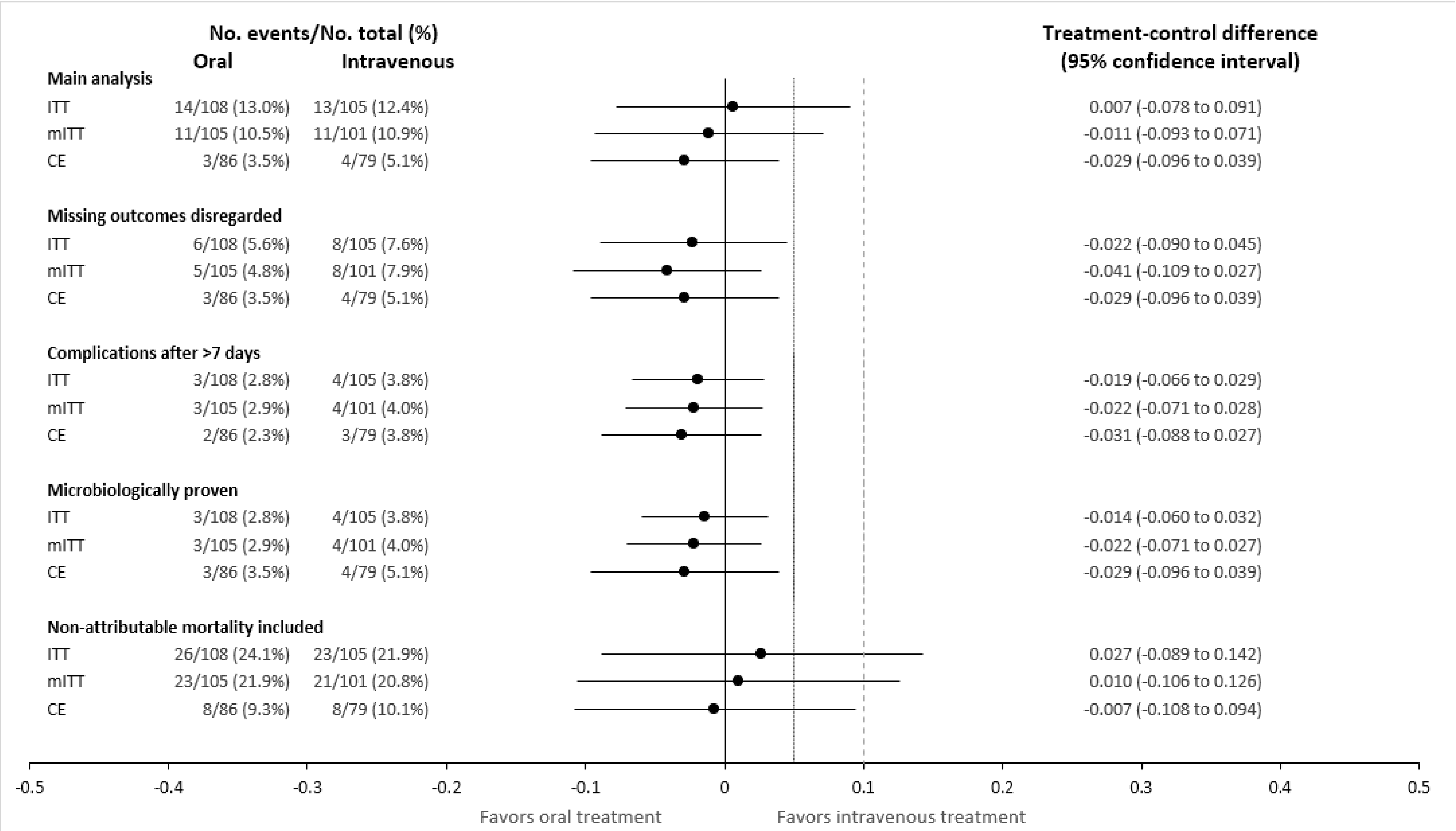
No. events/No. total (%)
Oral
Intravenous
Treatment-control difference
(95% confidence interval)
Main analysis
ITT 14/108 (13.0%) 13/105 (12.4%) 0.007 (-0.078 to 0.091)
mITT 11/105 (10.5%) 11/101 (10.9%) -0.011 (-0.093 to 0.071)
CE 3/86 (3.5%) 4/79 (5.1%) -0.029 (-0.096 to 0.039)
Missing outcomes disregarded
ITT 6/108 (5.6%) 8/105 (7.6%) -0.022 (-0.090 to 0.045)
mITT 5/105 (4.8%) 8/101 (7.9%) -0.041 (-0.109 to 0.027)
CE 3/86 (3.5%) 4/79 (5.1%) -0.029 (-0.096 to 0.039)
Complications after >7 days
ITT 3/108 (2.8%) 4/105 (3.8%) -0.019 (-0.066 to 0.029)
mITT 3/105 (2.9%) 4/101 (4.0%) -0.022 (-0.071 to 0.028)
CE 2/86 (2.3%) 3/79 (3.8%) -0.031 (-0.088 to 0.027)
Microbiologically proven
ITT 3/108 (2.8%) 4/105 (3.8%) -0.014 (-0.060 to 0.032)
mITT 3/105 (2.9%) 4/101 (4.0%) -0.022 (-0.071 to 0.027)
CE 3/86 (3.5%) 4/79 (5.1%) -0.029 (-0.096 to 0.039)
Non-attributable mortality included
ITT 26/108 (24.1%) 23/105 (21.9%) 0.027 (-0.089 to 0.142)
mITT 23/105 (21.9%) 21/101 (20.8%) 0.010 (-0.106 to 0.126)
CE 8/86 (9.3%) 8/79 (10.1%) -0.007 (-0.108 to 0.094)
-0.5
-0.4
-0.3
-0.2
-0.1
0
0.1
0.2
0.3
0.4
0.5
Favors oral treatment
Favors intravenous treatment

**Supplementary Figure S3 (Attributability of Death)**

Stacked probability plot of the cumulative incidence functions, as calculated from the Fine and Gray model. Data from the intention-to-treat population are shown for the oral switch group (E) and the intravenous standard therapy group (F). All-cause mortality was higher in the oral switch group, despite not statistically significant (see **Figure 1 and Table 2**). Deaths were classified as non-evaluable when there was insufficient information regarding the cause of death. Three of four non-evaluable deaths occurred within two weeks after SAB onset. The two attributable deaths occurred around 30 days after SAB onset.

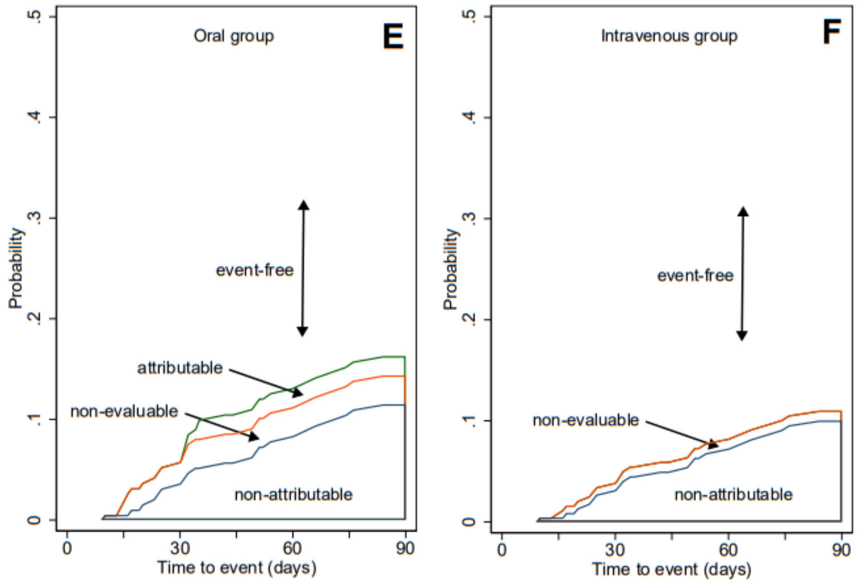

**Supplementary Table S1 (Sites and Enrolment)**
Country, Principal Investigator and number of participants enrolled per site.

| Country | Site | Principal Investigator | Participants enrolled |
|---|---|---|---|
| Germany | Cologne | Gerd Fätkenheuer/Achim Kaasch | 22 |
| Germany | Freiburg | Winfried Kern | 15 |
| Germany | Düsseldorf | Achim Kaasch | 8 |
| Germany | Krefeld | Katrin Kösters | 8 |
| Germany | Jena | Mathias Pletz | 6 |
| Germany | Hannover | Tobias Welte | 4 |
| Germany | Leverkusen | Stefan Reuter | 4 |
| Germany | Berlin | Keikawus Arasteh | 3 |
| Germany | Frankfurt | Christoph Stephan | 2 |
| Germany | Lübeck | Jan Rupp | 1 |
| Spain | Sevilla | Jesus Rodríguez-Baño | 29 |
| Spain | Sevilla | Jose-Miguel Cisneros | 24 |
| Spain | Barcelona | Alex Soriano | 7 |
| Spain | Palma de Mallorca | Luisa Martin | 4 |
| The Netherlands | Breda | Jan Kluytmans | 4 |
| The Netherlands | Utrecht | Marc Bonten | 2 |
| The Netherlands | Utrecht | Anneloes Vlek | 1 |
| France | Paris | Raphael Lepeule | 13 |
| France | Tours | Louis Bernard | 12 |
| France | Paris | Bruno Fantin | 6 |
| France | Paris | Jean Michel Molina | 6 |
| France | Quimper | Jean-Philippe Talarmin | 6 |
| France | La Roche sur Yon | Marine Morrier | 5 |
| France | Nantes | David Boutoille | 5 |
| France | Paris | Elisabeth Rouveix Nordon | 5 |
| France | Chambéry | Emmanuel Forestier | 4 |
| France | Annecy | Virginie Vitrat | 3 |
| France | Grenoble | Jean Paul Stahl | 1 |
| France | Orléans | Laurent Hocqueloux | 1 |
| France | Rennes | Pierre Tattevin | 1 |
| France | St. Etienne | Frédéric Lucht | 1 |

**Supplementary Table S2 (Admission wards)**
Admission ward of patients with SAB in routine care. Patients with SAB were retrospectively assessed for potential eligibility based on routine data collected by an infectious diseases or microbiology service. These routine data were anonymized and documented in a separate database.

| Specialty | N=5,063 | Percentage |
|---|---|---|
| Internal medicine (other than subspecialties below) | 1,273 | 25.1% |
| Intensive care unit | 351 | 6.9% |
| Nephrology | 345 | 6.8% |
| Haematology/oncology | 342 | 6.8% |
| Cardiothoracic surgery | 342 | 6.8% |
| Orthopaedic surgery | 278 | 5.5% |
| Abdominal surgery | 228 | 4.5% |
| Surgery (other) | 225 | 4.4% |
| Neurology | 193 | 3.8% |
| Neurosurgery | 191 | 3.8% |
| Cardiology | 171 | 3.4% |
| Infectious diseases | 146 | 2.9% |
| Emergency medicine | 114 | 2.3% |
| Dermatology | 110 | 2.2% |
| Gastroenterology | 105 | 2.1% |
| Other | 248 | 4.9% |
| Not recorded | 401 | 7.9% |

**Supplementary Table S3 (Screening Population)**
Characteristics and reason for exclusion of screened patients as collected in routine healthcare**.** Data were recorded until exclusion was determined.

| **Characteristic** | **Count (percentage)** | **Data available (N)** |
|---|---|---|
| Male sex | 3,340 (66.0%) | 5,063 |
| Age in years; median (IQR) | 68 (55-77) | 5,063 |
| Age < 18 years | 12 (0.2%) | 5,063 |
| Legally incapacitated | 280 (6.5%) | 4,302 |
| Blood culture judged as contamination | 64 (1.7%) | 3,702 |
| Negative follow-up blood culture at 48 to 72 hours (96 hours*) after start of adequate therapy not available | 1,068 (34.8%) | 3,072 |
| 5-7 days of appropriate intravenous antimicrobial therapy prior to randomization not documented | 319 (11%) | 2,901 |
| Polymicrobial bloodstream infection | 315 (8.6%) | 3,677 |
| Prior *S. aureus* bloodstream infection within 3 months | 104 (3%) | 3,461 |
| In vitro resistance to all oral study medications | 7 (0.2%) | 3,483 |
| Contraindications for all oral or all intravenous study medications | 35 (1%) | 3,401 |
| Previously planned treatment with drug active against *S. aureus* during intervention or follow-up phase (e.g. cotrimoxazole prophylaxis) | 109 (3.3%) | 3,053 |
| Signs and symptoms of complicated *S. aureus* bloodstream infection present | 1,692 (52.4%) | 3,232 |
| - Deep seated focus present (e.g. endocarditis, pneumonia, undrained abscess, empyema, and osteomyelitis) | 1,589 (47.5%) | 3,342 |
| - Septic shock within 4 days before randomization | 425 (14.8%) | 2,875 |
| - Prolonged bacteraemia (positive follow-up blood culture more than 72 hours after start of adequate antimicrobial therapy) | 560 (20.0%) | 2,797 |
| - Body temperature >38 °C on two separate days within 48h before randomization | 124 (4.5%) | 2,775 |
| Presence of non-removable foreign body (prosthetic heart valves, deep-seated vascular grafts with foreign material, ventriculo-atrial shunt); haemodialysis shunts are accepted | 847 (26.4%) | 3,207 |
| Intravascular catheter not removed within four days of the first positive blood culture | 169 (5.9%) | 2,883 |
| Severe liver disease† unless the following condition were fulfilled: catheter-related infection, skin and soft tissue infection, or surgical wound infection is present | 178 (5.9%) | 2,992 |
| End-stage renal disease† unless the following conditions were fulfilled: - catheter-related infection, skin and soft tissue infection, or surgical wound infection is present, and - no clinical signs of infective endocarditis, and - infective endocarditis unlikely by echocardiography (preferably TEE), and - in patients with a haemodialysis shunt with a non-removable foreign body (e.g., synthetic PTFE loop): no clinical signs of shunt infection | 258 (8.5%) | 3,052 |
| Severe immunodeficiency | 477 (15.5%) | 3,080 |
| 'Do not resuscitate" order or life expectancy below 3 months | 514 (16.7%) | 3,075 |
| Inability to take oral medication | 265 (8.9%) | 2,984 |
| Injection drug user | 110 (3.7%) | 2,942 |
| Expected low compliance with study medication | 225 (7.7%) | 2.928 |
| Participation in other interventional trials | 56 (1.9%) | 2,876 |
| Pregnant women and nursing mothers | 16 (1.7%) | 959 |
| Failing willingness to use highly effective contraceptives | 3 (0.3%) | 928 |
| Dependency on investigator | 1 (0.04%) | 2,852 |
| Persons held in an institution by legal or official order | 10 (0.4%) | 2,844 |

* With Amendment 2, the 72 hours were extended to 96 hours.
† Until Amendment 2, severe liver disease and end-stage renal disease were an exclusion criterion.

**Supplementary Table S4 (Antimicrobial Therapy)**

Characteristics of antimicrobial medication, active against the particular strain of *Staphylococcus aureus*, during the entire course of the study for the intention-to-treat and clinically evaluable populations. Qualitative data are reported as numbers (%), quantitative data as median (interquartile range).

| | **Intention-to-treat** | | **Clinically evaluable** | |
|---|---|---|---|---|
| | **Oral (n=108)** | **Intravenous (n=105)** | **Oral (n=86)** | **Intravenous (n=79)** |
| Duration of pre-study medication – days | | | | |
| Median | 6 | 6 | 7 | 6 |
| Inter-quartile range | 6-7 | 5-7 | 6-7 | 5-7 |
| Duration of study medication – days | | | | |
| Median | 8 | 8 | 8 | 8 |
| Inter-quartile range | 7-9 | 7-9 | 7-9 | 7-9 |
| Total duration of antimicrobial therapy for SAB – days* | | | | |
| Median | 14 | 14 | 14 | 14 |
| Inter-quartile range | 14-15 | 14-15 | 14-15 | 14-15 |
| Participants receiving >7 days of antimicrobials during follow-up phase – no (%)† | 14 (13.0%) | 17 (16.2%) | 6 (7.0%) | 12 (15.2%) |
| Initial intravenous study medication | | | | |
| Cefazolin i.v. | 0 | 46 (43.8%) | 0 | 37 (46.8%) |
| Cloxacillin i.v. | 0 | 16 (15.2%) | 0 | 13 (16.5%) |
| Flucloxacillin i.v. | 0 | 29 (27.6%) | 0 | 23 (29.1%) |
| Vancomycin i.v. | 0 | 7 (6.7%) | 0 | 3 (3.8%) |
| Daptomycin i.v. | 0 | 5 (4.8%) | 0 | 3 (3.8%) |
| Initial oral study medication | | | | |
| Cotrimoxazole p.o. | 63 (58.3%) | 0 | 57 (66%) | 0 |
| Clindamycin p.o. | 35 (32.4%) | 0 | 23 (27%) | 0 |
| Linezolid p.o. ‡ | 9 (8.3%) | 0 | 6 (7%) | 0 |
| Switch to other study medication | 4 (3.8%) | 5 (4.8%) | 4 (5%) | 4 (5.1%) |
| Not receiving study medication | 1 (0.9%) | 2 (1.9%) | 0 | 0 |

* allowing for a maximum of two days of interrupted therapy

† The duration of follow-up phase antimicrobials was counted starting from the planned end of the treatment course (including a 15% variation for the duration, i.e., overall 16 days), until the end of follow-up.

‡ Five participants received oral linezolid for methicillin-susceptible SAB instead of trimethoprim-sulfamethoxazole or clindamycin. These participants were accepted for CE analysis by the Clinical Review Committee.

**Supplementary Table S5 (SAB-related complications)**

Individual listing of SAB-related complications. Data are ordered according to treatment arm, analysis population, and number of days from start of study medication to the time when the complication occurred. Complications were classified as early deep-seated infections when they occurred within 7 days after start of study medication and were thus likely to have been undetected at randomization. Late complications were defined as complications occurring more than 7 days after the start of study medication. In participants that did not receive study medication, the day of randomization was used for calculating the time to event. Complications were labelled “microbiologically documented”, when *S. aureus* was cultured from a participant’s specimen (including blood culture). One additional participant in the oral treatment group (data not shown) had a new-onset hemodialysis-catheter-related infection 43 days after randomization, which was classified as “unrelated to SAB” according to the study protocol. CCI, Charlson comorbidity index; CE, clinically evaluable; CT, computed tomography; ITT, intention-to-treat; mITT, modified intention to-treat; OST, oral switch therapy; IST, intravenous standard therapy; EOT, end of treatment; PICC, peripherally inserted central venous catheter; SAB, *S. aureus* bloodstream infection.

| Arm | Analysis | Age group and sex | Admission diagnosis | CCI | Initial focus of SAB | Days to complication | Reported complication | Death | Microbiologically documented | Interpretation |
|---|---|---|---|---|---|---|---|---|---|---|
| **OST** | CE mITT ITT | 60-69, f | Oedema of lower limbs | 5 | Central venous catheter | 1 | Readmission on day 2 of study medication due to clinically suspected complicated infection with pulmonary focus on chest CT | No | No | Early deep-seated infection |
| **OST** | CE mITT ITT | 70-79, m | Hepatic lobectomy due to colorectal cancer metastasis | 7 | Central venous catheter | 19 | Septic knee arthritis and SAB followed by aortic dissection (Gram-positive cocci in pathology specimen) | Yes | Yes | Late complication (deep-seated focus, bacteremia, attributable death) |
| **OST** | CE mITT ITT | 60-69, m | Cardiac failure | 4 | Peripheral venous catheter | 28 | Participant had a second episode of SAB with *S. aureus* cultured from blood and a tibial ulcer | No | Yes | Late complication (bacteremia, deep-seated focus) |
| **OST** | mITT ITT | 80-89, m | Cellulitis | 4 | Skin and soft tissue infection | 4 | Participant with diabetic foot ulcer, a CT was performed on day 5 of study medication and showed osteomyelitis at the site of the ulcer. | No | No | Early deep-seated infection |
| **OST** | mITT ITT | 80-89, f | Hypertensive crisis | 2 | Peripheral venous catheter | 15 | Participant felt weak 3d after EOT but declined readmission. On day 8 after EOT, participant was found unconscious at home and was readmitted. Recurrent SAB due to suppurative thrombophlebitis at exit site of previous catheter. TEE unremarkable. Death one week later. | Yes | Yes | Late complication (extension of focus, bacteremia, attributable death) |
| **OST** | ITT | 50-59, f | Repeated falls | 5 | Skin and soft tissue infection (subcutaneous abscess) | 3 | On day 3 of study medication, an extension of the original focus occurred from the gluteal region to proximal inner thigh. Resolved with drainage and prolongation of oral antimicrobial therapy. | No | No | Early deep-seated infection (extension of focus) |

| | | | | | | | | | | |
|---|---|---|---|---|---|---|---|---|---|---|
| **IST** | CE mITT ITT | 70-79, m | Pancreatic adenocarcinoma | 7 | Central venous catheter | 1 | Septic thrombosis at the site of the central venous catheter, apparent on day 2 of study medication | No | No | Early deep-seated infection |
| **IST** | CE mITT ITT | 50-59, f | Abscess at peripheral venous catheter exit site from previous hospitalization | 3 | Peripheral venous catheter | 14 | Aortic valve endocarditis, which was not visible on TEE on the day before study medication. Repeat TEE on day 5 after EOT showed a vegetation that subsequently diminished in size with intravenous antimicrobial treatment. | No | No | Late complication (endocarditis) |
| **IST** | CE mITT ITT | 80-89, m | Syncope | 3 | Peripheral venous catheter | 53 | Readmission with vertebral osteomyelitis and SAB | No | Yes | Late complication (metastatic seeding, bacteremia) |
| **IST** | CE mITT ITT | 20-29, f | Sciatica | 0 | Urinary tract infection | 69 | Readmission with an iliac abscess and SAB | no | Yes | Late complication (metastatic seeding, bacteremia) |
| **IST** | mITT ITT | 40-49, m | Supratentorial intraventricular bleeding | 0 | PICC line | 4 | Multiple pulmonary and brain foci consistent with endocarditis. Participant declined echocardiography | No | No | Early deep-seated infection (presumed endocarditis) |
| **IST** | mITT ITT | 70-79, m | Hypopharynx carcinoma | 4 | PICC line | 6 | SAB from endogenous PICC line infection after line change; classified as deep-seated by CRC | No | Yes | Early deep-seated infection (bacteremia) |
| **IST** | mITT ITT | 50-59, m | Oedema of the left hemiface | 0 | Skin and soft tissue infection | 7 | Suspected pulmonary emboli in CT scan at EOT | No | No | Early deep-seated infection (presumed endocarditis) |
| **IST** | mITT ITT | 50-59, f | Transfer from other hospital with nosocomial bacteremia | 4 | Central venous catheter | 13 | Recurrent bacteremia due to osteomyelitis of midfoot as evidenced by MRI 8 days after EOT | No | Yes | Late complication (bacteremia) |

**Supplementary Table S6 (SAE leading to death)**

Listings of serious adverse events (SAE) leading to death within 90 days, ordered by study arm and analysis population. CE, clinically evaluable; ITT, intention-to-treat; mITT, modified intention to-treat; OST, oral switch therapy; IST, intravenous standard therapy; CCI, Charlson Comorbidity Index.

| Arm | Analysis set | Age group, sex | Admission diagnosis | CCI | Initial focus of SAB | Survival in days | Reported adverse event leading to death | Death attributable to SAB |
|---|---|---|---|---|---|---|---|---|
| OST | CE mITT ITT | 60-69, m | Chest pain | 3 | Peripheral catheter | 1 | Ischemic heart disease (cardiac arrest due to myocardial infarct) | no |
| OST | CE mITT ITT | 70-79, m | Liver metastectomy | 7 | Central venous catheter | 27 | Aortic dissection | yes |
| OST | CE mITT ITT | 80-89, m | Acute coronary syndrome | 8 | Peripheral catheter | 34 | Aspiration pneumonia | no |
| OST | CE mITT ITT | 70-79, f | Cutaneous eruption | 6 | Skin and soft tissue | 60 | Stage IV melanoma | no |
| OST | CE mITT ITT | 80-89, m | Septic shock | 1 | Skin and soft tissue | 69 | Respiratory infection (probable aspiration pneumonia) | no |
| OST | CE mITT ITT | 50-59, f | Intestinal pseudo-obstruction | 6 | Peripheral catheter | 78 | Progress of metastatic diseases - ovarian cancer stage IV | no |
| OST | mITT ITT | 70-79, m | Parotitis | 7 | Skin and soft tissue | 7 | Death from probable pulmonary embolism | non-evaluable |
| OST | mITT ITT | 80-89, m | Fall | 0 | Skin and soft tissue | 7 | Cardiorespiratory arrest | non-evaluable |
| OST | mITT ITT | 60-69, f | Stroke | 7 | Peripheral catheter | 15 | Left ventricular failure | no |
| OST | mITT ITT | ≥90, f | Progredient cough and increasing loss of appetite | 3 | Suspected broncho-pulmonary focus | 16 | Cardiac arrest | no |
| OST | mITT ITT | 70-79, m | Myocardial infarction | 6 | Central venous catheter | 24 | Cardiac arrest | no |
| OST | mITT ITT | 80-89, f | Hypertensive crisis | 2 | Peripheral catheter | 25 | Sepsis | yes |
| OST | mITT ITT | 60-69, m | Acute kidney injury | 10 | Skin and soft tissue | 30 | Hepatic decompensation in participant with hepatocellular carcinoma | no |
| OST | mITT ITT | 80-89, m | Acute shunt bleeding | 3 | Skin and soft tissue | 45 | Sepsis (blood culture negative; new inoperable urothelial carcinoma with ureter stenosis and recent pyelonephritis) | no |
| OST | mITT ITT | 60-69, f | Cardiac arrest | 2 | Peripheral catheter | 55 | Acute myocardial infarction | no |
| OST | mITT | 60-69, m | Abdominal pain | 7 | Central venous catheter | 60 | Pancreatic cancer with hepatic metastases | no |

| | | | | | | | | |
|---|---|---|---|---|---|---|---|---|
| | ITT | | | | | | | |
| OST | ITT | 80-89, f | Inflammatory syndrome related to pemphigus vulgaris | 1 | Central venous catheter | 10 | Cardiopulmonary arrest (in malnourished participant) | non-evaluable |
| IST | CE<br>mITT<br>ITT | 60-69, m | Decompensated liver cirrhosis | 3 | Central venous catheter | 9 | Hepatic encephalopathy (with liver failure and cardiac arrest) | no |
| IST | CE<br>mITT<br>ITT | 80-89, m | Fever | 8 | Skin and soft tissue | 43 | (Progressive) heart failure | no |
| IST | CE<br>mITT<br>ITT | 30-39, m | Recurrence of testicular cancer | 6 | Central venous catheter | 45 | Community acquired bilateral pneumonia (leading to multiorgan failure) | no |
| IST | CE<br>mITT<br>ITT | 70-79, m | Contrast medium-induced nephrotoxicity | 5 | Central venous catheter | 67 | Diffuse peritonitis secondary to bladder perforation | no |
| IST | mITT<br>ITT | 80-89, m | Digestive bleeding | 4 | Peripheral catheter | 18 | Ischemic hepatitis | no |
| IST | mITT<br>ITT | 60-69, m | Suspected heart attack | 1 | Peripheral catheter | 18 | Cardiogenic shock | no |
| IST | mITT<br>ITT | 80-89, f | Acute renal failure | 7 | Skin and soft tissue | 22 | Septic shock (probable urosepsis) | no |
| IST | mITT<br>ITT | 80-89, f | Mesenteric angina | 2 | Peripheral catheter | 26 | Severe malnutrition; ischemic intestinal failure | non-evaluable |
| IST | mITT<br>ITT | 70-79, m | Acute exacerbation of chronic pancreatitis | 1 | Peripheral catheter | 28 | Respiratory insufficiency (on acute exacerbation of chronic pancreatitis) | no |
| IST | mITT<br>ITT | 80-89, m | Electrical storm caused by Amiodaron medication | 6 | Central venous catheter | 42 | Progressive heart failure (and renal failure) | no |
| IST | mITT<br>ITT | 80-89, m | Cardiac failure | 9 | Central venous catheter | 68 | Cardiac failure | no |

**Supplementary Table S7 (Low-risk Criteria)**

Overview of criteria for low-risk *Staphylococcus aureus* bloodstream infection. For detailed in- and exclusion criteria see Appendix 3. AICD, automated implanted cardioverter defibrillator; SAB, *S. aureus* bloodstream infection.

| Category | Low-risk criteria |
|---|---|
| History | • No prior episode of SAB within preceding 3 months |
| Intravenous pre-treatment | • Five to seven days of appropriate intravenous antimicrobial therapy initiated within 72h after the first positive blood culture was drawn |
| Follow-up blood cultures | • Negative follow-up blood culture within 24-96 hours after the start of appropriate antimicrobial therapy and absence of further positive blood cultures |
| Focus | • No signs and symptoms of a deep-seated focus (endocarditis, pneumonia, undrained abscess, empyema, osteomyelitis etc.) |
| Clinical stability | • Absence of septic shock within four days prior to randomization<br>• Absence of fever (body temperature >38 °C) on two separate days within 48h prior to randomization |
| Intravascular catheter | • Removal within 4 days of any intravascular catheter present at onset |
| Foreign bodies | • Absence of prosthetic heart valve, pacemaker, prosthetic joint, deep-seated vascular graft with foreign material (hemodialysis shunts are accepted), or ventriculo-atrial shunt<br>• Pacemaker/AICD: accepted if<br>○ SAB due to catheter-related infection or skin and soft tissue infection, *and*<br>○ device implanted at least 6 months prior, *and*<br>○ no clinical signs of pocket infection, *and*<br>○ no clinical signs of infective endocarditis, *and*<br>○ echocardiography unremarkable<br>• Prosthetic joint: accepted, if<br>○ SAB due to catheter-related infection or skin and soft tissue infection, *and*<br>○ prosthesis implanted at least 6 months prior, *and*<br>○ no evidence of prosthesis infection (clinical and/or imaging) |
| Absence of severe immuno-deficiency or immunosuppression | • Primary immunodeficiency disorders<br>• Neutropenia at randomization or expected during intervention<br>• High-dose steroid therapy (> 1 mg/kg prednisone for > 4 weeks or during intervention) |

| | |
|---|---|
| | • Immunosuppressive combination therapy with two or more drugs with different mode of action<br>• Haematopoietic stem cell transplantation within the past 6 months or planned during treatment period<br>• Solid organ transplant<br>• Treatment with immunosuppressive biologicals<br>• Uncontrolled disease in HIV-positive patients |
| Absence of major comorbidities | • End-stage renal disease accepted, if<br> ○ SAB due to catheter-related infection or skin and soft tissue infection, *and*<br> ○ no clinical signs of infective endocarditis, *and*<br> ○ echocardiography unremarkable, *and*<br> ○ no clinical signs of a shunt infection in patients with a haemodialysis shunt containing a foreign body (e.g. synthetic PTFE loop)<br>• Severe liver disease accepted, if<br> ○ SAB due to catheter-related infection or skin and soft tissue infection |

#### Data sharing statement

| | |
|---|---|
| **Will individual patient data be available?** | Yes |
| **What data will be shared?** | Individual participants data (including data dictionary) that underlie the results reported in this article after deidentification |
| **What other documents will be available?** | Study protocol, statistical analysis plan, informed consent form |
| **When will data be available?** | Beginning 12 months after publication and ending 36 months following publication |
| **With whom data will be shared?** | Researchers who submit a methodologically sound proposal, which is approved by the Ethics Committee at the institution of the corresponding author |
| **For what type of analysis?** | Any analysis |
| **By what mechanism will data be made available?** | Requests should be directed to the corresponding author of the manuscript. To gain access a data sharing agreement will need to be signed. Data will be available from the University data warehouse. |

#### Frequently asked Questions regarding the SABATO trial

**Trial Rationale**

**Why was this trial needed, can we not deduce from Gram-negative bacteremia or other Gram-positive organisms that oral switch is possible?**

Studies addressing severe infections with other microorganisms cannot be extrapolated to *S. aureus*. Gram-negative bacteremia has different clinical characteristics than SAB. SAB differs from other bloodstream infections insofar as metastatic foci, the relapse rate, and late metastatic complications are more common. This risk posed by *S. aureus* is also considerably higher than in bacteremia with other Gram-positive organisms.

**What is the evidence for early oral switch before the SABATO trial?**

The trial is the first randomized controlled trial that addresses early oral switch in patients with low-risk SAB. When the trial was started there were very few observational data on oral switch therapy in any condition. In the mean-time, observational data has been published on oral switch in *S. aureus* bacteremia. Recent retrospective, observational studies found similar outcomes for oral switch therapy in complicated SAB,[9,10] as well as uncomplicated or low-risk SAB,[11–14] methicillin-resistant *S. aureus* (MRSA) bacteraemia,[15] or any SAB.[16] The drawback of observational studies is the inherent bias by indication. If indication bias is present, one would expect that patients with a better clinical prognosis are selected for oral switch therapy. Indeed, all observational studies showed a lower mortality in the group receiving oral medication, which is highly suggestive for indication bias.

**What was the Evidence from RCTs regarding oral therapy before the SABATO trial?**

Previous RCTs on *S. aureus* infections have included only few patients with bacteremia receiving oral therapy. Circumstantial evidence comes from a recent meta-analysis that assessed randomized controlled trials on oral treatment in severe systemic infections not specific to *S. aureus*. No significant difference for overall treatment success in ten studies on bacteraemia was found, whereas in four studies that assessed endocarditis, early oral therapy was superior.[17] Drugs previously studied were linezolid,[18–20] fleroxacin plus rifampicin,[21] trimethoprim-sulfamethoxazole,[22,23] trimethoprim-sulfamethoxazole plus rifampicin,[24] and several combination regimens for endocarditis.[25] Studies did not report specific information about duration and route of administration in participants with SAB, except for two studies. Schrenzel *et al.* reported cure in 15/19 participants having catheter-related bacteraemia and 10/11 participants having primary bacteraemia with oral fleroxacin-rifampicin versus 10/11 and 4/5 participants with parenteral standard medication.[21] Harbarth *et al.* found clinical cure in 6/9 participants receiving a combination of oral trimethoprim-sulfamethoxazole and rifampicin versus 7/9 participants receiving oral linezolid.[24] In participants with *S. aureus* endocarditis from the POET trial, 3/40 participants with oral medication reached the primary composite outcome (all-cause mortality, unplanned cardiac surgery, embolic events, or relapse of bacteraemia) versus 3/47 participants with standard intravenous medication (odds ratio 0.84, 95%-CI 0.15-4.78).[25]

**Why did you chose to test the hypothesis in low-risk SAB and not e.g. in complicated SAB?**

We defined low-risk SAB according to criteria that we deduced from our large cohort study INSTINCT. Inclusion and exclusion criteria were geared towards a study population, in which we expected that 14 days of antimicrobial therapy were sufficient for successful treatment of SAB. Many conditions of SAB require longer treatment, e.g. 4-6 weeks in endocarditis or up to 12 weeks in osteomyelitis. Furthermore, the clinical course is highly dependent on focus removal, which may not be feasible in conditions with “complicated SAB”. Thus, our inclusion and exclusion criteria selected for a population that mostly consisted of participants with catheter-related infection and skin and soft-tissue infection as source of SAB. This was supposed to yield a patient population that was as homogenous as possible regarding the focus, therapy duration, and the clinical course.

It is important to note that after 5-7 days of iv therapy, low-risk patients are usually stable with regard to SAB, and patients can be discharged, unless another condition precludes discharge.

**Trial Design**

**Why did you NOT chose PET-CT as an inclusion criterion?**

Access to PET-CT was limited in our study sites, especially since PET-CT would have to be performed before oral switch. We did not think that this study would have been feasible because of the small inclusion time window. Additionally, false positive results may potentially lead to exclusion of patients from the trial, since there may not be sufficient time to confirm or reject a positive PET-CT result.

**Why did you NOT chose echocardiography as an inclusion criterion?**

We left echocardiography to the discretion of the treating physicians. Given the stringent inclusion and exclusion criteria, we reasoned that the occurrence of unnoticed endocarditis before starting study drug would be low. This decision was based on our previous work.[26]

**Why does your statistical analysis plan highlight the clinically evaluable population and not the intention-to-treat population?**

The statistical analysis plan defined the per protocol (clinically evaluable) population as the primary analysis population with equal importance to the intention-to-treat analysis, based on EMA guidance documents.[6,7] A recent revision of the guidance document that came into effect after completion of the SABATO trial (19 May 2022) states that the primary analysis should be conducted in the all randomised (ITT) population.[3]

The two study populations serve different purposes and have different strengths and weaknesses.[8] We thus decided to present data from both populations in the main manuscript while discussing primarily ITT data.

**Why were participants lost between the ITT and CE population?**

The CE dataset included all study subjects who received study drug according to protocol, did not receive potentially interfering antimicrobial therapy during follow-up, and reached a defined endpoint in the trial. We assessed “treatment according to protocol” during the intervention AND the 90-day follow-up period. Antimicrobial therapy with study drug was considered as per protocol, when the total duration was between 12 and 16 days with at least 5 days of initial intravenous therapy. Furthermore, we took care to avoid masking of *S. aureus*-related complications by excluding participants from the CE analysis set. We excluded participants who received antimicrobial treatment for more than 3 days during the follow-up period, when treatment was active against *S. aureus* but given for other causes than *S. aureus*. This stringent evaluation led to loss of about 20% of participants between CE and ITT. However, the same loss (from 499 participants in ITT to 410 participants in CE) was seen in a trial on uncomplicated staphylococcal bacteremia.[27]

**Why did you NOT chose all-cause mortality as a primary outcome?**

The choice of a primary endpoint for trials in SAB can be difficult. Our composite primary endpoint included sequelae related to SAB, but did not include mortality unrelated to SAB.

Mortality in SAB is high, about 30% at 90 days. All-cause mortality in SAB is strongly dependent on comorbidities, age, MRSA, the infective foci and age. Especially in low-risk SAB, many patients are hospitalized for other reasons than SAB and they suffer e.g. a catheter-related infection during their hospital stay. Therefore, only part of the observed mortality can be directly attributed towards SAB. Switching to oral medication has the risk that (1) oral antimicrobials are not as effective as iv antimicrobials, e.g. due to lower plasma levels, and (2) oral antimicrobials have a unfavorable risk profile compared to iv antimicrobials. “Less effective” equates to a higher rate of relapse or treatment failures with recurrence of *S. aureus* bacteremia or infective foci, or a higher rate of death

attributable to *S. aureus*, all reflected in our primary endpoint. Regarding the risk profile, we had no indication that oral agents have a worse risk profile than i.v. agents.
In low-risk SAB, we expected a 90-day mortality of 10-15% from our cohort data. The rate of relapse, recurrence and attributable death was estimated at 2.5%. We therefore reasoned, that if we include all-cause mortality, it may be difficult to observe a small difference between the groups. However, 14-day, 30-day, and 90-day mortality are secondary endpoints and we conducted competing risk analysis to assess the contribution of all-cause mortality.

**Why do you disregard participants that died from causes other than SAB from the analysis of the primary endpoint?**
Participants that died from other causes would have varying time until the end-of-FU, if they had survived. Thus, death is a competing event towards the primary endpoint. There are several ways to address the issue. We decided to exclude these participants from the evaluation of the primary endpoint and address competing risk in a secondary analysis.

**Why did you use Zhao's test for hypothesis testing?**
Zhao's test has its strength in clinical trials with small sample sizes. The test inherently stratifies data by center, obviating the need for later stratification.

**How did you ensure that your results are robust?**
First we consider CE and ITT analyses as equivalent for interpretation and both demonstrated non-inferiority. Second, we conducted a series of sensitivity analyses (Figure S2). For example, we showed that findings remained robust, when incorporating non-attributable mortality in the primary endpoint or when only considering failures that occur later than 7 days after start of study drug. Furthermore, results remained robust, when treating non-attributable mortality as competing event.

**Trial conduct**
**From 6,063 assessed patients with SAB, only 215 were enrolled, why so few?**
The ratio of 1:28 is what is expected, when comparing to other RCT on SAB.[28] There are a number of hurdles that have to be overcome when recruiting patients with SAB to a trial. Patients are usually distributed across hospital wards, so access needs to be ensured. Timing is critical since the intervention in the trial is between 5-7 days after the positive blood culture. During this time the patient needs to be diagnosed properly and must be willing to consent. The fact that low-risk SAB is only present in a fraction of patients does impede fast recruitment with a limited number of sites.

**Why did the recruitment of participants take so long?**
A number of reasons caused a long recruitment period, the main reasons were: First, not many study centers were ready for the task to include patients in all wards of the hospital. It took a long time and more funds to add further study centers, especially in France. Second, at least 50% of eligible patients were not willing to participate in this clinical trial; some because they had preferences for either oral or intravenous medication, and others because they did not want to participate in any clinical trial. Third, patients that meet the low-risk criteria have become increasingly rare, at least in the German setting.

**Why was the trial terminated early?**
The main reasons were overestimation of the number of low-risk infections, the willingness of patients to consent, and the logistical hurdles for the study teams to enroll patients across the hospital within a limited time window. This led us to increase the number of study sites and to reduce the sample size by converting the interim analysis to a final analysis. Nevertheless, a 5% non-inferiority margin was met in the CE analysis and results are still sufficiently precise to draw

conclusions. This led the Steering Committee to recommend termination of the trial at 215 participants.

**Interpretation**

**You found an earlier discharge in the oral arm. How do differences in discharge compare to the literature?**

We observed earlier discharge in participants on oral treatment (median length of stay 11 days versus 15 days). Previous studies found even larger differences. Schrenzel *et al.* reported 12 days versus 23 days[21] and Iversen *et al*. reported 3 days versus 19 days in the POET trial.[25] We assume that differences between these studies reflect the patient populations and the fact that low-risk SAB patients are often hospitalised for reasons other than SAB and thus discharge does not depend as much on oral switch than e.g. in infectious endocarditis.

**There were a slightly higher 90-day mortality and more adverse events in the oral arm. Is this relevant?**

We found a higher 90-day mortality in the oral arm, although not statistically significant. In concordance, the rate of adverse events was higher in the oral arm. This points either towards random fluctuations, imbalances between the groups, or an adverse effect of oral study medication.

There were slight imbalances between the groups regarding Charlson comorbidity index, age, and body-mass index. However, we did not find convincing evidence that these imbalances drive mortality. We further cannot exclude that the oral medication influences outcome, although the oral regimens are generally considered as having a favorable safety profile. Nevertheless, future trials should confirm equivalence of oral therapy also regarding all-cause mortality.

**Do you use an oral switch in all patients with SAB?**

The trial has shown that early oral switch is possible. However, careful assessment of the individual patient, preferably by consulting an expert in infectious disease, is necessary to correctly assign a patient to the low-risk group and close monitoring for early detection of complications and potential prolongation of antimicrobial therapy is advised.

**What should future trials do?**

Future complementary trials should be conducted that assess early oral switch therapy in patients with medium-risk and high-risk SAB and requirement for longer treatment duration. This will potentially address a larger number of cases but will be a particular challenge due to the pleomorphic nature of SAB and will require sophisticated diagnostic pathways to assure source control in the study protocols. Also, alternative oral agents, such as high-dose oral beta-lactams or tetracyclines merit further study.

**Why was the study not published immediately after completion?**

The completion of the study was during the SARS-CoV-2 pandemic. Many authors, esp. the lead authors and the statisticians, were heavily involved in the pandemic response. This led to large delays in analyzing the data and preparing the manuscript.

6 Committee for Medicinal Products for Human Use. Points to consider on switching between superiority and non-inferiority (CPMP/EWP/482/99)., 2000.

7 Committee for Medicinal Products for Human Use. Guideline on the evaluation of medicinal products indicated for treatment of bacterial infections (CPMP/EWP/558/95 rev 2)., 2011.

8 Evans S, Rubin DB, Powers JH, Follmann D. Analysis Populations in Anti-Infective Clinical Trials: Whom to Analyze? *Stat Commun Infect Dis* 2018; **10. https://doi.org/**10.1515/scid-2017-0002.

9 Jorgensen SCJ, Lagnf AM, Bhatia S, Shamim M-D, Rybak MJ. Sequential intravenous-to-oral outpatient antibiotic therapy for MRSA bacteraemia: one step closer. *J Antimicrob Chemother* 2019; **74:** 489–98. https://doi.org/10.1093/jac/dky452.

10 Kouijzer IJE, van Leerdam EJ, Gompelman M, et al. Intravenous to Oral Switch in Complicated Staphylococcus aureus Bacteremia without Endovascular Infection: A Retrospective Single-Center Cohort Study. *Clin Infect Dis* 2021; **73:** 895–98. https://doi.org/10.1093/cid/ciab156.

11 Willekens R, Puig-Asensio M, Ruiz-Camps I, et al. Early Oral Switch to Linezolid for Low-risk Patients with Staphylococcus aureus Bloodstream Infections: A Propensity-matched Cohort Study. *Clin Infect Dis* 2019; **69:** 381–87. https://doi.org/10.1093/cid/ciy916.

12 Bupha-Intr O, Blackmore T, Bloomfield M. Efficacy of Early Oral Switch with β-Lactams for Low-Risk Staphylococcus aureus Bacteremia. *Antimicrob Agents Chemother* 2020; **64:** e02345-19. https://doi.org/10.1128/AAC.02345-19.

13 Mun SJ, Kim S-H, Huh K, et al. Oral step-down therapy in patients with uncomplicated Staphylococcus aureus primary bacteremia and catheter-related bloodstream infections. *J Chemother* 2022: 1–7. https://doi.org/10.1080/1120009X.2022.2031469.

14 Itoh N, Hadano Y, Saito S, Myokai M, Nakamura Y, Kurai H. Intravenous to oral switch therapy in cancer patients with catheter-related bloodstream infection due to methicillin-sensitive Staphylococcus aureus: A single-center retrospective observational study. *PLoS One* 2018; **13:** e0207413. https://doi.org/10.1371/journal.pone.0207413.

15 Yeager SD, Oliver JE, Shorman MA, Wright LR, Veve MP. Comparison of linezolid step-down therapy to standard parenteral therapy in methicillin-resistant Staphylococcus aureus bloodstream infections. *Int J Antimicrob Agents* 2021; **57:** 106329. https://doi.org/10.1016/j.ijantimicag.2021.106329.

16 Pérez-Rodríguez MT, Sousa A, Moreno-Flores A, et al. The benefits and safety of oral sequential antibiotic therapy in non-complicated and complicated Staphylococcus aureus bacteremia. *Int J Infect Dis* 2021; **102:** 554–60. https://doi.org/10.1016/j.ijid.2020.10.097.

17 Wald-Dickler N, Holtom PD, Phillips MC, et al. Oral Is the new IV. Challenging Decades of Blood and Bone Infection Dogma: A Systematic Review. *Am J Med* 2022; **135:** 369-379.e1. https://doi.org/10.1016/j.amjmed.2021.10.007.

18 Wilcox M, Nathwani D, Dryden M. Linezolid compared with teicoplanin for the treatment of suspected or proven Gram-positive infections. *J Antimicrob Chemother* 2004; **53:** 335–44. https://doi.org/10.1093/jac/dkh088.

19 Wilcox MH, Tack KJ, Bouza E, et al. Complicated skin and skin-structure infections and catheter-related bloodstream infections: noninferiority of linezolid in a phase 3 study. *Clin Infect Dis* 2009; **48:** 203–12. https://doi.org/10.1086/595686.
20 Stevens DL, Herr D, Lampiris H, Hunt JL, Batts DH, Hafkin B. Linezolid versus vancomycin for the treatment of methicillin-resistant Staphylococcus aureus infections. *Clin Infect Dis* 2002; **34:** 1481–90. https://doi.org/10.1086/340353.
21 Schrenzel J, Harbarth S, Schockmel G, et al. A randomized clinical trial to compare fleroxacin-rifampicin with flucloxacillin or vancomycin for the treatment of staphylococcal infection. *Clin Infect Dis* 2004; **39:** 1285–92. https://doi.org/10.1086/424506.
22 Markowitz N, Quinn EL, Saravolatz LD. Trimethoprim-sulfamethoxazole compared with vancomycin for the treatment of Staphylococcus aureus infection. *Ann Intern Med* 1992; **117:** 390–98. https://doi.org/10.7326/0003-4819-117-5-390.
23 Paul M, Bishara J, Yahav D, et al. Trimethoprim-sulfamethoxazole versus vancomycin for severe infections caused by meticillin resistant Staphylococcus aureus: randomised controlled trial. *BMJ* 2015; **350:** h2219. https://doi.org/10.1136/bmj.h2219.
24 Harbarth S, Dach E von, Pagani L, et al. Randomized non-inferiority trial to compare trimethoprim/sulfamethoxazole plus rifampicin versus linezolid for the treatment of MRSA infection. *J Antimicrob Chemother* 2015; **70:** 264–72. https://doi.org/10.1093/jac/dku352.
25 Iversen K, Ihlemann N, Gill SU, et al. Partial Oral versus Intravenous Antibiotic Treatment of Endocarditis. *N Engl J Med* 2019; **380:** 415–24. https://doi.org/10.1056/NEJMoa1808312.
26 Kaasch AJ, Fowler VG, Rieg S, et al. Use of a simple criteria set for guiding echocardiography in nosocomial Staphylococcus aureus bacteremia. *Clin Infect Dis* 2011; **53:** 1–9. https://doi.org/10.1093/cid/cir320.
27 Holland TL, Raad I, Boucher HW, et al. Effect of Algorithm-Based Therapy vs Usual Care on Clinical Success and Serious Adverse Events in Patients with Staphylococcal Bacteremia: A Randomized Clinical Trial. *JAMA* 2018; **320:** 1249–58. https://doi.org/10.1001/jama.2018.13155.
28 Dagher M, Fowler VG, Wright PW, Staub MB. A Narrative Review of Early Oral Stepdown Therapy for the Treatment of Uncomplicated Staphylococcus aureus Bacteremia: Yay or Nay? *Open Forum Infect Dis* 2020; **7:** ofaa151. https://doi.org/10.1093/ofid/ofaa151.
