## Supplementary material for "Early oral switch in low-risk *Staphylococcus aureus* bloodstream infection": Study Protocol

TRIAL PROTOCOL

ACRONYM: SABATO (Staphylococcus aureus Bacteremia Antibiotic Treatment Options)

**Sponsor:**
Heinrich-Heine-Universität Düsseldorf
Universitätsstr.1
40225 Düsseldorf
Germany

**Principal Coordinating Investigator:**
Prof. Dr. med. Achim Kaasch
Institute of Medical Microbiology and Hospital Hygiene
Düsseldorf University Hospital
Universitätsstr.1
40225 Düsseldorf
Germany

Trial protocol code: Uni-Koeln-1400

ClinicalTrials.gov Identifier: NCT01792804

German Clinical Trials Register Identifier: DRKS00004741

EudraCT number: 2013-000577-77

Amendment I, Version v01-01-F, 03 December 2014, Study Protocol Version v03-04-F of 03 December 2014

Amendment II, Version V01_0, 20JUL 2016, Study Protocol Version V05_0 of 20JUL2016

Amendment III Version V01_0, Study Protocol Version V07-F of 20MAR2018

Amendment IV Version V01_0, Study Protocol Version V08-F of 18JUL2019

The information in this trial protocol is strictly confidential. It is for the use of the sponsor, investigator, trial personnel, ethics committee, the authorities, and trial subjects only. This trial protocol may not be passed on to third parties without the express agreement of the sponsor or the Principal Coordinating Investigator.

### I. Signatures

**Prof. Dr. Achim Kaasch**
On behalf of the sponsor
Heinrich-Heine-Universität Düsseldorf
and
**Principal Coordinating Investigator (PCI)**
and Clinical Project Manager
Institute of Medical Mikrobiology and Hospital Hygiene
Düsseldorf University Hospital

18. Juli 2019 A. Kaasch

Date Signature

**Prof. Dr. Martin Hellmich**
Statistician
Institute of Medical Statistics and
Computational Biology, University of Cologne

Date Signature

### I. Signatures

Prof. Dr. Achim Kaasch
On behalf of the sponsor
Heinrich-Heine-Universität Düsseldorf
and
Principal Coordinating Investigator (PCI)
and Clinical Project Manager
Institute of Medical Mikrobiology and Hospital Hygiene
Düsseldorf University Hospital

Date Signature

Prof. Dr. Martin Hellmich
Statistician
Institute of Medical Statistics and
Computational Biology, University of Cologne

18.7.2019 [signature]

Date Signature

### II. Synopsis

| | |
|---|---|
| Sponsor: | Heinrich-Heine-Universität Düsseldorf<br>Universitätsstr. 1<br>40225 Düsseldorf<br>Germany<br><br>Represented by:<br>Prof. Dr. med. Achim Kaasch<br>Institute of Medical Microbiology and Hospital Hygiene<br>Düsseldorf University Hospital<br>Universitätsstr. 1<br>40255 Düsseldorf, Germany |
| Principal Coordinating Investigator: | See above |
| Title of the clinical trial: | Early oral switch therapy in low-risk Staphylococcus aureus bloodstream infection (SABATO=S. aureus Bacteremia Antibiotic Treatment Options) |
| Indication: | Staphylococcus aureus bloodstream infection |
| Phase: | III (therapeutic confirmatory) |
| Type of trial, trial design, methodology: | Multicenter, multinational clinical trial<br>Two arms, randomized, open-label, non-inferiority, parallel-group |
| Number of subjects: | 215 allocated 1:1 in 2 treatment groups |

| | |
|---|---|
| Primary trial objective: | To demonstrate that in patients with low-risk S. aureus bloodstream infection (SAB) a switch from intravenous to oral antimicrobial therapy (oral switch therapy, OST) is non-inferior to a conventional course of intravenous therapy (intravenous standard therapy, IST) |
| Study endpoints: | Primary endpoint:<br>• SAB-related complications (relapsing SAB, deep-seated infection with S. aureus, or attributable mortality) within 90 days<br>Secondary endpoint:<br>• Length of hospital stay<br>Other variables:<br>• 14, 30, and 90-day survival<br>• Complications of intravenous therapy |
| Criteria for evaluation: | Efficacy:<br>• SAB-related complications (by telephone / in person interviews), blood cultures, imaging for assessment of deep-seated infections, vital signs (esp. body temperature), 90-day mortality, length of stay in ICU and in hospital, complications of intravenous therapy<br>Safety:<br>• Clostridium difficile associated diarrhea (CDAD)<br>• AEs and SAEs |

| | |
|---|---|
| Medical Condition and Principal inclusion criteria: | Medical condition or disease to be investigated:<br>• Staphylococcus aureus bloodstream infection<br><br>Principal inclusion criteria:<br>• Blood culture positive for S. aureus not considered to represent contamination<br>• 5-7 days of adequate intravenous antimicrobial therapy<br><br>Principal exclusion criteria:<br>• Polymicrobial bloodstream infection<br>• Signs and symptoms of complicated SAB (deep-seated infection, hematogenous dissemination, septic shock, prolonged bacteremia)<br>• Severe comorbidity |
| Name of study drug: | Protocol-approved antimicrobial (s. below) with administration route according to study arm |
| Study drug – dosage and method of administration: | Orally administered antimicrobial – Oral switch therapy (OST)<br>First choice (MRSA and MSSA): trimethoprim-sulfamethoxazole 160/800mg q12h<br>Second choice (MSSA): clindamycin 600 mg q8h<br>Second choice (MRSA): linezolid 600 mg q12h |
| Study drug used as a comparator – dosage and method of administration: | Intravenously aministered antimicrobial – Intravenous standard therapy (IST)<br>First choice (MSSA): flucloxacillin 2g q6h<br>[Spain/France: cloxacillin (2g q6h)] or cefazolin 2g q8h<br>Second choice (MSSA): vancomycin 1g q12h<br>First choice (MRSA): vancomycin 1g q12h<br>Second choice (MRSA): daptomycin 6 mg/kg q24h) |

| | | |
|---|---|---|
| Duration of treatment: | 7-9 days | |
| Time plan: | First patient first visit (FPFV): | 20 December 2013 |
| | Last patient first visit (LPFV): | 31 January 2019 |
| | Last patient last visit (LPLV): | 30 April 2020 |
| | Final study report: | 30 April 2021 |

Statistician:

Prof. Dr. Martin Hellmich
Institute of Medical Statistics and Computational Biology (IMSB)
University of Cologne
Bachemer Str. 86
50931 Cologne
Germany

Statistical methods:

Efficacy: Non-inferiority (with margins of 10 and 5 percentage points) regarding incidence of SAB-related complications at 90 days as tested by Zhao’s test of non-null hypothesis on proportions stratified by center based on the per-protocol set (PP, primary analysis set); the full analysis set (intention-to-treat, ITT) is of equal importance and should lead to similar conclusions for a robust interpretation.

Safety and other endpoints: Descriptive methods, e.g. contingency tables and listings, generalized linear modeling, methods for rates and proportions

| | |
|---|---|
| GCP conformance: | The present trial will be conducted in accordance with the valid versions of the trial protocol and the internationally recognised Good Clinical Practice Guidelines (ICH-GCP), including archiving of essential documents |
| Financing: | Deutsche Forschungsgemeinschaft (DFG, German Research Foundation; grant number KA 3104/2-1) |

### III. Table of contents


II. Synopsis 3

III. Table of contents 8

a. List of tables 13

b. List of figures 13

IV. Abbreviations 14

1. Introduction 16

2. Objectives of the clinical trial 19

2.1. Rationale for the clinical trial 19

2.2. Primary objective 19

2.3. Secondary and other objectives 19

3. Organisational and administrative aspects of the trial 21

3.1. Sponsor 21

3.2. Statistics 21

3.3. Data Monitoring Committee 22

3.4. Further committees 22

3.4.1. Steering Committee 22

3.4.2. Scientific Advisory Committee 23

3.4.3. Clinical Review Committee 23

3.5. Study laboratories and other technical services 23

3.6. Central organisation units 23

3.7. Investigators and trial sites 24

3.8. Financing 25

4. Trial conduct 26

4.1. General aspects of trial design 26
4.1.1. Time plan 26
4.2. Discussion of trial design 29
4.3. Selection of trial population 30
4.3.1. Inclusion criteria 30
4.3.2. Exclusion criteria 31
4.4. Withdrawal of trial subjects after trial start 36
4.4.1. Procedures for premature withdrawal from treatment or trial 36
4.5. Closure of trial sites/Premature termination of the clinical trial 37
4.5.1. Closure of trial sites 37
4.5.2. Premature termination of trial 38
4.6. Treatment 38
4.6.1. Treatments to be given 38
4.6.2. Description of study drugs 40
4.6.2.1. Manufacture of study drugs 41
4.6.2.2. Labelling of study drugs 41
4.6.2.3. Storage and reconstitution of study drugs 41
4.6.3. Compliance with treatment / Dispensing and return of study drugs 41
4.6.4. Assignment of trial subjects to treatment groups 41
4.6.5. Dose selection of study drugs 42
4.6.6. Time of administration and dose adjustments of study drugs in the individual trial subject 44
4.6.7. Blinding 46
4.6.7.1. Unblinding 46
4.6.8. Previous and concomitant medication 46

4.6.8.1. Rescue therapy for emergencies 47

4.6.9. Continuation of treatment after the end of the clinical trial 47

4.7. Efficacy and safety variables 47

4.7.1. Measurement of efficacy and safety variables 48

4.7.1.1. Primary target variable 48

4.7.1.2. Secondary and other target variables 50

4.7.1.3. Safety analysis 51

4.7.1.4. Description of visits 51

4.7.2. Rationale for assessment procedures 55

4.7.3. Pharmacokinetics/Determination of drug levels 56

4.8. Data quality assurance 56

4.8.1. Monitoring 56

4.8.2. Audits/Inspections 57

4.9. Documentation 58

4.9.1. Data management 58

4.9.2. Archiving 59

5. Ethical and regulatory aspects 60

5.1. Independent ethics committee 60

5.2. Ethical basis for the clinical trial 60

5.2.1. Legislation and guidelines used for preparation 60

5.3. Notification of the authorities, approval and registration 61

5.4. Obtaining informed consent from trial subjects 61

5.5. Insurance of trial subjects 62

5.6. Data protection 62

6. Statistical methods and sample size calculation 63

6.1. Statistical and analytical plan 63
6.1.1. Analysis sets 63
6.1.2. Description of trial subject groups 65
6.1.3. Primary target variable 65
6.1.4. Secondary target variables 66
6.1.5. Safety variables 66
6.1.6. Subgroup analyses 66
6.1.7. Interim analysis 66
6.2. Sample size calculation 67
7. Safety 69
7.1. Definitions of adverse events and adverse drug reactions 69
7.1.1. Adverse event 69
7.1.2. Adverse drug reaction 70
7.1.3. Serious adverse events and serious adverse reactions 70
7.1.4. Unexpected adverse drug reaction 70
7.1.5. Suspected unexpected serious adverse reactions 71
7.1.6. Other possible trial-specific complications or risks 71
7.2. Documentation and follow-up of adverse events 72
7.2.1. Documentation of adverse events and adverse drug reactions 72
7.2.2. Severity of the adverse event 72
7.2.3. Causal relationship between adverse event and study drugs 73
7.3. Reporting and follow up of serious adverse events, pregnancy and changes in risk-benefit assessment 74
7.3.1. SAE reports from the authorized member of the study group to the sponsor 75
7.3.2. Pregnancy reports from the study group to the sponsor 75

7.3.3. Unblinding when treatment is blinded 76
7.3.4. Notification of ethics committee and competent authorities 76
7.3.5. Review and reporting of changes in the risk-benefit ratio 76
7.3.6. Informing the Data Monitoring Committee 77
7.3.7. Informing the investigators 77
7.3.8. Informing the marketing authorisation holder 77
7.4. Annual safety report of trial subjects 78
8. Use of trial findings and publication 79
8.1. Reports 79
8.1.1. Interim reports 79
8.1.2. Final report 79
8.2. Publication 79
9. Amendments to the trial protocol 81
10. References 82
11. Appendices 87
11.1. Trial sites and principle investigators 87
11.2. Protocol Agreement Form 88
11.3. Steering Committee 88
11.4. Data Monitoring Committee 89
11.5. Scientific Advisory Committee 89
11.6. Clinical Review Committee 90
11.7. Central study laboratory 90
11.8. Reference documents for study drugs 90
11.9. Patient information sheet and informed consent form 91
11.10. Prescribing information 91

11.11. Confirmation of insurance 91

11.12. Conditions of insurance 91

#### a. List of tables

Table 1: Time plan of the trial 28

Table 2: Dosing of study drugs 43

Table 3: Dose adjustment for trimethoprim-sulfamethoxazole 44

Table 4: Dose adjustment for flucloxacillin 45

Table 5: Dose adjustment for cefazolin 45

Table 6: Dose adjustment for vancomycin 45

Table 7: Dose adjustment for daptomycin 45

Table 8: Study flow chart of the clinical trial 54

Table 9: Visit schedule 55

#### b. List of figures

Figure 1: Trial flowchart 28

Figure 2: Analysis sets 64

### IV. Abbreviations

| Abbreviation | Meaning |
|---|---|
| ACCP/SCCM | American College of Chest Physicians/Society of Critical Care Med. |
| ADR | Adverse drug reaction |
| AE | Adverse event |
| AEMyPS | Agencia Española de Medicamentos y Productos Sanitarios |
| AMG | Arzneimittelgesetz (German Federal Drug Law) |
| BfArM | Bundesinstitut für Arzneimittel und Medizinprodukte (Federal Institute for Drugs and Medical Devices) |
| CDAD | Clostridium difficile associated diarrhoe |
| CTCAE | Common Terminology Criteria of Adverse Events |
| CRC | Clinical Review Committee |
| DMC | Data Monitoring Committee |
| DSUR | Development Safety Update Report |
| eCRF | Electronic case report form |
| EOS | End of study |
| EOT | End of treatment |
| FU1 | Follow-up visit 1 |
| GCP | Good Clinical Practice |
| GI | Gastrointestinal |
| ICH | International Conference on Harmonisation |
| ICU | Intensive Care Unit |
| IRB | Institutional Review Board (Ethics Committee) |

| | |
|---|---|
| IST | Intravenous standard therapy |
| ITT | Intention to treat |
| MEB | Medicines Evaluation Board |
| MHRA | Medicines and Healthcare products Regulatory Agency |
| MRSA | Methicillin-resistant Staphylococcus aureus |
| MSSA | Methicillin-susceptible Staphylococcus aureus |
| OPAT | Outpatient parenteral antimicrobial therapy |
| OST | Oral switch therapy |
| PCI | Principal Coordinating Investigator |
| PEI | Paul-Ehrlich-Institut |
| PFGE | Pulsed-field gel electrophoresis |
| PP | Per Protocol |
| RCT | Randomized Controlled Trial |
| SAB | Staphylococcus aureus bloodstream infection |
| SAC | Scientific Advisory Committee |
| SADR | Serious adverse drug reaction |
| SC | Steering Committee |
| SAE | Serious Adverse Event |
| SOC | Standard of care |
| SUSAR | Suspected Unexpected Serious Adverse Reaction |
| TDM | Therapeutic drug monitoring |
| ZKS | Zentrum für Klinische Studien (Clinical Trials Center), Cologne |

### 1. Introduction

Increasing resistance to antimicrobial agents has been recognized as a major health problem worldwide that will even aggravate due to the lack of new antimicrobial agents within the next decade (1). This threat underscores the need to maximize clinical utility of existing antimicrobials, through more rational prescription, e.g. optimizing duration of treatment.

Staphylococcus aureus bloodstream infection (SAB) is a major cause for prolonged antimicrobial therapy. With an approximate incidence of 25 cases per 100,000, about 200,000 cases occur annually in Europe (2). Recent data for Western Europe demonstrate a crude mortality of 20-30% (in-hospital or 30-day mortality) in patients with SAB (2).

In many cases SAB can be cured by antimicrobial therapy. However, SAB differs from other bloodstream infections with respect to SAB-related complications: relapses, local extension and distant metastatic foci are relatively common events and occur in about 2-25% of infections (3-5). Therefore, antbiotic therapy is considered to be especially important in this disease and standard treatment schedules are significantly longer than in other bloodstream infections.

A course of at least 14 days of intravenous antimicrobials is considered standard therapy (6-8) in “uncomplicated SAB”. Generally, “uncomplicated SAB” is defined by absence of: community acquisition, skin examination findings suggesting acute systemic infection, positive follow-up blood cultures and persistent fever at 72h (9). Shorter courses of intravenous treatment are currently not recommended due to the lack of sound clinical evidence. The SABATO trial will specifically address this issue and examine the effectiveness and safety of an abbreviated course of intravenous therapy in patients that have a low-risk of SAB-related complications.

This trial poses specific risks for the patient. A shorter course of effective antimicrobial therapy may lead to relapsing SAB, local spread of the infection, or hematogenous dissemination of S. aureus with resulting deep-seated infection. To minimize the risk, a population of patients with a very low-risk of SAB-related complications is described by in- and exlusion criteria. This population has been validated by using data from two prospective cohort studies. Data from the INSTINCT (Invasive Staphylococcus aureus Infection Cohort) study (10) shows a low incidence of SAB-related complications in low-risk patients (3%; 4 of 135 patients). A pilot study for the SABATO trial with 236 SAB patients from 10 German study centers provided further evidence for a very low risk of complications in these patients: Only 1 of 89 patients had a SAB-related complication.

Abbreviated or early i.v. to oral switch treatment strategies have been successfully applied to other infectious diseases such as nosocomial pneumonia (11), meningococcal disease (12), and febrile neutropenia (13). These strategies allow shorter intravenous antimicrobial therapy and offer options for early discharge from hospital. This, in turn, increases the patients' quality of life, decreases treatment costs, reduces the risk of nosocomial infections and may help to diminish antimicrobial resistance development and spread.

The SABATO trial is the first randomized controlled trial addressing early oral switch therapy in SAB. In a recent search (January 2013) of PubMed and metaRegister of controlled trials (www.controlled-trials.com) no trials on oral switch therapy in SAB were identified.

In fact, there have been very few trials on SAB. Less than 1,500 patients with SAB have been randomized in 16 controlled trials of antimicrobial therapy published over the last 45 years. The recommendations on treatment duration of SAB are mainly based on expert opinion and a few observational epidemiological studies.

There is one controlled trial with 11 patients on short course therapy. The trial compared a 2-week intravenous antimicrobial regimen with a 4-week course in patients with uncomplicated SAB and failed to show a difference (14). A meta-analysis of this controlled trial and 10 uncontrolled, epidemiological studies (15) showed great potential for bias imprecision and recommended randomized trials.

Although not supported by current recommendations, shorter duration of intravenous therapy has become management practice for SAB in some countries: Thwaites et al. reviewed management practices of patients with SAB in 8 UK centers and found that 25% of patients received oral antimicrobials alone for more than 50% of the treatment duration whereas 16% of patients received less than the recommended 14 days of therapy (16). In this study efficacy and safety of oral therapy was not assessed.

The effectiveness of oral antimicrobial therapy in SAB has been assessed in a single controlled study: In a randomized controlled trial (17), 104 patients with SAB either received oral fleroxacin plus rifampicin or intravenous study therapy. The cure rate in both groups was similar (82% vs. 80%). Therefore, in principle SAB can be treated with orally administered antimicrobials.

### 2. Objectives of the clinical trial

#### 2.1. Rationale for the clinical trial

A successfully performed trial will have a great impact on clinical decision making worldwide. It will provide a rationale for optimizing treatment of patients with low-risk SAB - a common clinical scenario - that will most certainly be integrated into evidence-based treatment guidelines. An early switch to oral therapy may have an impact on patient's well-being: an abbreviated hospital stay can increase the quality of life and reduces the risk of line-associated infections. For hospitals, earlier discharge may result in significant cost savings.

#### 2.2. Primary objective

The hypothesis is that a switch from intravenous to oral antimicrobial therapy is non-inferior to standard intravenous therapy in patients with low-risk SAB. Therefore, the primary objective of the trial is to demonstrate, that oral switch therapy (OST) is as safe and effective as intravenous standard therapy (IST). This will be achieved by comparing the rate of **SAB-related complications** (relapsing SAB, deep-seated infection with S. aureus, or mortality attributable to SAB) within 90 days.

Low-risk SAB manifests itself typically in patients with comorbidities. Therefore, survival is largely determined by the underlying disease (18) and was not chosen as a primary endpoint. However, death related to SAB is comprised in the primary endpoint. Death unrelated to SAB will be carefully evaluated and compared.

#### 2.3. Secondary and other objectives

The secondary objective is to measure the potential benefit for the patient. This is achieved by evaluating the **length of hospital stay** after the first positive blood culture and **complications of intravenous therapy**. A considerable number of patients on OST are expected to be discharged earlier from hospital, since hospital stay due to intravenous therapy is no longer required. This will reduce the risks associated with hospitalization and i.v. therapy (catheter-related infection, venous thrombosis, and septic thrombophlebitis) and is likely to improve patients’ quality of life.

### 3. Organisational and administrative aspects of the trial

#### 3.1. Sponsor

| | |
|---|---|
| Sponsor: | Heinrich-Heine-Universität Düsseldorf<br>Universitätsstr. 1<br>40225 Düsseldorf<br>Germany |
| Represented by: | Prof. Dr. med. Achim Kaasch<br>Institute of Medical Microbiology and Hospital Hygiene<br>Düsseldorf University Hospital<br>Universitätsstr.1<br>40225 Düsseldorf<br>Germany |

Principal Coordinating Investigator (PCI):

Prof. Dr. med. Achim Kaasch
Institute of Medical Microbiology and Hospital Hygiene
Düsseldorf University Hospital
Universitätsstr.1
40225 Düsseldorf
Germany

In the subsequent text, the PCI will be described either in his or her function as PCI or as a representative of the sponsor.

#### 3.2. Statistics

| | |
|---|---|
| Statistician: | Prof. Dr. Martin Hellmich<br>Institute of Medical Statistics and Computational Biology (IMSB)<br>University of Cologne |

Bachemer Strasse 86
50931 Cologne
Germany

#### 3.3. Data Monitoring Committee

A Data Monitoring Committee (DMC) made up of independent experts will be set up. It consists of two physicians and a statistician who are not involved in the conduct of the trial (see Section 11.4). The task of the DMC is to oversee the safety of the trial subjects in the clinical trial by periodically assessing the safety and efficacy of the trial therapy, and to monitor the integrity and validity of the data collected and the conduct of the clinical trial. One of the DMC members will be appointed to be the chairperson. The members of the DMC will neither be clinical investigators involved in the study nor employees of the sponsors. All activities of the DMC will be documented including data summaries and analysis provided to the DMC. Documentation will remain confidential within the DMC until the study is finished.

Throughout this process of surveillance, the DMC provides the sponsor with recommendations with regard to continuing the trial (e.g. termination or modification) based on the data collected. The data necessary for the DMC to fulfill this function are provided by the sponsor as determined by the DMC. Amongst other datasets, these must include listings providing information on serious adverse events (including SAB-related complications) and further variables that the DMC considers necessary in appropriate intervals at least every 6 months and when formal interim analyses are conducted. SAB-related complications are to be forwarded to the DMC without delay. Data is at first provided blinded to treatment arm and can be unblinded upon request of the DMC.

#### 3.4. Further committees

##### 3.4.1. Steering Committee

The Steering Committee (SC) is responsible for protocol development and oversight of study progress. It will take majority decisions. For questions with special relevance to international study sites, the SC will be enlarged to comprise national representatives for each country. The SC will convene as needed. A list of the members is given in Appendix 11.3.

##### 3.4.2. Scientific Advisory Committee

The Scientific Advisory Committee (SAC) gives advice on all aspects of the trial, esp. on trial design. It will convene as needed. A list of the members of the SAC is given in Appendix 11.5.

##### 3.4.3. Clinical Review Committee

The masked Clinical Review Committee (CRC) will be responsible for evaluating cases regarding protocol violations, and treatment failures blinded for treatment arm. If felt necessary, unblinding can be requested by the CRC for an individual case. A list of the members of the CRC is given in Appendix 11.6.

#### 3.5. Study laboratories and other technical services

In patients with recurrent S. aureus infections, isolates from the first and the recurrent episode will be compared based on susceptibility data and genetic molecular characteristics (e.g. pulsed-field gel electrophoresis) by local microbiological laboratories. In case local microbiological laboratories have conflicting results, genetic analysis will be performed in a central laboratory (see Appendix 11.7).

#### 3.6. Central organisation units

| | |
|---|---|
| Project management: | Institut für Medizinische Mikrobiologie und Krankenhaushygiene<br>Universitätsklinikum der Heinrich-Heine-Universität Düsseldorf,<br>40225 Düsseldorf<br>Germany |

| | |
|---|---|
| Monitoring: | Koordinierungszentrum für Klinische Studien Düsseldorf (KKSD)<br>Moorenstr. 5<br>40225 Düsseldorf<br>Germany |
| Data management: | Clinical Trials Center Cologne (ZKS Köln) |
| SAE management: | Clinical Trials Center Cologne (ZKS Köln)<br>Gleueler Strasse 269<br>50935 Cologne<br>Germany |

##### 3.7. Investigators and trial sites

This clinical trial will be carried out as a multicentre open trial at trial sites in Germany, trial sites in the Netherlands, trial sites in France and trial sites in Spain (see Appendix 11). If necessary, further qualified trial sites may be recruited to the trial.

A list of the trial sites with names of the principal investigators is given in Appendix 11.1. The listing of trial sites, principal investigators, subinvestigators, and further trial staff, will be kept and continuously updated in a separate list. The final version of this list will be attached to the final report of the clinical trial.

Only investigators and participating trial sites are selected for the SABATO trial that meet the regulatory requirements with qualification and experience to perform a clinical investigation including trials of pharmaceutical preparations. The sponsor will appoint the principle investigator at each study site, who in turn, will select qualified and experienced staff for trial conduction.

Requirements for investigators and trial sites

- all investigators need to be physicians with proof of knowledge of regulatory procedures, e.g. with track record of conducting clinical studies

- a trial site needs adequate personnel for conducting the trial
- principle investigators need to have experience with management and treatment of S. aureus bloodstream infection as well as knowledge of current standard of care
- access to microbiological laboratory with state-of-the-art testing procedures, and fast information relay
- investigators need access to basic clinical laboratory testing

##### 3.8. Financing

The clinical trial will be funded by the Deutsche Forschungsgemeinschaft (DFG; German Research Foundation; grant number KA 3104/2-1).

### 4. Trial conduct

#### 4.1. General aspects of trial design

Phase III, multicenter, open-label, randomized, controlled, non-inferiority trial with a total of 215 patients enrolled.

##### 4.1.1. Time plan

The trial starts with the first patient visit and ends with the last visit of the last patient (for time line see table 1).

Individual patients go through a screening, intervention and follow-up phase (s. figure 1).

First, patients with SAB are reported from the microbiological department to the principle investigator. Then, individual patients with SAB are screened for possible enrolment by the principle investigator.

The intervention phase (7-9 days) starts when patients have given informed consent and all in- and exclusion requirements are fulfilled.The length of the intervention phase depends on how long patients have received appropriate pre-randomization antimicrobials. All patients will receive an overall course of 14 days appropriate antimicrobial therapy, e.g. patients having received five days of appropriate pre-randomization antimicrobials will receive nine days of OST or IST.

Patients on OST can be discharged to home before end of therapy (EOT) according to clinical and psychosocial criteria. Patients on IST can only be discharged to home when an OPAT (outpatient parenteral antimicrobial therapy) service is in operation at the local study site and reliable intravenous medication can be assured. Patients on either IST or OST can be transferred to another hospital or rehabilitation unit when it is assured that the patient will receive study medication for the required duration and reliable information study medication (batch number, dosing, duration) and adverse events can be obtained by study personnel.

The follow-up phase starts at EOT and ends 90 days after the first positive blood culture. Patients that are still in hospital will be visited on the ward to collect follow-up information. Discharged patients are followed by a structured telephone interview at day 85-99.

**Table 1: Time plan of the trial**

| | |
|---|---|
| First patient first visit (FPFV): | 20 December 2013 |
| Last patient first visit (LPFV): | 31 January 2020 |
| Last patient last visit (LPLV): | 30 April 2020 |
| Final study report: | 30 April 2021 |

**Figure 1: Trial flowchart**

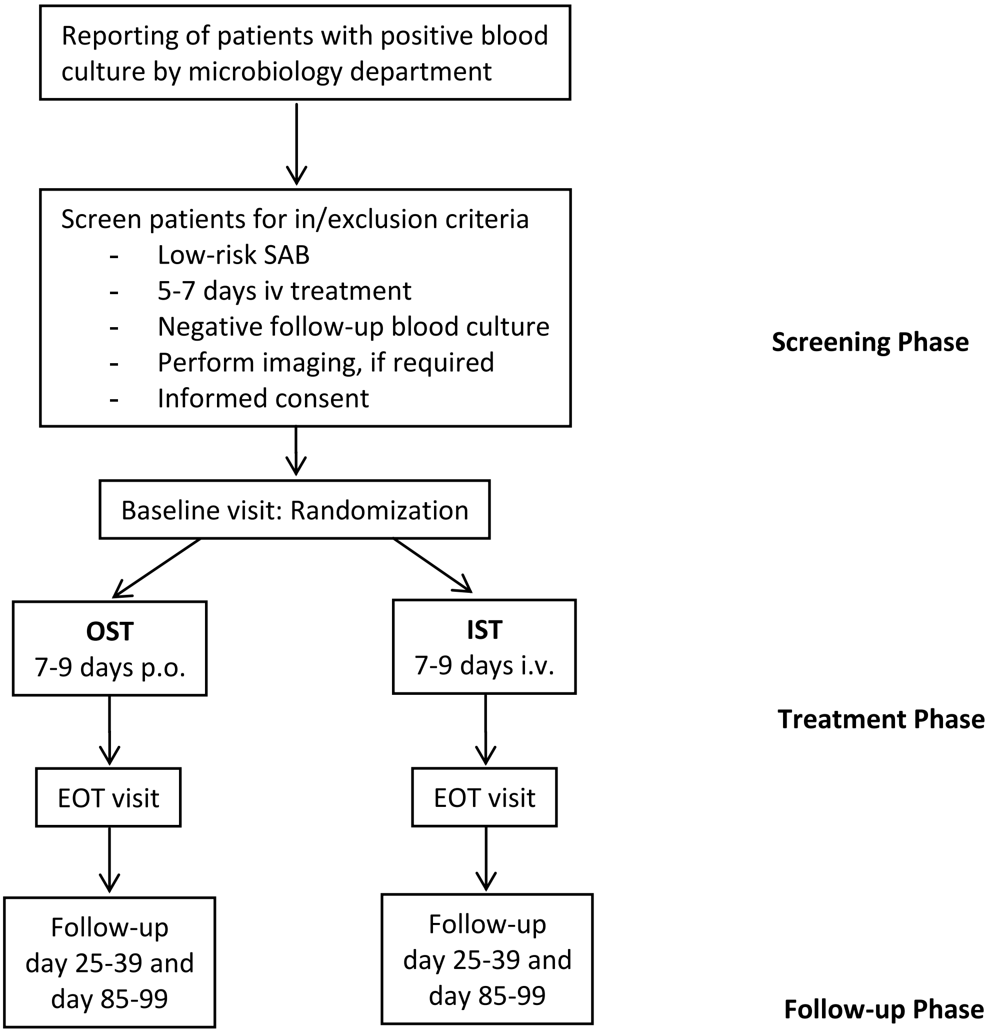

#### 4.2. Discussion of trial design

This trial compares OST to IST in a parallel group design, which is the preferred design for a non-inferiority study of a largely monophasic, curable disease. Intravenous treatment for 14 days is standard therapy for patients with low-risk SAB in all study centers. In this trial the overall duration of appropriate antimicrobial therapy will be kept at 14 days, however the mode of administration (oral vs. intravenous) differs between the study arms.

Patients are randomly allocated to treatment arms (1:1) by means of the central 24-7 Internet randomization service ALEA (stratified by study center, permuted blocks of varying length). Neither patients nor investigators can be fully masked (regarding treatment), however patients will be carefully instructed and investigators trained to comply with study procedures. Compliance with oral follow-up medication after discharge will be assessed by telephone calls at least every two to three days. Adverse events and endpoints will be either reported by the patient to the principle investigator, or captured at the follow-up interview.

Key efficacy endpoints can be determined in a rather objective manner (e.g. bacterial culture from deep-seated infection, death); however, final assessment of clinical response and absence of complications will be done by a masked Clinical Review Committee. Most cases of recurrent S. aureus infections occur within 30 to 60 days of the first positive bloodculture. The later a recurrent infection occurs, the more likely it is an independent infection event and not a late complication of the initial infection. Therefore, two follow-up telephone contacts 30 and 90 days after the first positive blood culture are considered sufficient to detect recurrent infection. In the case of a suspected recurrent infection, the newly isolated strain and the initial isolate will e compared by PFGE-analysis or spa-typing to differentiate between relapse and an independent infection.

According to the guideline on the evaluation of medicinal products indicated for treatment of bacterial infections (CPMP/EWP/558/95 rev 2) (47), “In all studies there should at least be a comparison between the planned primary analysis and an analysis of all randomized patients in which indeterminate or missing outcomes are counted as failures.” Moreover, the guideline CPMP/EWP/482/99 (48) states, “In a non-inferiority trial, the full analysis set and PP analysis set have equal importance and their use should lead to similar conclusions for a robust interpretation." Thus, the primary analysis is based on the per-protocol set; the full analysis set (intention-to-treat, all randomized patients, indeterminate or missing outcomes counted as failures) will be of equal importance and should lead to similar conclusions for a robust interpretation.

Auditing and monitoring of the study is carried out according to international GCP guidelines. Central quality control ensures high quality data.

##### 4.3. Selection of trial population

In- and exclusion criteria are designed to select a group of patients with SAB that have a low-risk for SAB-related complications. Patients entering the study will have already received five to seven days of adequate intravenous antimicrobial therapy and have no signs and symptoms of complicated S. aureus infection prior to enrolment. Patients with a higher a priori risk for SAB-related complications are excluded (e.g. severe immunosuppression), or additional diagnostic steps to rule out deep seated infection are required as defined in section 4.3.2.

Reasons for gender distribution

SAB is more frequent in male patients (m:f = 2:1) (18). The reason for this phenomenon is not known. However, since most infections arise from the skin and nasal flora of the patient, this may reflect a higher nasal colonization rate in men (21, 22). However, mortality does not vary between male and femal gender (18). Gender specific differences in efficacy and safety of the used antimicrobial therapy are not expected.

##### 4.3.1. Inclusion criteria

- Age at least 18 years
- Not legally incapacitated
- Written informed consent from the trial subject has been obtained
- Blood culture positive for S. aureus not considered to represent contamination

- At least one negative follow-up blood culture obtained within 24-96 hours after the start of adequate antimicrobial therapy to rule out persistent bacteremia and absence of a blood culture positive for S. aureus at the same time or thereafter.
- Five to seven full days of appropriate i.v. antimicrobial therapy administered prior to randomization documented in the patient chart. Appropriate therapy has all of the following characteristics:
  - Antimicrobial therapy has to be initiated within 72h after the first positive blood culture was drawn.
  - Provided in-vitro susceptibility and adequate dosing (as judged by the principle investigator) preferred agents for pre-randomization antimicrobial therapy are: flucloxacillin, cloxacillin, vancomycin, and daptomycin. However, the following parenteral antimicrobials are allowed:
    - MSSA: penicillinase-resistant penicillins (e.g. flucloxacillin, cloxacillin), β-lactam plus β-lactamase-inhibitors (e.g. ampicillin+sulbactam, piperacillin+tazobactam), cephalosporins (except ceftazidime), carbapenems, clindamycin, fluoroquinolones, trimethoprim-sulfamethoxazole, doxycycline, tigecycline, vancomycin, teicoplanin, telavancin, linezolid, daptomycin, ceftaroline, ceftobiprole, and macrolides.
    - MRSA: vancomycin, teicoplanin, telavancin, fluoroquinolones, clindamycin, trimethoprim-sulfamethoxazole, doxycycline, tigecycline, linezolid, daptomycin, macrolides, ceftaroline, and ceftobiprole.

##### 4.3.2. Exclusion criteria

- Polymicrobial bloodstream infection, defined as isolation of pathogens other than S. aureus from a blood culture obtained in the time from two days prior to the first positive blood culture with S. aureus until randomization. Common skin contaminants (coagulase-negative staphylococci, diphteroids, Bacillus spp., and

Propionibacterium spp.) detected in one of several blood cultures will not be considered to represent polymicrobial infection

- Recent history (within 3 months) of prior S. aureus bloodstream infection
- In vitro resistance of S. aureus to all oral or all i.v. study drugs
- Contraindications in reference document for all oral or all i.v. study drugs
- Previously planned treatment with active drug against S. aureus during intervention phase (e.g. cotrimoxazol prohylaxis)
- Signs and symptoms of complicated SAB as judged by an ID physician. Complicated infection is defined as at least one of the following:
    - deep-seated focus: e.g. endocarditis, pneumonia, undrained abscess, empyema, and osteomyelitis
    - septic shock, as defined by the AACP criteria (23), within 4 days before randomization
    - prolonged bacteremia: positive follow-up blood culture more than 72h after the start of adequate antimicrobial therapy
    - body temperature >38 °C on two separate days within 48h before randomization
- Presence of the following non-removable foreign bodies (if not removed 2 days or more before randomization):
    - prosthetic heart valve
    - deep-seated vascular graft with foreign material (e.g. PTFE or dacron graft). Hemodialysis shunts are not considered deep-seated vascular grafts (s. below).
    - ventriculo-atrial shunt
- Presence of a prosthetic joint (if not removed 2 days or more before randomization). This is **not** an exclusion criterion, if all of the following conditions are fulfilled:

  - prosthetic joint was implanted at least 6 months prior, *and*
  - catheter-related infection, skin and soft tissue infection, or surgical wound infection is present (as defined below), *and*
  - joint infection unlikely (no clinical or imaging signs)

- Presence of a pacemaker or an automated implantable cardioverter defibrillator (AICD) device (if not removed 2 days or more before randomization). This is **not** an exclusion criterion, if all of the following conditions are fulfilled:
  - pacemaker or AICD was implanted at least 6 months prior, *and*
  - catheter-related infection, skin and soft tissue infection, or surgical wound infection is present (as defined below), *and*
  - no clinical signs of infective endocarditis, *and*
  - infective endocarditis unlikely by echocardiography (preferably TEE), *and*
  - pocket infection unlikely (no clinical or imaging signs)
- Failure to remove any intravascular catheter which was present when first positive blood culture was drawn within 4 days of the first positive blood culture
- Severe liver disease. This is **not** an exclusion criterion, if the following condition is fulfilled:
  - catheter-related infection, skin and soft tissue infection, or surgical wound infection is present (as defined below)
- End-stage renal disease. This is **not** an exclusion criterion, if all of the following conditions are fulfilled:
  - catheter-related infection, skin and soft tissue infection, or surgical wound infection is present (as defined below), *and*
  - no clinical signs of infective endocarditis, *and*
  - infective endocarditis unlikely by echocardiography (preferably TEE), *and*

  - in patients with a hemodialysis shunt with a non-removable foreign body (e.g. synthetic PTFE loop): no clinical signs of a shunt infection
- Severe immunodeficiency
  - primary immunodeficiency disorders
  - neutropenia (<500 neutrophils/µl) at randomization or neutropenia expected during intervention phase due to immunosuppressive treatment
  - uncontrolled disease in HIV-positive patients
  - high-dose steroid therapy (>1 mg/kg prednisone or equivalent doses given for >4 weeks or planned during intervention)
  - immunosuppressive combination therapy with two or more drugs with different mode of action
  - hematopoietic stem cell transplantation within the past 6 months or planned during treatment period
  - solid organ transplant
  - treatment with biologicals within the previous year
- Life expectancy < 3 months
- Inability to take oral drugs
- Injection drug user
- Expected low compliance with drug regimen
- Participation in other interventional trials within the previous three months or ongoing
- Pregnant women and nursing mothers
- For premenopausal women: Failure to use highly-effective contraceptive methods for 1 month after receiving study drug. The following contraceptive methods with a Pearl Index lower than 1% are regarded as highly-effective:
  - oral hormonal contraception ('pill')

- dermal hormonal contraception
- vaginal hormonal contraception (NuvaRing®)
- contraceptive plaster
- long-acting injectable contraceptives
- implants that release progesterone (Implanon®)
- tubal ligation (female sterilisation)
- intrauterine devices that release hormones (hormone spiral)
- double barrier methods

This means that the following are not regarded as safe: condom plus spermicide, simple barrier methods (vaginal pessaries, condom, female condoms), copper spirals, the rhythm method, basal temperature method, and the withdrawal method (coitus interruptus). Due to possible interactions and side effects of the study medication (applies to cotrimoxazol, clindamycin, and flucloxacillin), hormonal contraception may not be safe and another highly-effective contraceptive method needs to be employed.

- Persons with any kind of dependency on the investigator or employed by the sponsor or investigator
- Persons held in an institution by legal or official order

Definition of catheter-related infection (for use in exclusion criteria):

- The same S. aureus isolate (based on antibiotic susceptibility) is present in the positive blood culture and in the catheter tip culture, *or*
- The same S. aureus isolate is present in the positive blood culture and in pus or skin swab from the catheter exit site, *or*
- Two initial blood cultures positive for S. aureus exhibit a positive differential time to positivity and there is no other plausible source of infection, *or*

- Clinically strongly expected catheter-related infection: e.g. pus/reddening/pain at exit site, or shivers during infusion and no other plausible cause of infection.

Definition of skin and soft tissue infection (for use in exclusion criteria):

- S. aureus in wound swab, *or*
- Clinical signs of skin and soft tissue infection (abscess, thrombophlebitis, furuncle, etc.) and no signs of any other infective focus

Definition of surgical wound infection (for use in exclusion criteria):

- S. aureus in wound swab, *or*
- Clinical signs of an infected wound and no signs of any other infective focus

##### 4.4. Withdrawal of trial subjects after trial start

A patient may discontinue from the SABATO trial at any time for any reason. It is the right and the duty of the investigator to stop or modify the study treatment in any case the risk/benefit ratio is unacceptable to the individual subject or due to unmanageable factors that may interfere significantly with the study procedures and/or the interpretation of the results. However, individual circumstances will be carefully documented (see below).

###### 4.4.1. Procedures for premature withdrawal from treatment or trial

A premature withdrawl occurs when an enrolled patient ceases participation in the study prior to the completion of the protocol (e.g. withdrawn consent). All patients prematurely discontinuing from the study, regardless of cause, must receive a final evaluation at the time of withdrawal, preferably by a study visit. Premature withdrawal is considered a protocol violation and patients may be classified as non-evaluable (to be decided upon by the masked Clinical Review Committee (CRC)). All data collected until this point of time will be stored according to AMG §40, 2a, 3. Patients will not be replaced. If possible, EOS and EOT data should be collected from each patient (this is relevant for the ITT analysis)

The reason(s) for early discontinuation should be documented in the study termination record of the eCRF. If a patient withdrawing from the study has an ongoing AE at the time of withdrawal, the AE will be followed for 72 hours after the last dose of study therapy received; an ongoing SAE will be followed until resolved (recovery or death). In case a patient is withdrawn from the study before the first dose of study drug (e.g. due to withdrawn informed consent, or a late positive follow-up blood culture), baseline information will be collected, but follow-up visits are not performed.

#### 4.5. Closure of trial sites/Premature termination of the clinical trial

##### 4.5.1. Closure of trial sites

The Steering Committee shall have the right to terminate the study at individual trial sites at its discretion with written notice to the institution and the principle investigator. The individual principle investigator or institution shall have the right to terminate this study at its discretion with written notice to the Steering Committee. The sponsor has the right to terminate or suspend the trial prematurely in a trial site, if there are any relevant medical or ethical concerns, or if completing the trial is no longer practicable.

Possible reasons for closure of trial sites include, but are not limited to:

- Excessive delay at trial start
- Unsatisfactory enrolment with respect to quantity (<3 patients/year) or quality
- Inaccurate or incomplete data collection
- Falsification of records
- Failure to adhere to the protocol
- At the request of DFG, local IRB, DMC, or local authorities

In case a local IRB suspends or terminates the study at their site, it will not impact the status of the study at other sites, if the reason for suspension or termination is specific to that site.

##### 4.5.2. Premature termination of trial

The Steering Committee (SC) under the leadership of the PCI has the right to terminate the trial prematurely after appropriate recommendations from the DMC, and any statistician the SC seeks advice from. If such action is taken, the reasons for terminating the trial must be documented in detail. All trial subjects still under treatment at the time of termination are asked to undergo a final examination which must be fully documented. The PCI must be informed without delay if any principle investigator has ethical concerns about continuation of the trial.

Premature termination of the trial will be considered if:

- The risk-benefit balance for the trial subject changes markedly
- It is no longer ethical to continue treatment with the study drug
- The sponsor considers that the trial must be discontinued for safety or efficacy reasons (e.g. on the advice of the DMC)
- It is no longer practicable to complete the trial

#### 4.6. Treatment

##### 4.6.1. Treatments to be given

This study is an open-label study that uses standard doses of medication. Therefore, the study drugs are not centrally distributed, repackaged, or labelled. All antimicrobial treatments are commercially available antimicrobials approved by the respective national authorities.

Depending on the susceptibility of the isolates, expected drug interactions, contraindications and expected side effects, the drug is chosen from the corresponding list below. The drugs of first choice must be administered unless allergy or intolerance is suspected or resistance of the S. aureus isolate has been demonstrated.

**OST (oral administration)**

| MSSA | MRSA |
| --- | --- |
| 1st choice:Trimethoprim/sulfamethoxazole p.o. | 1st choice: Trimethoprim/sulfamethoxazole p.o. |
| 2nd choice: Clindamycin p.o. | 2nd choice: Linezolid p.o. |

**IST (intravenous administration)**

| MSSA | MRSA |
| --- | --- |
| 1st choice: Flucloxacillin i.v. (Cloxacillin in Spain & France) or Cefazolin i.v. | 1st choice: Vancomycin i.v. |
| 2nd choice: Vancomycin i.v. | 2nd choice: Daptomycin i.v. |

All listed i.v. study drugs (flucloxacillin, cloxacillin, cefazolin, vancomycin, daptomycin) are current standard treatment (6, 8). However, all intravenous and oral antimicrobials, with the exception of daptomycin and cefazolin, are being used off-label in SAB.

Regarding the oral study drug, there is some evidence for equal safety and efficacy. All oral study drugs are known to have excellent bioavailability and a good safety profile.They further provide adequate tissue concentrations needed to treat S. aureus infections and rates of resistance to trimethoprim-sulfamethoxazole and linezolid, are low.

The best evidence for the equivalence of oral formulations with standard therapy in SAB is available for trimethoprim-sulfamethoxazole and linezolid. Trimethoprim-sulfamethoxazole is in use for several decades, shows good activity against S. aureus (37), and synergistic effects in vitro and in vivo (38). In a retrospective analysis of SAB patients treated with oral or i.v. trimethoprim-sulfamethoxazole, safety and efficacy was judged to be similar to vancomycin (38). In a RCT involving 101 i.v. drug users with severe S. aureus infections, trimethoprin-sulfamethoxazole performed comparable to vancomycin in the subset of MRSA patients (39). A pooled post-hoc analysis of five RCTs that compared linezolid to standard vancomycin therapy in MRSA bacteremia found no difference between linezolid and vancomycin (40). In these studies, linezolid could be switched to oral administration after 7 days of i.v. treatment. Unfortunately, no data comparing oral and i.v. treatment have been published.

Evidence for the safety of oral antimicrobial therapy is also available from a few other clinical studies: Data from a randomized controlled trial on right-sided S. aureusendocarditis in 44 injection drug users showed that oral ciprofloxacin plus rifampicin was equally effective and associated with less drug toxicity than i.v. oxacillin or vancomycin therapy (41). In another RCT involving 104 patients with deep-seated S. aureus infection including SAB, a combination of oral fleroxacin plus rifampicin was compared to standard i.v. therapy (17). Cure rates were similar, and although underpowered, oral therapy was considered an effective alternative to standard parenteral therapy leading to a significant reduction in length of hospital stay by 11 days. In another retrospective study, clindamycin proved to be effective in children with invasive S. aureus infections (42). Oral β-lactams in contrast, are not considered adequate for treatment of invasive S. aureusinfections due to their unfavorable bioavailability and dose-limiting gastrointestinal toxicity (43). Further circumstantial evidence for the safety and efficacy of oral formulations can be gathered from trials concerning other types of infections. Oral versus i.v. quinolones (ciprofloxacin) were found equally effective for serious infections of various origins in a RCT involving 105 patients (44). Oral linezolid was effective and safe in the treatment of complicated skin and skin structure infections (45). A post-hoc analysis of data from two RCT in patients with community-acquired pneumonia (CAP) showed that oral therapy with moxifloxacin was safe and effective (46).

###### 4.6.2. Description of study drugs

Study drugs will be as per marketed drug available from the local hospital pharmacy stock. Therefore, study drug may differ between participating hospitals. In accordance with regulations, one reference document for each active substance has been selected. Reference documents for all study drugs are listed in Appendix 11.8. For oral study drugs all orally available formulations (suspensions, tablets) can be used.

###### *4.6.2.1. Manufacture of the study drugs*

Formulation, packaging and labeling of study drugs will be as per marketed drug and available from the local hospital pharmacy stock.

###### *4.6.2.2. Labelling of study drugs*

Labelling of study drugs will be as per marketed drug.

###### *4.6.2.3. Storage and reconstitution of study drugs*

Storage conditions are as specified by the manufacturer. Unreconstituted vancomyin should be stored below 25°C. Daptomycin should be stored at 4-8°C. Flucloxacillin, cloxacillin, cefazolin, linezolid, trimethoprim-sulfamethoxazol, and clindamycin can be stored at room temperature. Cefazolin should be protected from light in its unreconstituted and reconstituted form. All intravenous formulations will be reconstituted according to the manufacturer's instruction under the supervision of a physician. All reconstituted intravenous formulations should be immediately used or stored at 4°C for a maximum of 24h.

##### 4.6.3. Compliance with treatment / Dispensing and return of study drugs

Oral and intravenous study drugs will be accounted for and documented by the study physician. Package inserts of study drugs will be archived in the patient chart and the Investigator Site File. If package inserts are not available,, the batch number and brand name of the study drug can be documented in the patient chart and Investigator site file. Compliance of outpatient treatment (OST or IST on OPAT) will be assessed by telephone contacts every two to three days.

##### 4.6.4. Assignment of trial subjects to treatment groups

Patients are randomly allocated to treatment arms (1:1) not earlier than one day before starting study drug. This is achieved by a central 24-7 Internet randomization service ALEA (stratified by study center, permuted blocks of varying length). Authorized local study staff may login to a secure website, randomize a patient and receive an email with attached pdf giving all the details on the allocated treatment. The randomization service is set up and maintained by IMSIE, University of Cologne.

###### 4.6.5. Dose selection for study drugs

The study drug is administered according to standard dosing schemes (s. table 2).

**Table 2: Dosing of study drug**

| **OST** | Minimum daily dose | Suggested regimen | Acceptable dosing | Dose adjustment |
|---|---|---|---|---|
| Trimethoprim-sulfamethoxazole | 320/1600 mg | 160/800mg twice a day | | Severe renal impairment |
| Clindamycin | 1800 mg | 600mg three times a day | | No |
| Linezolid | 1200 mg | 600mg twice a day | | No |
| **IST** | | | | |
| Flucloxacillin | 6 g (in at least 4 doses a day, or continuous infusion) | 2g four times a day | 4g three times a day | Severe renal impairment |
| Cloxacillin | 6 g (in at least 4 doses a day, or continuous infusion) | 2g four times a day | 2g six times a day | No |
| Cefazolin | 1g three times a day | 2g three times a day | 3g four times a day | Renal impairment |
| Vancomycin | as determined by TDM | 1g twice a day | Loading dose and continuous infusion are acceptable | TDM |
| Daptomycin | 6 mg/kg once per day | 6-10 mg/kg once per day | | Renal impairment |

##### 4.6.6. Time of administration and dose adjustments of study drugs in the individual trial subject

Dose adjustments in individual patients will be performed as judged appropriate by the principle investigator for weight, renal clearance, and (in case of vancomycin) therapeutic drug monitoring. Guidance on how to adjust dosage is provided below (tables 3 to 6); dose adjustments for patients undergoing dialysis are detailed in the appropriate reference document (Appendix 11.8). The reason why a dose adjustment is performed needs to be documented.

In both arms, the preferable study drug is the the first choice regimen. The alternative regimen can be used when S. aureus is resistant to the first choice, contraindications are present, and intolerance or severe side effects are expected. Study drug can also be switched during therapy from first choice to the respective alternative medication when intolerance (e.g. allergy), contraindication or severe side effects (e.g. Lyell syndrome) arise.

If a contraindication (resistance, known allergy, etc.) for the alternative drug arises during treatment and the drug of first choice cannot be used the study drug is discontinued and the principle investigator decides about further antimicrobial therapy. Early discontinuation of study drug or combination therapy (e.g. addition of gentamicin, rifampicin, fosfomycin) is regarded as a protocol violation.

**Table 3: Suggested dose adjustments for trimethoprim-sulfamethoxazole**

| Glomerular filtration rate (ml/min) | Dose adjustment |
|---|---|
| Above 30 | No adjustment |
| 15-30 | 160/800mg once a day |
| Below 15 | contraindicated |

**Table 4: Suggested dose adjustments for Flucloxacillin**

| Glomerular filtration rate ml/min | Dose 70 kg | Dose 100 kg |
|---|---|---|
| 18 | 1.5g four times a day | 2g four times a day |
| 8 | 1.5g three times a day | 2g three times a day |
| 2 | 1g three times a day | 1.5g three times a day |
| 0.5 | 2g once a day | 3g once a day |

**Table 5: Suggested dose adjustment for cefazolin**

| Glomerular filtration rate ml/min | Dose |
|---|---|
| 35-54 | 2g three times a day |
| 10-34 | 1g two times a day |
| <10 | 1g every 18-24h |

**Table 6: Suggested dose adjustment for vancomycin**

| Glomerular filtration rate ml/min | Dose | Dose 70 kg |
|---|---|---|
| >50 ml/min | 15-20 mg/kg two to three times a day | 1g-1,5g two to three times a day |
| 20-49 | 15-20 mg/kg once a day | 1g-1,5g once a day |
| <20 | Guided by TDM | Guided by TDM |

**Table 7: Suggested dose adjustment for daptomycin**

| Glomerular filtration rate ml/min | Dose |
|---|---|
| ≥30 | No adjustment necessary |
| <30 | 6-10 mg/kg every 48h |

Therapeutic drug monitoring (TDM) for vancomycin is carried out according to local standard procedures. However, monitoring vancomycin trough levels once weekly (goal: 10-20 µg/ml) starting prior to the fourth dose, is encouraged. For vancomycin, a loading dose can be used for seriously ill patients (25-30 mg/kg) to rapidly achieve target concentration. Intravenous dosing should be based on actual body weight (15-20 mg/kg/dose 2-3 times a day) and subsequent dosing should be adjusted based on serum trough levels. In the critically ill patient with renal impairment an initial loading dose of 25-30 mg/kg should not be reduced.

##### 4.6.7. Blinding

This study compares intravenous and oral treatment regimens. Neither patients nor investigators can be fully masked regarding treatment. However, clinical response to treatment will be evaluated in a blind manner by the Clinical Review Committee (CRC). Moreover, the Data Monitoring Committee (DMC) will be masked, though unblinding may be requested.

###### *4.6.7.1. Unblinding*

Not applicable.

##### 4.6.8. Previous and concomitant medication

There is no restriction on previous medication. Concomitant medication must be used in accordance with the Summary of Product Caracteristics, avoiding the initiation of any medication with a potential to interact with the study drug.

Administration of oral or intravenous antimicrobials will be recorded retrospectively for 10 days prior to the first dose of study drug, during the intervention, and during follow-up. All other medication will not be recorded in the eCRF.

During the intervention phase, systemic antimicrobial agents with activity against Staphylococcus aureus other than study drug are not permitted for targeted therapy in both arms. The addition of such an antimicrobial agent during the intervention or follow-up phase will be considered a potential protocol violation. Its significance has to be judged by the Clinical Review Committee (CRC).

###### 4.6.9. Rescue therapy for emergencies

When a relapsing SAB or deep-seated infection with S. aureus occurs as a complication during the intervention or follow-up phase, the endpoint is reached. Treatment will then be instituted according to the standard of care at each study site. Most likely, patients on oral study drug will then receive i.v. treatment. Patients on intravenous study drug may receive another antimicrobial agent. Furthermore, a longer course of antimicrobial therapy, addition of another antimicrobial agent, or a surgical intervention may be necessary.

###### 4.6.10. Continuation of treatment after the end of the clinical trial

Study drug is discontinued at EOT without special measures.

##### 4.7. Efficacy and safety variables

The **primary endpoint** measure, SAB-related complication, reflects the failure rate of antimicrobial therapy in preventing late complications. This includes relapsing SAB, deep-seated S. aureus infection, and attributable mortality within 90 days and is thus the most appropriate clinical outcome measure.

Microbiological success is sometimes demonstrated by a negative blood culture as a test of cure at EOT. Since patients in this trial have already been treated for seven days with antimicrobials before randomization, almost all blood cultures obtained at EOT are expected to yield a negative result. Therefore, microbiological success has not been chosen as an endpoint.

Death unrelated to SAB is expected at about 5% within 30 days. It was not included in the primary endpoint because this would compromise the power of the trial by variance inflation. However, SAB-related and all-cause mortality will be carefully assessed and compared (secondary/safety endpoints).

The **secondary endpoint**, length of hospital stay, reflects the potential benefits for patients who have been switched to oral medication. Furthermore, 14- and 30-day survival and complications related to i.v. therapy, e.g. chemical or septic (thrombo-)phlebitis will be measured.

The **safety** of study drugs is assessed by monitoring Clostridium difficile associated diarrhea (CDAD), AEs and SAEs.

##### 4.7.1. Measurement of efficacy and safety variables

*4.7.1.1. Primary target variable*

All data will be obtained at the study visits or telephone contacts and are based on the assessment of the study physician, patient interviews, laboratory reports, and chart data. After discharge, patients are encouraged to report to the study center any changes in health (e.g. adverse events). Study site staff will follow up on reported issues.

The primary endpoint, **SAB-related complication**, defined as relapsing SAB or deep-seated infection, will be derived from laboratory and clinical reports. Patients with either condition will be classified as “failure”. To ensure a high quality of data, late complications need to be assessed by the principle investigator. SAB-related complications are classified as either “microbiologically documented” or “clinically suspected”. All cases of SAB-related complication are carefully evaluated for plausibility by the masked Clinical Review Committee (CRC) on the basis of clinical symptoms, vital signs, laboratory parameters, the assessment of the study physician, patient interviews and chart data. The CRC will be provided with further information by the principle investigator as needed.

To qualify for a “microbiologically documented” relapsing SAB or deep-seated infection, the S. aureus isolate needs to exhibit the same characteristics as the original infecting isolate (based on antimicrobial susceptibility and genotyping tests as appropriate). Furthermore, the isolated strain needs to be judged not to represent a contaminant by the local investigator.

**Relapsing SAB** is defined as positive blood culture for S. aureus within the intervention or follow-up period. During the follow-up phase blood cultures will be taken according to standard of care at the local site, when a bloodstream infection is clinically suspected. Since every blood culture carries the risk of contamination, study sites are encouraged to draw at least two blood cultures, when clinically indicated.

Proven catheter-related S. aureus bloodstream infections during the follow-up period are not considered relapsing SAB, since they are highly likely to result from a new infection. Catheter-related blood-stream infection is considered “proven”, when:

- The same S. aureus isolate is present in the positive blood culture and in the catheter tip culture, or
- The same S. aureus isolate is present in the positive blood culture and in pus or skin swab from the catheter exit site, or
- Two initial bloodcultures positive for S. aureus exhibit a positive differential time to positivity and there is no other plausible source of infection.

**Deep-seated infection** is any deep-seated focus of S. aureus infection resulting from hematogenous dissemination. Diagnosis requires either a positive culture from the respective site, or a blood culture positive with S. aureus plus imaging studies showing the presumed focus.

Deep-seated foci consist of, but are not limited to:

- Infective endocarditis, judged by modified Duke criteria (22)
- Vertebral and non-vertebral osteomyelitis
- Suppurative arthritis
- Spinal empyema
- Muscle abscess (e.g. psoas abscess)
- Meningitis, brain abscess
- Lung abscess
- Visceral abscess (kidney, liver, spleen, etc.)

Catheter-related infections, superficial skin-and soft tissue infections such as thrombophlebitis, or superficial wound infections do not qualify as “deep-seated”, since they are likely to result from a new infection.

There is a possibility that late complications of SAB may be overlooked. By educating the patient about signs and symptoms of potential late complications, we expect that a diagnostic work-up will be performed in nearly all suspected cases. In the case of a suspected complication, the patient’s current care-providers will be contacted by the principle investigator for relevant clinical information and lab reports.

*4.7.1.2. Secondary and other target variables*

The **length of hospital stay** is defined as the number of days a patient spends in the hospital from randomization to discharge. When a patient is transferred to another hospital, days spent at the other hospital are included.

**Survival** will be assessed during the hospital stay and at the follow-up telephone interviews. Death will be attributed to SAB when at least one of the following conditions is present [16]:

- positive blood culture for S. aureus drawn within 72h before death
- persistent focus of deep-seated S. aureus infection at time of death
- persistent signs and symptoms of systemic infection at time of death as judged by study physician
- post-mortem analysis proving S. aureus related complication as cause of death

All other causes will be classified as unrelated to SAB.

**Complications of i.v. therapy** will be assessed from chart data, assessment of the study physician and from the patient interview. Complications may include, but are not limited to:

- local complications, such as: infiltration, extravasation, hematoma, phlebitis, thrombosis, thrombophlebitis, or infection at catheter insertion site

- systemic complications, such as: embolism, systemic infection, circulatory overload, allergic reaction
- in vancomycin therapy: “red man syndrome” due to inappropriately fast i.v. administration
- any other complication felt to be due to intravenous therapy by the principle investigator

*4.7.1.3. Safety analysis*

In patients that report diarrhea during the intervention or follow-up phase, testing for **Clostridium difficile associated diarrhea (CDAD)** is not mandatory and will be performed according to the standard of care at the respective trial site. Furthermore, AEs and SAEs are collected until EOS.

*4.7.1.4. Description of visits*

An overview of the visits is provided in a study flow chart and the visit time schedule (s. table 3 and 4).

Screening

Principle investigators will follow local procedures to obtain permission to monitor blood culture results daily. In patients with SAB, eligibility will be assessed by reviewing in/exclusion criteria against patient charts, laboratory results and by performing patient interviews.

During in-person visits, a member of the local study team assesses patient history, clinical and laboratory data, as well as imaging studies to assess the risk of late complications and verify in- and exclusion criteria (e.g. duration of appropriate antimicrobial therapy, presence of deep-seated infection or prosthetic devices, catheter removal, negative follow-up blood cultures). Diagnostic procedures related to assessing patients with SAB during the screening phase (laboratory tests, blood cultures, imaging studies for assessment of deep-seated infections) are standard of care in all participating hospitals. The following laboratory results are standard of care and will be documented once within 3 days before randomization (Day -3 to 1) to establish baseline values for liver, kidney, bone marrow, and muscle function, and the inflammatory response: hemoglobin, white blood count (WBC), red blood count (RBC), platelets, sodium, potassium, creatinine, prothrombin time (PT), partial thromboplastin time (aPTT), alanine aminotransferase (ALAT), aspartate transaminase (ASAT), bilirubin, C-reactive protein (CRP), and creatine-phosphokinase (CPK).

Written informed consent must be obtained from each patient before data is entered into the study database. Only patients who meet all inclusion criteria and none of the exclusion criteria will be randomized to receive either OST or IST.

Data on patients screened will be entered anonymously in a separate database that is not linked to the study database. The following data will be recorded for each screened patient: age, month and year of positive blood culture, medical specialty, and key exclusion critera.

Intervention visits

The **baseline visit** (first visit after randomization) will be performed after the patient has signed informed consent. It takes place on the day when the first study medication is given or on the day before. At the baseline visit a physical examination is performed and vital signs are taken. Furthermore, SAB-related complications, current medication, adverse events, Clostridium difficile associated diarrhea (CDAD), and complications of i.v. therapy are assessed.

The **EOT** visit will be performed within 2 days after the last dose of study drug. Current medication, SAB-related complications, adverse events, CDAD, and complications of i.v. therapy are assessed. If patients are discharged during the intervention phase (possible for OST or OPAT), patients will be contacted every two to three days by site staff by telephone and asked about study drug administration, concomitant medications and adverse events. If there is the suspicion of deteriorating liver, kidney or bone-marrow function, the patient will be asked to have blood tests performed at the study center.

Interim visits are performed at the discretion of the principle investigator.

Follow-up visit 1 (day 25-39) and EOS visit (day 85-99)

The Follow-up visit 1 (FU1) and EOS visits serve to collect data for the primary and secondary endpoints, current medication, and adverse events and can be conducted by phone or in clinic. The EOS evaluation will be the final assessment of “failure”.

If a patient is readmitted to any hospital before final assessment, the patient or family member should notify study staff of the reason for admission.

Early termination visit

If the patient terminates the study early for any reason, the final assessment should be performed at time of termination. However, every effort will be made to receive data at the final assessment (EOS).

Duration of the clinical trial in the individual subject

The final assessment of endpoints will be 85-99 days after the first dose of study drug.

**Table 8: Study flow chart of the clinical trial**

| | Screening | Treatment | | FU1 & EOS |
|---|---|---|---|---|
| **Visit number** | **0** | **1** | **2 *** | **3, 4 *** |
| **Day** | **-5 to-1** | **-1 to 1** | **7-11** (EOT)** | **25-39 & 85-99** |
| Informed consent | X | | | |
| Check in/exclusion criteria | X | | | |
| Randomization | X | | | |
| Demographic data | X | | | |
| Medical history | X | | | |
| Charlson score | X | | | |
| Pitt bacteremia score | X | | | |
| Current medication | X | X*** | X | X |
| Infective focus | X | | | |
| **Clinical data** | | | | |
| Physical examination | X | X | | |
| Vital signs | X | X | | |
| **Outcome assessment** | | | | |
| SAB-related complications | | X | X | X |
| Length of stay in ICU and in hospital (days) | | | X | X |
| 90-day mortality | | | | X |
| **Safety** | | | | |
| Adverse events | | X | X | X |
| CDAD | | X | X | X |
| Complications of iv therapy | | X | X | X |
| **Laboratory data** | | | | |
| The following routine laboratory results are documented once from day -3 to 1, if available: Hemoglobin, RBC, WBC, platelets, Na, K, creatinine, PT, aPTT, ALAT, ASAT, bilirubin, CPK, CRP, blood culture | X | | | |
| Pregnancy test in premenopausal women | X | | | |

* **FU1,** EOT and EOS visits may be telephone contacts

** Patients that are discharged early will be followed by telephone contact every other day until EOT.

*** Administration of oral or intravenous antimicrobials will be recorded retrospectively for 10 days prior to the first dose of study drug, during the intervention, and during follow-up. All other medication will not be recorded in the eCRF.

**Table 9: Visit schedule**

| Visit | Trial day | First day possible | Last day possible | Comments |
|---|---|---|---|---|
| 0 | 0 | -5 | -1 | Screening |
| 1 | 1 | -1 | 1 | Baseline intervention visit |
| 2 | 7 | 7 | 11 | EOT (within 2 days after last study medication) |
| 3 | 25 | 25 | 39 | Follow-up visit 1 (FU1) |
| 4 | 85 | 85 | 99 | End of study (telephone contact) |

##### 4.7.2. Rationale for assessment procedures

Efficacy

The primary endpoint, development of SAB-related complications, will be assessed by follow-up telephone or in-person interviews. This has been done in similar studies (23) and is the most economic way to assess the primary endpoint.

Since no mandatory in-person visit will be done during follow-up, there is the possibility that SAB-related complications may be overlooked. However, untreated complications will be clinically overt after some time. By educating the patient about signs and symptoms, we expect that a diagnostic work-up will be initiated in nearly all cases. Cases may still be overlooked when patients die before appropriate diagnostic testing can be performed, the impact of which on results will be assessed by sensitivity analysis.

Safety

All study drugs have been on the market for years and are considered safe. In case of a suspected adverse event, e.g. fever, fatigue, pain, or any other complaints, patients will be asked to consult the local study site immediately or seek emergency treatment. Furthermore, information on AEs that were not reported by the patient is collected at EOS.

##### 4.7.3. Pharmacokinetics/Determination of drug levels

Pharmacokinetic measurements will not be part of the trial. Since there is no consensus on drug-level guided antimicrobial therapy, determining vancomycin through levels is encouraged, but levels are not recorded in this study.

#### 4.8. Data quality assurance

##### 4.8.1. Monitoring

The trial sites will be monitored to ensure the quality of the data collected. The objectives of the monitoring procedures are to ensure that the trial subject's safety and rights as a study participant are respected, that accurate, valid and complete data are collected, and that the trial is conducted in accordance with the trial protocol, the principles of GCP and local legislation.

The monitoring is based on the adaptive on-site monitoring strategy developed by the ADAMON project (36), complemented with central quality assurance measures. The necessary amount of on-site monitoring is deducted from the checklist for risk assessment provided by the ADAMON protocol: Due to the fact that the given medication is established and is intended to be given according to the SmPC, the class "K3" for on-site monitoring has been determined. The class K3 requires one on-site visit for each trial site early in the trial. Further visits are planned if the on-site visit or central quality measures reveal that the trial is not performed in accordance with GCP or local legislation. Additionally, the site will be monitored by central monitoring activities. Parameters that are monitored by central monitoring are e.g. timeliness of data entry, missing documentation, number of patients lost to follow up, number of occurring AEs and SAEs.

All investigators agree that the monitor regularly visits the trial site and assure that the monitor will receive appropriate support in their activities at the trial site, as agreed in separate contracts with each trial site. The declaration of informed consent (see Section 5.4) includes a statement to the effect that the monitor has the right – while observing the provisions of data protection legislation – to compare the electronic case report forms (eCRFs) with the trial subject's medical records (doctor's notes, laboratory printouts etc.).

The investigator will secure access for the monitor to all necessary documentation for trial-related monitoring. The exact extent of the monitoring procedures is described in a separate monitoring manual. The aims of the monitoring visits are as follows:

- To check the declarations of informed consent
- To monitor trial subject safety (occurrence and documentation/reporting of AEs and SAEs)
- To check the completeness and accuracy of entries on the eCRFs
- To validate the entries on the eCRFs against those in the source documents (source data verification),
- To perform drug accountability checks
- To evaluate the progress of the trial
- To evaluate compliance with the trial protocol
- To assess whether the trial is being performed according to GCP at the trial site
- To discuss with the investigator aspects of trial conduct and any deficiencies found

A monitoring visit report is prepared for each visit describing the progress of the clinical trial and any problems (e.g. refusal to give access to documentation).

##### 4.8.2. Audits/Inspections

As part of quality assurance, the sponsor has the right to audit the trial sites and any other institutions involved in the trial. The aim of an audit is to verify the validity, accuracy and completeness of data, to establish the credibility of the clinical trial, and to check whether the trial subject’s rights and trial subject’s safety are being maintained. The sponsor may assign these activities to persons otherwise not involved in the trial (auditors). These persons are allowed access to all trial documentation (especially the trial protocol, case report forms, trial subjects’ medical records, drug accountability documentation, and trial-related correspondence).

The sponsor and all trial sites involved undertake to support auditors and inspections by the competent authorities at all times and to allow the persons charged with these duties access to the necessary original documentation.

All persons conducting audits undertake to keep all trial subject data and other trial data confidential.

##### 4.9. Documentation

All data relevant to the trial are documented soon after measurement by the investigator responsible in the electronic case report form supplied. Entering data may be delegated to members of the trial team. Entries regarding the primary endpoint, SAB-related complications, may be made only by the principle investigator. The eCRFs are signed by the principle investigator.

###### 4.9.1. Data management

The IT infrastructure and data management staff will be supplied by the ZKS Cologne. The trial database will be developed and validated before data entry based on standard operating procedures at the ZKS Cologne. The data management system is based on commercial trial software and stores the data in a database. All changes made to the data are documented in an audit trail. The trial software has a user and role concept that can be adjusted on a trial-specific basis. The database is integrated into a general IT infrastructure and safety concept with a firewall and backup system. The data are backed up daily. After completion and cleaning of data, the database is locked and the data exported for statistical analysis.

The data will be entered online at the trial sites via the Internet. Plausibility checks are run during data entry, thereby detecting many discrepancies immediately. The ZKS Cologne Data Management will conduct further checks for completeness and plausibility and will clarify any questions with the trial sites electronically via the trial software. These electronic queries have to be answered by the trial site without unreasonable delay. Additionally, central quality assurance located at the data management facility will provide regular reports to provide information to project management to identify centres that might benefit from additional quality assurance measures. Reports will provide e.g. center-based information regarding quality of eCRF documentation, query response time or missing data. Further details will be specified in the data management manual.

##### 4.9.2. Archiving

The eCRFs, informed consent forms and other important trial materials will be archived for at least 10 years in accordance with §13 Sec. 10 of the GCP Regulations. Trial subject identification lists at each trial site will be stored separately from trial documentation.

### 5. Ethical and regulatory aspects

#### 5.1. Independent ethics committee

The clinical trial will not be started before approval of the competent ethics committee.

In each trial site, the clinical study will not be started before approval of the competent local ethics committee concerning the suitability of the trial site and the qualifications of the investigators.

#### 5.2. Ethical basis for the clinical trial

The present trial protocol and any amendments were and will be prepared in accordance with the Declaration of Helsinki in the version of October 1996 (48th General Assembly of the World Medical Association, Somerset West, Republic of South Africa).

##### 5.2.1. Legislation and guidelines used for preparation

The present clinical trial will be conducted in accordance with the published principles of the guidelines for Good Clinical Practice (ICH-GCP) and applicable legislation (especially the Federal Drug Law [AMG] and the GCP-V). These principles cover, amongst other aspects, ethics committee procedures, the obtaining of informed consent from trial subjects, adherence to the trial protocol, administrative documentation, documentation regarding the study medication, data collection, trial subjects' medical records (source documents), documentation and reporting of adverse events (AEs), preparation for inspections and audits, and the archiving of trial documentation. All investigators and other staff directly concerned with the study will be informed that domestic and foreign supervisory bodies, the competent authorities and authorised representatives of the sponsor have the right to review trial documentation and the trial subjects' medical records at any time.

#### 5.3. Notification of the authorities, approval and registration

Before the start of the clinical trial, all necessary documentation will be submitted to the competent authorities for approval (Bundesinstitut für Arzneimittel und Medizinprodukte [BfArM], Agencia Espanola de Medicamentos y Productos Sanitarios [AEMyPS], Centrale Commissie Mendsgebondenen Onderzoek [CCMO], and Agence nationale de sécurité du medicament et des produits de santé [ANSM]). The respective regional authorities will also be notified.

The University Hospital of Cologne (Drittmittelverwaltung der Uniklinik Köln) and the University Hospital Düsseldorf (Drittmittelverwaltung der Uniklinik Düsseldorf) were informed that the trial is being conducted. Before the trial randomizes the first participant central items plus the protocol will be registered under Current Controlled Trials (www.controlled-trials.com) or another trial register approved by the World Health Organisation (WHO) (http://www.who.int/ictrp/en/).

#### 5.4. Obtaining informed consent from trial subjects

Trial subjects may not be enrolled into the present trial unless they have consented to take part in the trial after having been informed verbally and in writing in comprehensible language of the nature, scope and possible consequences by an authorized member of the study team Together with the consent to take part in the trial, the trial subject must also agree to representatives of the sponsor (e.g. monitors or auditors) or the supervisory authorities having access to the data recorded within the framework of the clinical trial. The trial subject will be informed of the potential benefit and possible side effects of the drugs. It must be clear to trial subjects that he or she can withdraw his or her consent at any time without giving reasons and without jeopardizing his / her further course of treatment.

The originally signed consent form is archived in the investigator site file. Trial subjects receive copies of the written information sheet, confirmation of insurance with conditions, and the signed informed consent form. A copy of the written information sheet and the signed informed consent form will be filed in the patient’s record.

The patient information sheet and informed consent form are supplied in Appendix 11.9.

The patient information sheet, informed consent form, all other documents handed out to the trial subject and any recruitment advertisements must be submitted for approval before use to the ethics committee. Part of the monitoring activities are to check that the most recent informed consent form was used before the trial subject was enrolled and that it was dated and signed by the trial subject himself or herself.

##### 5.5. Insurance of trial subjects

All trial subjects enrolled are insured in accordance with § 40 AMG under the group insurance contract of the Heinrich-Heine University Düsseldorf with Marsh Medical Consulting. The headquarters, policy number and telephone and fax number will be included in the patient information sheet.

##### 5.6. Data protection

The provisions of data protection legislation will be observed. It is assured by the sponsor that all investigational materials and data will be pseudonymised in accordance with data protection legislation before scientific processing.

Trial subjects will be informed that their pseudonymised data will be passed on in accordance with provisions for documentation and notification pursuant to § 12 and § 13 of the GCP Regulations to the recipients described there. Subjects who do not agree that the information may be passed on in this way will not be enrolled into the trial.

### 6. Statistical methods and sample size calculation

#### 6.1. Statistical and analytical plan

The essentials of the planned statistical analysis are given below. Further details are deferred to the Statistical Analysis Plan to be finalized before randomization of the first patient.

##### 6.1.1. Analysis sets

All analyses will be done on three study populations (s. figure 2):

The primary analysis set is derived from the per-protocol (PP) population. This dataset includes all study subjects who were essentially treated according to protocol and reached a defined endpoint in the trial (SAB-unrelated deaths will be excluded). The evaluability of study subjects will be assessed in a blind manner by the CRC.

The secondary analysis set is derived from the intention-to-treat (ITT) population. This dataset includes all randomized study subjects, analyzed as assigned, with indeterminate and missing outcomes counted as failures. Following current recommendations, CPMP/EWP/558/95 rev 2 (47) and CPMP/EWP/482/99 (49), the primary analysis is based on the per-protocol set; the analysis of the full analysis set (intention-to-treat, all randomized patients) will be of equal importance and should lead to similar conclusions for a robust interpretation.

The tertiary analysis set is the safety population. This dataset includes all study subjects who received any study drug.

**Figure 2:** Analysis sets

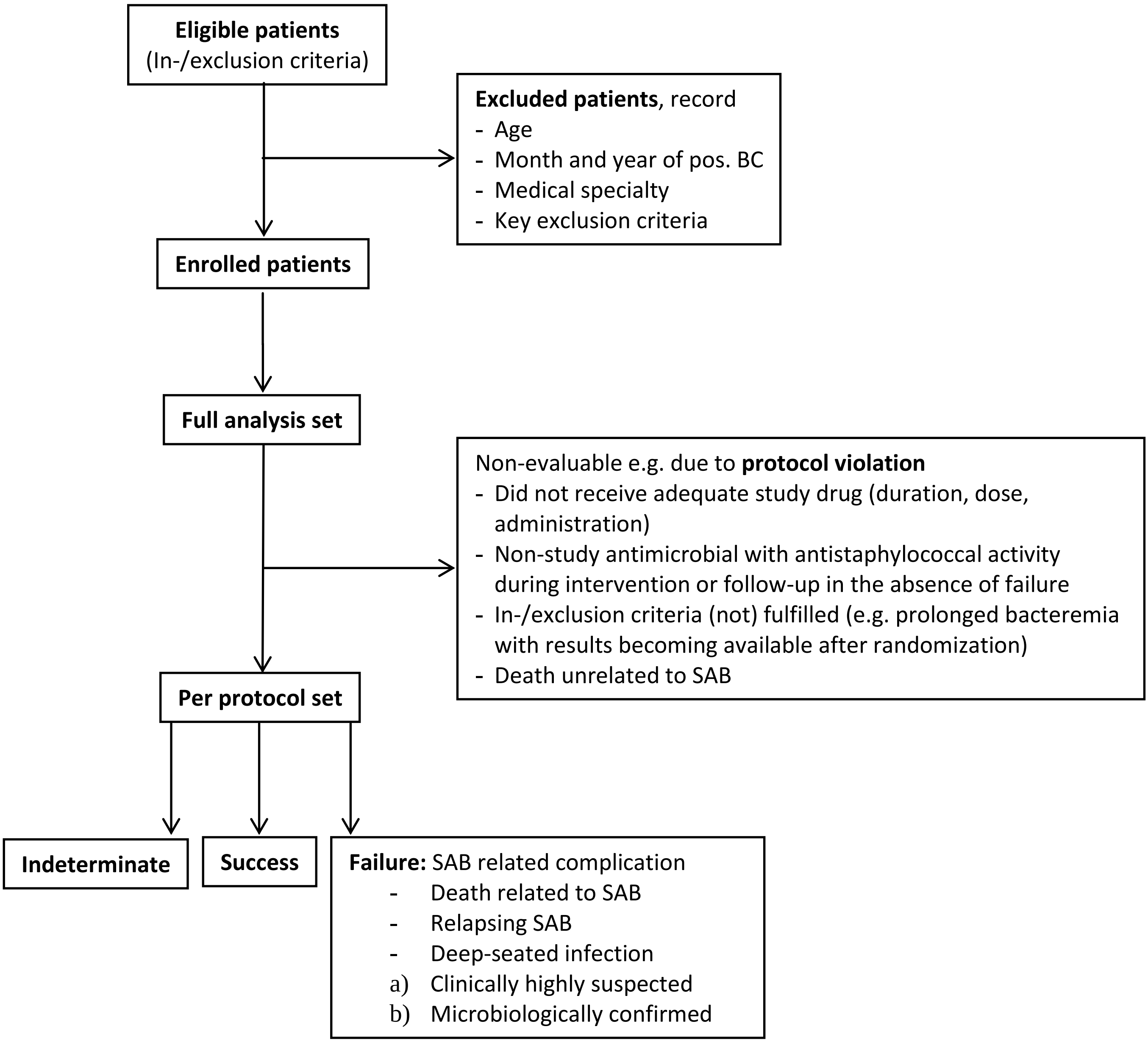

##### 6.1.2. Description of trial subject groups

Distributions of demographic and baseline characteristics (including age, sex, weight, transfer from another hospital, prior hospitalization during one year before randomization, length of hospital stay before randomization, surgery during the 6 months before randomization, infection during the 3 months before randomization, immunosuppressive therapy, underlying condition (24)) will be summarized by treatment group using valid count and either percentage (qualitative data) or mean, standard deviation and (0, 25, 50, 75, 100) percentiles (quantitative data).

##### 6.1.3. Primary target variable

The primary endpoint SAB-related complications (relapsing SAB or deep-seated infection with S. aureus) within 90 days will be evaluated regarding non-inferiority of oral vs. intravenous antimicrobial therapy by Zhao's test (test 1) of non-null hypothesis on proportions stratified by study center (25) at one-sided level 5% and with a non-inferiority margin of 10%. Thus, the hypotheses in terms of the proportion p of patients with SAB-related complications to be decided upon are

(A: null hypothesis) $H_0$: $p_{OST} > p_{IST} + 0.10$ vs

(A: alternative hypothesis) $H_a$: $p_{OST} \leq p_{IST} + 0.10$

If this null hypothesis can also be rejected (fixed sequence of hypotheses, thus no alpha-inflation), the above test (A) will be repeated at one-sided level 2.5%.

If this null hypothesis can be rejected (fixed sequence of hypotheses, thus no alpha-inflation), the non-inferiority margin of 5% will be applied, i.e.

(B: null hypothesis) $H_0$: $p_{OST} > p_{IST} + 0.05$ vs

(B: alternative hypothesis) $H_a$: $p_{OST} \leq p_{IST} + 0.05$

If this null hypothesis can also be rejected (fixed sequence of hypotheses, thus no alpha-inflation), the above test (B) will be repeated at one-sided level 2.5%.

Corresponding test-based confidence intervals will be calculated to aid interpretation.

The impact of missing data will be assessed in a sensitivity analysis by single/multiple imputation (based on logistic regression modelling).

##### 6.1.4. Secondary target variables

Secondary endpoints are evaluated by descriptive methods (by treatment group), generalised linear modelling, methods for rates, proportions and the time to event.

##### 6.1.5. Safety variables

AE/SAEs will be MedDRA coded and listed / summarized by treatment group, system organ class, preferred term, severity and relationship. Further safety variables (esp. laboratory data) will be listed / summarised by treatment group using valid count and either percentage (qualitative data) or mean, standard deviation and (0, 25, 50, 75, 100) percentiles (quantitative data).

##### 6.1.6. Subgroup analyses

Subgroup analysis will be performed by sex (male-female ration 3:2, study center and study drugs (see 4.6.1.) received as OST and IST, respectively.

##### 6.1.7. Interim analysis

An initially planned interim analysis will be converted into a final analysis, mainly due to difficulties in reaching the full sample size. Previously, an interim analysis was planned in the following way: A possibly adaptive interim analysis based on 215 included patients (at information fraction 0.5) serves to assess the risk-benefit ratio and may lead to (1) stopping (albeit non-mandatory) due to overwhelming non-inferiority, (2) stopping for futility/safety, (3) continuation as planned, or (4) recalculation of the sample size based on conditional power. No other adaptations will be allowed. Any adaptation of the study design suggested by the masked DMC will be based on the observed proportions of SAB-related late complications applying the inverse normal method (49).

#### 6.2. Sample size calculation

The Scientific Advisory Committee suggested a non-inferiority margin of 10 percentage points. However, since this may be felt too large by some clinicians for the clinical question to be addressed, the study sample size was initially calculated to ensure sufficient power (i.e. 80%) even with a reduced margin of 5 percentage points (as a compromise of precision and feasibility). Both margins, i.e. 10% and 5%, are tested hierarchically in order to ensure both sufficient power and type I error control. Due to difficulties in reaching the full sample, the previously planned interim analysis at 215 patients was converted into the final analysis. This conversion accommodates a non-inferiority margin of 10% while keeping all other parameters constant (s. below).

A large prospective cohort study of 324 SAB patients indicates that patients with a removable focus of infection have a low risk (2.4%) for late complications (26). In a similar prospective study of 211 SAB patients with a removable focus of infection, the relapse rate with antimicrobial therapy for less than 14 days (n=134) was 3.7% (27). A meta-analysis of studies performed between 1967 and 1993 reported a combined rate of late complications of 6.1% in patients with catheter-related infections (15).

Since the in-/exclusion criteria target patients at low risk for complications, we expect a true incidence of 2.5% of late complications in study patients. This is in line with our own (INSTINCT 3%, preSABATO 1%) and other studies (2.4% (19), and 3.7% (27)).

The sample size for a 10% non-inferiority margin without employing an interim analysis is calculated as follows: assuming 2.5% complications per arm, a non-inferiority margin of 10%, a one-sided alpha of 2.5% and a power of 80% requires 144 subjects in total (calculated with R version 3.3.3, package gsDesign, function nBinomial; R Foundation for Statistical Computing, Vienna, Austria), i.e. 144/0.9/0.9/0.95 = 187 subjects adjusted for deaths unrelated to SAB (10%), for protocol violations (10%) and for stratification (5%). Thus a sample size of 215 patients does accommodate a 10% non-inferiority margin.

Observing 2.5% late complications in the final analysis (with 215*0.9*0.9*0.95/2≈83 patients per group), i.e. 0.025*83≈2 late complications in each group, yields a 90% confidence interval for the difference of -0.049 to +0.049 (Stata 15.1, StataCorp LLC, College Station, TX, USA; rdcii).

The original sample size, which included an interim analysis and a 5% non-inferiority margin, was calculated as follows: The sample size for each study arm is 165.8 (non-inferiority margin 5%, one-sided $\alpha=0.05$, $\beta=0.2$, one interim analysis at information fraction 0.5 using the O'Brien-Fleming bound 2.373; calculated using ADDPLAN 6.0.1, ADDPLAN GmbH, Cologne). An allowance of 10% for deaths unrelated to SAB, of 10% for protocol violations and of 5% for stratification yields 331.6/0.9/0.9/0.95≈430 patients in total to be randomized. Note, if non-attributable mortality (i.e. about 10% within 90 days) were added to the composite endpoint, the sample size needed would approximately double (i.e. from 430 to 823 patients). Unfortunately, this cannot be achieved by the clinical network.

At the initially planned interim analysis observing 2.5% late complications, i.e. 0.025*166/2≈2 in each group, yields a 98.235% confidence interval (corresponds to the O'Brien-Fleming bound 2.373) for the difference of -8.2% to +8.2% (Newcombe's method 10; R. G. Newcombe, Interval estimation for the difference between independent proportions: comparison of eleven methods. Statistics in Medicine 17:873-90, 1998). At the initially planned final analysis observing 2.5% late complications, i.e. 0.025*166≈4 in each group, yields a 91.765% confidence interval (corresponds to the O'Brien-Fleming bound 1.678) for the difference of -3.4% to +3.4%.

### 7. Safety

#### 7.1. Definitions of adverse events and adverse drug reactions

##### 7.1.1. Adverse event

An adverse event (AE) is any clinically relevant untoward medical occurrence in a trial subject that received study drug and occurs in the time from first application of study drug until EOS. An AE includes any unfavourable and unintended sign (e.g. abnormal laboratory parameter), symptom, or disease that is temporally associated with the study drug, whether or not a causal relationship to the study drug is suspected.

For reasons of drug safety, pregnancy of a trial subject is to be regarded as an AE, when pregnancy occurs in the time from the first application of study drug until EOS. In female trial subjects, stopping the study drug should be considered when pregnancy becomes known.

Efficacy endpoint events as defined in chapter 4.7 (SAB related complication) are not only reported as AEs but are additionally reported in the appropriate endpoint module. When these events become serious AEs (SAE) they will be additionally reported to the DMC without delay.

Exceptions:

- Elevation of inflammatory markers (e.g. C-reactive protein [CRP], procalcitonin [PCT]) up to two-fold above baseline, as determined in the screening phase, does not represent an AE since it is expected as part of the underlying disease.
- All AEs CTCAE grade <3 (Common Terminology Criteria of Adverse Events, see 7.2.2) are not to be regarded as AEs.
- Concomitant diseases: Detoriations caused by pre-existing disease will be recorded as AEs if occurring after the first application of study drug until EOS. A preexisting disease that led to a treatment measure planned before the start of the clinical trial (e.g. admission to hospital as an inpatient) is not considered an AE.

##### 7.1.2. Adverse drug reaction

An adverse drug reaction (ADR) is any noxious and unintended response to a study drug related to any dose with at least a reasonably possible causal relationship with the study drug.

##### 7.1.3. Serious adverse events and serious adverse reactions

A serious AE (SAE) or serious ADR (SADR) is any AE that

1. Results in death,
2. Is life-threatening at the time of the event
3. Requires hospitalization or prolongation of existing hospitalization
4. results in persistent or significant disability/incapacity
5. is a congenital anomaly or birth defect (1.-4.: § 3(8) GCP Regulations)
6. In the opinion of the investigator, fulfils any other criteria similar to 1.–4.

Hospitalization is defined as any stay in hospital on the part of a trial subject that includes at least one night (midnight to 06:00).

The exceptions for AEs, as defined in 7.1.1, also apply to SAEs.

Admission to hospital as an inpatient scheduled prior to enrolment are not SAEs, but must be documented in the proper manner in the trial subject's medical records and eCRF (see Section 7.1.1).

If an AE is classified as an SAE, this is documented on a separate SAE report form in addition to the standard AE documentation.

SAEs have to be reported according to national and international regulations (for procedure, see 7.3)

##### 7.1.4. Unexpected adverse drug reaction

An unexpected ADR is an ADR that is not consistent in nature or severity with ADR listed in the appropriate reference document (Appendix 11.8).

##### 7.1.5. Suspected unexpected serious adverse reactions

A suspected unexpected serious adverse reaction (SUSAR) is an adverse event where the nature or severity of the AE is not consistent with the appropriate reference document (Appendix 11.8), is regarded as serious, and has at least a possible causal relationship with the study drug.

##### 7.1.6. Other possible trial-specific complications or risks

This study compares oral and standard intravenous treatment regimen in patients with low-risk SAB. The primary potential risk of the study is that the oral application of antimicrobials results in lower drug levels. This may result in a shorter duration of effective antimicrobial therapy. Possible consequences include failure to cure the S. aureus infection, progression of infection to other sites or relapse of the infection.

To detect and promptly treat complications of S. aureus infection, patients are asked to consult the local study site immediately or seek emergency treatment in case of fever, fatigue, pain, or any other complaints. In every study center, patients will have access to a 24/7 emergency department staffed with appropriately trained doctors. Patients will be provided with a card that contains information on the trial, trial-specific risks, and emergency contact numbers.

All antimicrobials in this trial are commercially available, have been in use for many years and are generally regarded as safe. However, there is still a risk of adverse events (AE) related to antimicrobial therapy. A description of AEs associated with each study drug is provided in the reference documents (Appendix 11.8). The appropriate reference document and additional information to the antimicrobial agents in the study protocol (e.g chapter 4.6) should be consulted prior to prescription and dose changes of any of the protocol-approved, commercially-available antimicrobials.

A specific risk for all antimicrobial agents is Clostridium difficile associated diarrhea (CDAD). Nearly all antimicrobial agents have been described to cause CDAD. Clinical symptoms may range from mild diarrhea to fatal colitis. Antibacterial agents alter the intestinal flora and lead to overgrowth of C. difficile. If CDAD is confirmed or suspected, ongoing antimicrobial use not directed against C. difficile may need to be discontinued. Fluid and electrolyte management, protein supplementation, antimicrobial treatment of CDAD and surgical evaluation should be instituted as clinically indicated.

##### 7.2. Documentation and follow-up of adverse events

The sponsor ensures that every person involved in the treatment of trial subjects is adequately informed about the responsibilities and actions required when an AE occurs. Patients will be asked at each visit or telephone interview during follow-up whether they have experienced AEs or SAEs.

###### 7.2.1. Documentation of adverse events and adverse drug reactions

For all AEs, unless exempted in section 7.1.1, that occur in the time from first application of study drug until EOS, the following information is documented in the eCRF and in the medical record:

- Date and time of onset and resolution
- Severity, according to CTCAE Version 4.0 (s. section 7.2.2)
- Causal relationship with study drug (s. section 7.2.3)
- Seriousness (s. section 7.1.3)
- Interruption or withdrawal of study treatment and other measures taken

Regardless of whether a causal relationship between the AE and the study drug is suspected, trial subjects who develop adverse events must be monitored until all symptoms have been subsided, pathological laboratory values have returned to pre-event levels, a plausible explanation is found for the AE, the trial subject has died, or the study has been terminated for the trial subject concerned.

###### 7.2.2. Severity of the adverse event

The authorized member of the study group will classify the severity of AEs according to the CTCAE Version 4.0. The CTCAE displays grades 1 through 5 with unique clinical descriptions of severity for each AE based on this general guideline:

- Grade 1 = mild: asymptomatic or mild symptoms; clinical or diagnostic observations only; intervention not indicated.
- Grade 2 = moderate: minimal, local or noninvasive intervention indicated; limiting age-appropriate instrumental activities of daily living (ADL)
- Grade 3 = severe: medically significant but not immediately life-threatening; hospitalization or prolongation of hospitalization indicated; disabling; limiting self care ADL
- Grade 4 = life-threatening consequences; urgent intervention indicated
- Grade 5 = death related to AE

###### 7.2.3. Causal relationship between adverse event and study drugs

The authorized member of the study group will assess for every AE whether a causal relationship with the study drug can be assumed or not. The assessment includes consideration of the nature and type of reaction, the temporal relationship with the study drug, the clinical status of the trial subject, concomitant medication and other relevant clinical factors. If the event is considered to be caused by lack of efficacy of the study drug or as a symptom or sign of the bloodstream infection, no causal relationship will be assumed.

The following definitions are used to assess the causal relationship between all AEs and the study drug (WHO Causality Assessment of Suspected Adverse Reactions):

- Certain: A clinical event, including laboratory test abnormality, occurring in a plausible time relationship to drug administration, and which cannot be explained by concurrent disease or other drugs or chemicals. The response to withdrawal of the drug (dechallenge) should be clinically plausible. The event must be definitive pharmacologically or phenomenologically, using a satisfactory rechallenge procedure if necessary.
- Probable/likely: A clinical event, including laboratory test abnormality, with a reasonable time sequence to administration of the drug, unlikely to be attributed to concurrent disease or other drugs or chemicals, and which follows a clinically

reasonable response on withdrawal (dechallenge). Rechallenge information is not required to fulfill this definition.

- Possible: A clinical event, including laboratory test abnormality, with a reasonable time sequence to administration of the drug, but which could also be explained by concurrent disease or other drugs or chemicals. Information on drug withdrawal may be lacking or unclear.
- Unlikely: A clinical event, including laboratory test abnormality, with a temporal relationship to drug administration which makes a causal relationship improbable, and in which other drugs, chemicals or underlying disease provide plausible explanations.
- Conditional/unclassified: A clinical event, including laboratory test abnormality, reported as an adverse reaction, about which more data is essential for a proper assessment or the additional data are under examination.
- Unassessable/unclassifiable: A report suggesting an adverse reaction which cannot be judged because information is insufficient or contradictory, and which cannot be supplemented or verified.

An ADR is suspected if the causal relationship is at least ‘possible’ or ‘conditional/unclassified’ or ‘unassessable/unclassifiable’. Events assessed as ‘unlikely’ are not suspected ADRs.

##### 7.3. Reporting and follow up of serious adverse events, pregnancy and changes in risk-benefit assessment

Regardless of the assumed causal relationship, every SAE and every pregnancy that qualifies as AE (s. section 7.1.1) must be documented in the appropriate part of the eCRF and additionally on the corresponding report form. The report form must be sent to the sponsor via Fax within 24h of becoming aware of the event (s. section 7.3.1. and 7.3.2). Details of SAE management are defined in a trial-specific SAE manual, which describes information and communication pathways, interfaces and clarification of responsibilities, and local statutory requirements of each participating country.

##### 7.3.1. SAE reports from the authorized member of the study group to the sponsor

The authorized member of the study group will inform the ZKS Cologne about the occurrence of an SAE without delay (at latest 24 hours after being made aware of the SAE) by sending a SAE report form , via fax to

ZKS Cologne, Gleueler Str. 269, 50935 Cologne

Each SAE must be followed up until:

- the SAE is no longer serious
- the patient dies
- the study ends for the individual patient.

The recurrence, deterioration or ending of an existing SAE will be documented as follow-up of the original SAE using a new SAE report form and will be reported to the sponsor immediately.

All SAEs are assessed by the sponsor with regard to seriousness (see Section 7.1.3.; 7.2.2.), causality (see Section 7.2.3) and expectedness (see Section 7.1.4), regardless of the assessment of the authorized member of the study group. If an AE is “serious”, at least “possibly related” and “unexpected”, the criteria for an expedited report (suspected unexpected serious adverse reaction (SUSAR)) are fulfilled.

Details concerning the SAE reports are described in the trial-specific SAE manual.

##### 7.3.2. Pregnancy reports from the study group to the sponsor

The authorized member of the study group will inform the ZKS Cologne without delay (at latest after 24h of being made aware) about any pregnancy that qualifies as AE (s. section 7.1.1). This will be documented on “Pregnancy report form I”. The pregnant trial subject will be asked to give separate informed consent for pregnancy follow up. A separate “Pregnancy report form II” has to be sent to ZKS Cologne immediately after being made aware of delivery (at latest 24h). Both parents are asked to give separate informed consent for child follow-up. The health of the child is monitored for 8 weeks post delivery.

Details concerning the reports are described in the trial-specific SAE manual.

If a SAE/SAR/SUSAR of mother or child occurs in the course of pregnancy or delivery, it has to be reported according to statutory requirements.

##### 7.3.3. Unblinding when treatment is blinded

Not applicable.

##### 7.3.4. Notification of ethics committee and competent authorities

Every SUSAR that becomes known in a clinical trial will be reported by the sponsor to the competent authorities of all countries where the trial is being conducted (unless waived), to the responsible ethic committee or other bodies according to local statutory requirements of each participating country, to the principle investigator of each participating site, and to the principle investigator of all clinical trials investigating the same active substance (study drug) by the sponsor. Every SUSAR must be sent without delay, respecting the timelines of each country (as detailed in the SAE manual), after the minimum reporting criteria are compiled.

Minimum reporting criteria include:

- Valid EudraCT number
- Sponsor study number
- Identifiable coded subject
- Identifiable reporter
- One SUSAR
- One suspect study drug (including active substance name)
- Causality assessment

##### 7.3.5. Review and reporting of changes in the risk-benefit ratio

The sponsor will inform without delay (respecting the timelines of each country) the competent authorities of all countries where the trial is conducted and the appropriate ethics committees about any event or factor that may change the risk-benefit ratio of the study drug. A change in the risk-benefit ratio may occur in the following cases:

- Individual reports of expected serious ADRs with an unexpected outcome
- A clinically relevant increase in the rate of occurrence of expected SADRs
- SUSARs in trial subjects who have already completed the follow-up period of the clinical trial (”end-of-trial visit”)
- Factors emerging in connection with trial conduct or the development of the study drug that may affect the safety of persons concerned.

##### 7.3.6. Informing the Data Monitoring Committee

Efficacy endpoint events (SAB related complication as defined in chapter 4.7) that have become serious (SAE) will be reported to the DMC without delay. Listings on these events will be presented to the DMC in appropriate intervals.

The sponsor will inform the DMC yearly of all safety-relevant events by sending a copy of the Development Safety Update Report (DSUR). The DSUR includes a cumulative list of all serious adverse events and a line listing of every serious adverse reaction during the reporting period. Details are defined in the DMC manual.

##### 7.3.7. Informing the investigators

The sponsor is responsible to provide the following information within the timeline set by the respective competent authorities to the investigator of each participating study site, who is responsible to disseminate the information within his study group (details will be described in the SAE manual):

- all relevant information from all SUSARs within the trial
- SUSAR-reports from other clinical trials investigating the same active substance by the same sponsor
- new scientific information on study drugs that becomes available

##### 7.3.8. Informing the marketing authorisation holder

There are no contractual agreements with the marketing authorisation holders.

#### 7.4. Annual safety report of trial subjects

Once per year or on request, the sponsor will supply a report on the safety of trial subjects with all available relevant information concerning patient safety during the reference period to the competent authorities of all countries where the trial is being conducted. This report will also be supplied to the DMC and the responsible ethics committee.

The annual safety report will be compiled according to the corresponding ICH guideline E2F „Development Safety Update Report – DSUR“. The annual data lock point for the patient data to be included defined in the trial-specific SAE manual. The sponsor will supply the report within 60 days after the annual data-lock point. The last report should be submitted within 60 days after the last visit of the last patient in the respective country.

### 8. Use of trial findings and publication

#### 8.1. Reports

##### 8.1.1. Interim reports

Section 7.4 describes the requirements for annual reports on the safety of trial subjects.

Interim reports are not part of the trial.

##### 8.1.2. Final report

The competent authorities and ethics committees will be informed within 90 days that the trial has officially ended. Within one year of the completion of the trial, the competent authorities and the ethics committees will be supplied with a summary of the final report on the clinical trial containing the main results.

#### 8.2. Publication

It is planned to publish the trial results, in mutual agreement with the PCI, in a peer-reviewed scientific journal. Publication of the results of the trial as a whole is intended. Any publication will take account of the ‘Uniform requirements for manuscripts submitted to biomedical journals’ (28).

The trial has been registered in a public register (see also Section 5.3).

Any published data will observe data protection legislation covering the trial subject and investigators. Success rates or individual findings at individual trial sites are known only to the sponsor.

A raw anonymised data set will be made available to the scientific community upon legitimate request to the sponsor once the trial is completed. Furthermore, an anomyzed dataset will be archived indefinetely in a publicly owned and controlled database, like clinicaltrials.gov.

Publications or lectures on the findings of the present clinical trial either as a whole or at individual investigation sites must be approved by the sponsor in advance, and the sponsor reserves the right to review and comment on such documentation before publication.

By signing the contract to participate in this trial, the investigator declares that he or she agrees to submission of the results of this trial to national and international authorities for approval and surveillance purposes, and to the Federal Physicians Association, the Association of Statutory Health Fund Physicians and to statutory health fund organisations, if required. At the same time, the investigator agrees that his or her name, address, qualifications and details of his or her involvement in the clinical trial may be made known to these bodies.

#### 9. Amendments to the trial protocol

To ensure that comparable conditions are achieved as far as possible at individual trial sites and in the interests of a consistent and valid data analysis, changes to the provisions of this trial protocol are not planned. In exceptional cases, however, changes may be made to the trial protocol. Such changes can only be made if agreed by the sponsor, sponsor's representative, the PCI and biometrician, and all Authors of this trial protocol. Any changes to the trial procedures must be made in writing and must be documented with reasons and signed by all Authors of the original trial protocol.

Amendments made in accordance with § 10 Secs. 1 and 4 GCP Regulations that require approval are submitted to the ethics committee and the competent authorities and will not be implemented until approved. Exceptions to this are amendments made to avoid immediate dangers.

29. The European Agency for the Evaluation of Medicinal Product. Note for Guidance on Good Clinical Practice (CPMP/ICH/135/95).

30. The European Agency for the Evaluation of Medicinal Product. Note for Guidance Structure and Content of Clinical Study Reports (CPMP/ICH/137/95).

31. National Cancer Institute. Protocol Templates, Applications and Guidelines http://ctep.cancer.gov/guidelines/templates.html.

32. EMEA-Guideline On Data Monitoring Committees: EMEA/CHMP/EWP/5872/03 Corr

33. The DAMOCLES Study Group. A proposed charter for clinical trial 2005 data monitoring committees: helping them do their job well. Lancet 2005; 365: 711-22

34. Clinical trial registration: a statement from the International Committee of Medical Journal Editors. Accessed at http://www.icmje.org/clin_trial.pdf on 22 May 2007.

35. WHO. Causality Assessment of Suspected Adverse Reactions. http://www.who-umc.org/DynPage.aspx?id=22682

47. Committee for Medicinal Products for Human Use (CHMP), European Medicines Agency (15 December 2011) Guideline on the evaluation of medicinal products indicated for treatment of bacterial infections (CPMP/EWP/558/95 rev 2)

48. Committee for Medicinal Products for Human Use (CHMP), European Medicines Agency (27 July 2000) Points to consider on switching between superiority and non-inferiority (CPMP/EWP/482/99)

49. Lehmacher W, Wassmer G. Adaptive Sample Size Calculations in Group Sequential Trials. Biometrics. 1999; 55(4): 1286-90


This document is based on a document by G. Grass and C. Weiß which is subject to the UVM licence for unprotected content and may only be used if the license is adhered to (http://www.ifross.de/Lizenzen/LizenzFuerFreieInhalte.html).

### 11. Appendices

#### 11.1. Trial sites and principle investigators

| Nr | Site | PI | Institution |
|---|---|---|---|
| 1 | Cologne | Prof. G. Fätkenheuer | Department I of Internal Medicine, University of Cologne |
| 2 | Freiburg | Prof. W.V. Kern | Department of Medicine, University Hospital Freiburg |
| 3 | Berlin (closed) | Prof. K. Arastéh | Vivantes Auguste-Viktoria-Klinikum, and Vivantes Wenckebach-Klinikum, Berlin |
| 4 | Krefeld | Dr. K. Kösters | Helios Klinikum Krefeld |
| 5 | Hannover | Prof. T. Welte | Department of Respiratory Medicine, Hannover Medical School, Hannover |
| 6 | Jena | Prof. M. Pletz | Department of Anesthesiology and Intensive Care Medicine, Jena University Hospital, Jena |
| 7 | Aachen (closed) | Prof. S. Lemmen | Central Unit of Hospital Infection and Hygiene, Universitätsklinikum Aachen |
| 8 | Lübeck | Prof. J. Rupp | Medical Clinic III, University of Schleswig-Holstein, Lübeck |
| 9 | Leverkusen (closed) | Prof. Dr. S. Reuter | Medical Clinic 4, Klinikum Leverkusen |
| 10 | Regensburg (closed) | Prof. B. Salzberger | Medical Clinic I, University Hospital Regensburg |
| 11 | Frankfurt (closed) | Prof Dr. C. Stephan | Department of Infectious Diseases, J.W. Goethe University Hospital Frankfurt |
| 12 | Ulm (dropped) | | Section of Infectius Diseases, Department of Medicine III, University Hospital Ulm |
| 13 | Groningen (closed) | Dr. M. Wouthuyzen-Bakker | Department of Infectious Diseases, University Medical Center Groningen, NL |
| 14 | Breda (closed) | Prof. J. Kluytmans | Amphia Hospital, Breda, NL |
| 15 | Tilburg Nl (dropped) | Prof. J. Kluytmans | St. Elisabeth Hospital, Tilburg |
| 16 | Amsterdam (dropped) | Prof. J.T.M. van der Meer | Department of Internal Medicine and Infectious Diseases, Academic Medical Center, Amsterdam, NL |
| 41 | VUmc Amsterdam (closed) | Dr. K. van Dijk | Arts-microbioloog, VUmc, Afd. Medische Microbiologie en Infectiepreventie, Amsterdam, NL |
| 17 | UMC Utrecht (closed) | Prof. M. Bonten | University Medical Center Utrecht, NL |

| | | | |
|---|---|---|---|
| 18 | Barcelona 1 | Prof. A. Soriano | Department of Infectious Diseases, Hospital Clínic of Barcelona, ES |
| 40 | Utrecht (closed) | Dr. M. Vlek | Diakonessenhuis,Utrecht, NL |
| 19 | Barcelona 2 (dropped) | Prof. B. Almirante | Hospital Universitari Vall d'Hebron, Barcelona, ES |
| 20 | Sevilla VM | Prof. J. Rodríguez-Baño | Infectious Diseases Section, Hospital Universitario Virgen Macarena, Sevilla, ES |
| 21 | Sevilla VR | Prof. J. Cisneros | Infectious diseases, Microbiology and Preventive Medicine Department, Hospital Universitario Virgen del Rocío, Sevilla, ES |
| 22 | Nottingham (dropped) | Prof. D. Turner | Nottingham University Hospitals NHS Trust, UK |
| 23 | Palma | Prof. M. Riera | Seccio de Malalties Infeccioses, Hospital Universitario Son Espases, Palma de Mallorca, ES |
| 24 | Düsseldorf | Prof. A. Kaasch | Institute of Medical Microbiology and Hospital Hygiene, University Hospital Düsseldorf |
| 25 | Annecy | Dr. J. Gaillat | Centre Hospitalier Annecy Genevois, F |
| 26 | Paris 1 | Prof. E. Rouveix-Nordon | APHP Ambroise Paré, Paris, F |
| 27 | Paris 2 (closed) | Dr. F. Mechai | APHP Avicenne, Paris, F |
| 28 | Paris 3 | Prof. B. Fantin | APHP Beaujon, Paris, F |
| 29 | Paris 4 | Dr. R. Lepeule | APHP Mondor, Paris, F |
| 30 | Paris 5 | Prof. J. Molina | APHP St. Louis, Paris, F |
| 31 | Chambéry | Dr. E. Forestier | CH Métropole Savoie, Chambéry, F |
| 32 | La Roche sur Yon | Dr. T. Guimard | CH Départemental de Vendée, La Roche sur Yon, F |
| 33 | Nantes | Prof. D. Boutoille | CHU Nantes, F |
| 34 | Orléans | Dr. L. Hocqeloux | CH Orléans, F |
| 35 | Quimper | Prof. J. Tallarmin | CH De Cournouaille, Quimper, F |
| 36 | Rennes | Prof. P. Tattevin | CHU Rennes, F |
| 37 | St. Etienne | Prof. F. Lucht | CHU St. Etienne, F |
| 38 | Grenoble | Prof. J.-P. Stahl | CHU Grenoble Alpes, F |
| 39 | Tours | Prof. L. Bernard | CHRU Tours, F |

#### 11.2. Protocol Agreement Form

#### 11.3. Steering Committee

Prof. Achim Kaasch, Düsseldorf, Germany

Prof. Harald Seifert, Cologne, Germany

Prof. Winfried Kern, Freiburg, Germany

PD Dr. Siegbert Rieg, Freiburg, Germany

Prof. Gerd Fätkenheuer, Cologne, Germany

Prof. Dr. Martin Hellmich, Cologne, Germany

Prof. Frank Brunkhorst, Jena, Germany

International members:

Prof. Alex Soriano, Barcelona, Spain

Prof. Marc Bonten, Utrecht, The Netherlands

#### 11.4. Data Monitoring Committee

Prof. Werner Haefeli, Department of Clinical Pharmacology and Pharmacoepidemiology, University Hospital Heidelberg, Germany

Prof. Alexandra Heininger, Department für Infektiologie, Sektion Krankenhaushygiene, Heidelberg University, Germany

Dr. Geraldine Rauch, Institute of Medical Biometry and Informatics, Heidelberg University, Germany

#### 11.5. Scientific Advisory Committee

Prof. Vance Fowler, Center for Microbial Pathogenesis, Duke University, Durham, USA

Prof. Guy Thwaites, Oxford University Clinical Research Unit, Ho Chi Minh City, Vietnam

Prof. Stephan Harbarth, Maladies Infectieuses, Hôpitaux Universitaires de Genève, CH

Prof. Winfried Kern, Freiburg, Germany

Prof. Harald Seifert, Cologne, Germany

#### 11.6. Clinical Review Committee

Prof. Oliver Cornely, University of Cologne, Germany

Estee Török, Cambridge Univerisity, UK

One further member to be named.

#### 11.7. Central study laboratory

Institute of Medical Microbiology and Hospital Hygiene, Universitätsstr. 1, Heinrich-Heine-University, Düsseldorf, Germany

#### 11.8. Reference documents for study drugs

One reference document is chosen for the active substance.

Trimethoprim/sulfamethoxazole: http://www.uptodate.com/contents/trimethoprim-sulfamethoxazole-co-trimoxazole-drug-information (as accessed 2018-03-20)

Clindamycin:http://www.uptodate.com/contents/clindamycin-systemic-drug-information (as accessed 2018-03-20)

Linezolid: http://www.uptodate.com/contents/linezolid-drug-information (as accessed 2018-03-20)

Flucloxacillin: 1g powder by Wockhardt UK Ltd, http://www.medicines.org.uk/emc/product/2239/smpc (as updated 2018-01-12, accessed 2018-03-20)

Cloxacillin: http://www.uptodate.com/contents/cloxacillin-drug-information (as accessed 2018-03-20)

Cefazolin: http://www.uptodate.com/contents/cefazolin-drug-information (as accessed 2018-03-20)

Vancomycin: http://www.uptodate.com/contents/vancomycin-drug-information (as accessed 2018-03-20)

Daptomycin: http://www.uptodate.com/contents/daptomycin-drug-information (as accessed 2018-03-20)

#### 11.9. Patient information sheet and informed consent form

#### 11.10. Prescribing information

Not applicable

#### 11.11. Confirmation of insurance

#### 11.12. Conditions of insurance
