## Supplementary material for "Early oral switch in low-risk *Staphylococcus aureus* bloodstream infection": Statistical Analysis Plan

UNIKLINIK KÖLN
Institut für Medizinische Statistik und Bioinformatik

imsb

**AG Medizinische Statistik**
**Prof. Dr. Martin Hellmich**

### Statistical Analysis Plan

| | |
|---|---|
| Study title: | Early oral switch therapy in low-risk staphylococcus aureus bloodstream infection |
| Study code: | SABATO |
| Study design: | Multicenter, multinational clinical trial, two arms, randomized, open-label, non-inferiority, parallel-group |
| Indication: | *Staphylococcus aureus* bloodstream infection |
| Investigational intervention: | Orally administered antimicrobial – Oral switch therapy (OST) |
| Comparator: | Intravenously administered antimicrobial – Intravenous standard therapy (IST) |
| Sponsor (or representative): | Heinrich-Heine-Universität Düsseldorf<br>Universitätsstr. 1<br>40225 Düsseldorf<br>Germany<br>represented by the principal investigator |
| Financial support: | Deutsche Forschungsgemeinschaft (DFG, German Research Foundation; grant number KA 3104/2-1 and KA 3104/2-2) |
| Protocol identification: | Uni-Koeln-1400 |
| Principal Coordinating investigator: | Prof. Dr. Achim Kaasch<br>Institute of Medical Microbiology and Hospital Hygiene<br>Düsseldorf University Hospital<br>Universitätsstr. 1<br>40225 Düsseldorf<br>Germany |
| Statistics: | Institute of Medical Statistics and Computational Biology (IMSB)<br>Kerpener Str. 62<br>50937 Cologne<br>Germany |
| SAP author: | Anne Adams (IMSB) |

CONFIDENTIAL

SAP version: V02 final Date: 15.04.2021

**Approved by**

| | | |
|---|---|---|
| Prof. Dr. Achim Kaasch<br>Principal Investigator | 21.04.2021<br>Date | A. Kaasch<br>Signature |
| Prof. Dr. Martin Hellmich<br>Statistician | 22.04.2021<br>Date | M. Hellmich<br>Signature |
| Anne Adams, MSc<br>Statistician | 15.04.2021<br>Date | Anne Adams<br>Signature |

### 1 Background

#### 1.1 Trial objective

The primary objective of this trial is to demonstrate that in patients with low-risk *S. aureus* bloodstream infection (SAB) a switch from intravenous to oral antimicrobial therapy (oral switch therapy, OST) is non-inferior to a conventional course of intravenous therapy (intravenous standard therapy, IST). This will be achieved by comparing the rate of SAB-related complications (relapsing SAB, deep-seated infection with *S. aureus*, or mortality attributable to SAB) within 90 days. Low-risk SAB manifests itself typically in patients with comorbidities. Therefore, survival is largely determined by the underlying disease and was not chosen as a primary endpoint. However, death related to SAB is included in the primary endpoint. Death unrelated to SAB will be carefully evaluated and compared.
The secondary objective is to measure the potential benefit for the patient. This is achieved by evaluating the length of hospital stay after the first positive blood culture and complications of intravenous therapy. A considerable number of patients on OST are expected to be discharged earlier from hospital, since hospital stay due to intravenous therapy is no longer required. This will reduce the risks associated with hospitalization and i.v. therapy (catheter-related infection, venous thrombosis, and septic thrombophlebitis) and is likely to improve patients' quality of life.

#### 1.2 Trial design

The study is a Phase III, multicentre, open-label, randomized, controlled, non-inferiority trial to test whether a switch form intravenous to oral antimicrobial therapy (oral switch therapy, OST) is non-inferior to a conventional course of intravenous therapy (intravenous standard therapy, IST) in patients with SAB. The clinical trial will be carried out as a multicentre open trial at trial sites in Germany, the Netherlands, France, and Spain.
Patients are randomly allocated to treatment arms (1:1) not earlier than one day before starting study drug. This is achieved by a central 24-7 Internet randomization service ALEA (stratified by study center, permuted blocks of varying length). Authorized local study staff may login to a secure website, randomize a patient and receive an email with attached pdf giving all the details on the allocated treatment. The randomization service is set up and maintained by IMSB, University of Cologne.

**Figure 1: Trial flowchart**

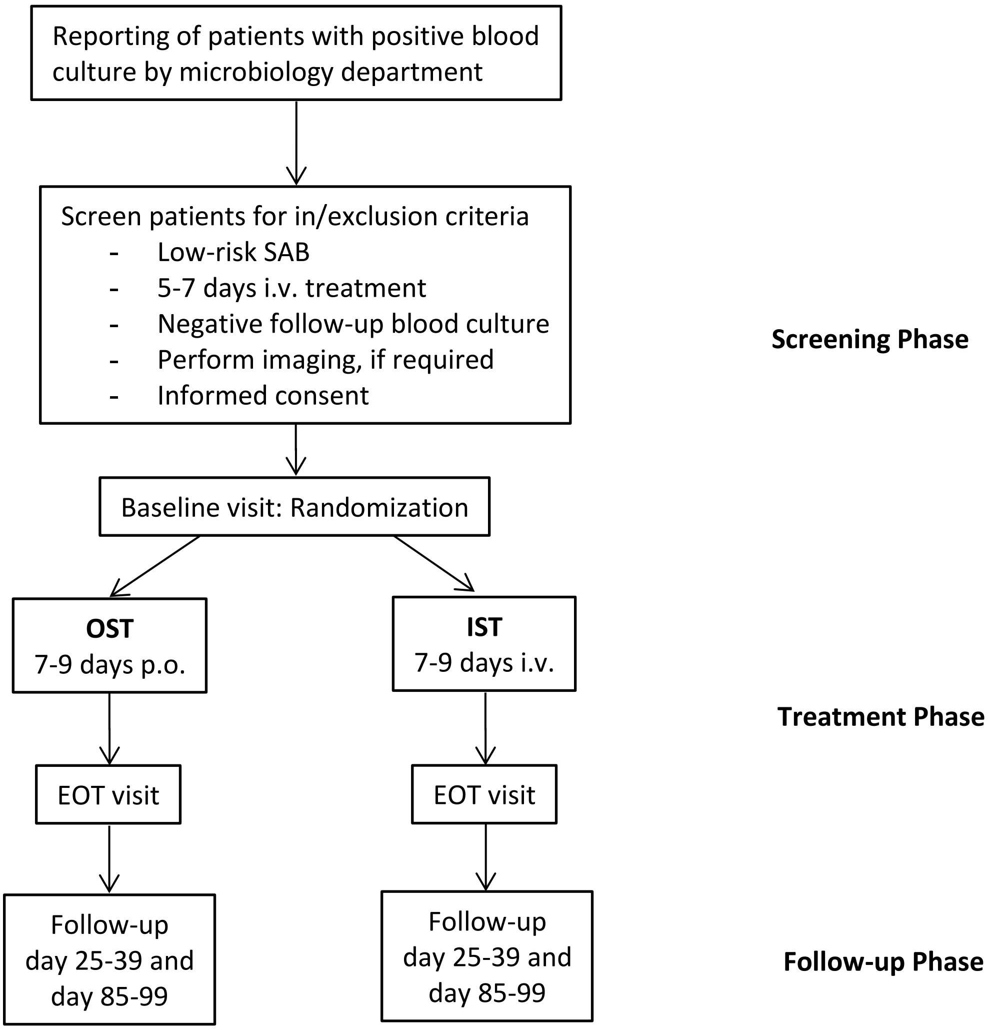

This study is an open-label study that uses standard doses of medication. Therefore, the study drugs are not centrally distributed, repackaged, or labelled. All antimicrobial treatments are commercially available antimicrobials approved by the respective national authorities.

Depending on the susceptibility of the isolates, expected drug interactions, contraindications and expected side effects, the drug is chosen from the corresponding list below. The drugs of first choice must be administered unless allergy or intolerance is suspected or resistance of the *S. aureus* isolate has been demonstrated.

**OST (oral administration)**

| MSSA | MRSA |
|---|---|
| 1st choice: Trimethoprim/sulfamethoxazole p.o. | 1st choice: Trimethoprim/sulfamethoxazole p.o. |
| 2nd choice: Clindamycin p.o. | 2nd choice: Linezolid p.o. |

**IST (intravenous administration)**

| MSSA | MRSA |
|---|---|
| 1st choice: Flucloxacillin i.v. | 1st choice: Vancomycin i.v. |
| (Cloxacillin in Spain & France) or Cefazolin i.v. | |
| 2nd choice: Vancomycin i.v. | 2nd choice: Daptomycin i.v. |

This study compares intravenous and oral treatment regimens. Neither patients nor investigators can be fully masked regarding treatment. However, clinical response to treatment will be evaluated in a blind manner by the Clinical Review Committee (CRC), though unblinding may be requested (or may be unavoidable, e.g. when judging appropriateness of antibiotic therapy) for individual cases. Moreover, the Data Monitoring Committee (DMC) will be masked, though unblinding may be requested for individual cases.

#### 1.3 Timing of analyses

After completion of the last visit of the last subject, when (at best) 215 subjects passed the complete trial, data will be cleaned and approved by CTCC according to their SOPs. The cleaned data will then be transferred to the statistician and the final analysis will be performed after the finalization and approval of this statistical analysis plan (SAP).

### 2 Analysis populations

#### 2.1 Definitions

All analyses will be done on three study populations:

The primary analysis set is derived from the per-protocol (PP) population. This dataset includes all study subjects who were essentially treated according to protocol and reached a defined endpoint in the trial (SAB-unrelated deaths will be excluded). The evaluability of study subjects will be assessed in a blind manner by the CRC. Details of the selection criteria and evaluation criteria are laid down in the CRC manual.

The secondary analysis set is derived from the intention-to-treat (ITT) population. This dataset includes all randomized study subjects, analyzed as assigned, with indeterminate and missing outcomes counted as failures. For time-to-event outcomes these cases are censored. Following current recommendations, CPMP/EWP/558/95 rev 2 and CPMP/EWP/482/99, the primary analysis is based on the per-protocol set; the analysis of the full analysis set (intention-to-treat, all randomized patients) will be of equal importance and should lead to similar conclusions for a robust interpretation.

The ITT population will be analysed as (ITT-1) any patient randomized, (ITT-2) only patients that were randomized AND received study drug without patients in whom a major inclusion criterion was violated.

The tertiary analysis set is for safety purposes (safety population). This dataset includes all study subjects who received any study drug as treated. Specifically, patients who ever received an oral antibiotic are compared to patients who never received an oral antibiotic.

### 4 Analysis variables

#### 4.1 Demography and baseline characteristics

Demography and baseline characteristics will include age, sex, weight, transfer from another hospital, prior hospitalization during one year before randomization, length of hospital stay before randomization, surgery within 30 days before randomization, immunosuppressive therapy and underlying condition (e.g. Charlson score).

- Two initial bloodcultures positive for *S. aureus* exhibit a positive differential time to positivity and there is no other plausible source of infection.

**Deep-seated infection** is any deep-seated focus of *S. aureus* infection resulting from hematogenous dissemination. In case of a microbiologically documented deep-seated infection, diagnosis requires either a positive culture from the respective site, or a blood culture positive with *S. aureus* plus imaging studies showing the presumed focus. In case of a clinically suspected deep-seated infection, microbiological results are not available or there is a plausible reason for a negative result, as finally judged by the CRC on all available evidence.
Deep-seated foci consist of, but are not limited to:

- Infective endocarditis, judged by modified Duke criteria
- Vertebral and non-vertebral osteomyelitis
- Suppurative arthritis
- Spinal empyema
- Muscle abscess (e.g. psoas abscess)
- Meningitis, brain abscess
- Lung abscess
- Visceral abscess (kidney, liver, spleen, etc.)

Catheter-related infections, superficial skin-and soft tissue infections such as thrombophlebitis, or superficial wound infections do not qualify as “deep-seated”, since they are likely to result from a new infection.

There is a possibility that late complications of SAB may be overlooked. By educating the patient about signs and symptoms of potential late complications, we expect that a diagnostic work-up will be performed in nearly all suspected cases. In the case of a suspected complication, the patient’s current care-providers will be contacted by the principle investigator for relevant clinical information and lab reports.

In the final report detailed patient data will be listed in a table for patients that reached the primary endpoint.

##### 4.3 Secondary variables

The **length of hospital stay** is defined as the number of days a patient spends in the hospital from date of the first positive blood culture to discharge. If the date of the first positive blood culture is prior to the date of admission to the hospital, the date of admission is used instead. When a patient is transferred to another hospital, days spent at the other hospital are included. Note: In-hospital days due to re-admissions are not counted.

**Survival** will be assessed during the hospital stay and at the follow-up telephone interviews. Death will be attributed to SAB when at least one of the following conditions is present:

- positive blood culture for *S. aureus* drawn within 72h before death
- persistent focus of deep-seated *S. aureus* infection at time of death

- persistent signs and symptoms of systemic infection at time of death as judged by study physician
- post-mortem analysis proving *S. aureus* related complication as cause of death

All other causes will be classified as unrelated to SAB.

The survival time is defined as the number of days a patient survives from the date of the first positive blood culture to (i) death, or (ii) end of the study, or (iii) loss of contact or withdrawal from the study.

**Complications of i.v. therapy** will be assessed from chart data, assessment of the study physician and from the patient interview. Complications may include, but are not limited to:

- local complications, such as: infiltration, extravasation, hematoma, phlebitis, thrombosis, thrombophlebitis, or infection at catheter insertion site
- systemic complications, such as: embolism, systemic infection, circulatory overload, allergic reaction
- in vancomycin therapy: “red man syndrome” due to inappropriately fast i.v. administration
- any other complication felt to be due to intravenous therapy by the principle investigator

In patients that report diarrhea during the intervention or follow-up phase, testing for ***Clostridium difficile*** **associated diarrhea (CDAD)** is not mandatory and will be performed according to the standard of care at the respective trial site. Furthermore, AEs and SAEs are collected until EOS.

### 5 Handling of missing values and outliers

#### 5.1 Missing values

Missing values will not be substituted. If subjects drop-out before end of study all data collected up until the time of subject withdrawal is to be entered into the electronic case report form (eCRF). In addition, every attempt should be made to collect follow-up information except for those subjects who specifically withdraw consent for release of such information.

For analysis of the primary endpoint the impact of missing data will be assessed in a sensitivity analysis by single/multiple imputation (based on logistic regression modelling).

For the analyses of the intention-to-treat population indeterminate and missing outcomes will be counted as failures.

#### 5.2 Outliers

No outlier analysis will be done.

### 6 Statistical analyses / methods

#### 6.1 Patient disposition

The number of patients that were screened will be counted from the screening data. The frequency of screening failures due to inclusion and exclusion criteria will be reported. Multiple selections per patient were possible.

Application of the inclusion and exclusion criteria to all included subjects will be verified.

Frequencies will be shown in a subject flow diagram. Number of and reasons for drop-outs will be included.

#### 6.2 Demography and baseline characteristics

The first measurements (first study visit) are the baseline values. Demography and baseline characteristics (including age, sex, weight, transfer from another hospital, prior hospitalization during one year before randomization, length of hospital stay before randomization, surgery within the 30 days before randomization, immunosuppressive therapy, underlying condition, Charlson score etc.) will be summarized descriptively by treatment arm and in total for PP, ITT, and safety population. Further variables are listed in the appendix.

Descriptive statistical analyses include: number of subjects (n), arithmetic mean, standard deviation (SD), median, first quartile (Q1), third quartile (Q3), minimum (min), maximum (max) for quantitative variables and absolute and relative frequencies for qualitative variables.

#### 6.3 Primary analysis

The primary endpoint SAB-related complications (relapsing SAB, deep-seated infection with *S. aureus*, or mortality attributable to SAB) within 90 days will be evaluated regarding non-inferiority of oral vs. intravenous antimicrobial therapy by Zhao's test (test 1) of non-null hypothesis on proportions stratified by study center at one-sided level 5% and with a non-inferiority margin of 10%. Thus, the hypotheses in terms of the proportion p of patients with SAB-related complications to be decided upon are

(A: null hypothesis) $H_0$: $p_{OST} > p_{IST} + 0.10$ vs

(A: alternative hypothesis) $H_a$: $p_{OST} \leq p_{IST} + 0.10$

If this null hypothesis can be rejected (fixed sequence of hypotheses, thus no alpha-inflation), the above test (A) will be repeated at one-sided level 2.5%.

If this null hypothesis can also be rejected (fixed sequence of hypotheses, thus no alpha-inflation), the non-inferiority margin of 5% will be applied, i.e.

(B: null hypothesis) $H_0$: $p_{OST} > p_{IST} + 0.05$ vs

(B: alternative hypothesis) $H_a$: $p_{OST} \leq p_{IST} + 0.05$

If this null hypothesis can also be rejected (fixed sequence of hypotheses, thus no alpha-inflation), the above test (B) will be repeated at one-sided level 2.5%.

Corresponding test-based confidence intervals will be calculated to aid interpretation.

The impact of missing data will be assessed in a sensitivity analysis by single/multiple imputation (based on logistic regression modelling).

#### 6.4 Secondary analyses

Secondary endpoints are evaluated by descriptive methods (by treatment group), including generalised linear modelling, methods for rates, proportions and the time to event.

Time to event endpoints such as length of hospital stay or 14, 30 and 90-day survival are analysed with the Kaplan-Meier method. The log-rank test is used to compare survival curves between the two treatment groups. In addition, multivariable Cox regression models can be fitted.

The analysis of length of hospital stay is as follows: (i) Patients who died in hospital or were discharged are counted as events (no censoring); (ii) patients who died in hospital are censored, patients discharged are counted as events; (iii) patients who died in hospital are counted as events, patients discharged are censored (“reverse” Kaplan-Meier method). In addition to Kaplan-Meier curves, descriptive measures such as median and interquartile are reported, too.

Complications of intravenous therapy will be counted by treatment group and evaluated with methods for rates or proportions.

#### 6.5 Safety

AE/SAEs will be MedDRA coded and listed / summarized by treatment group, system organ class, preferred term, severity and relationship. Further safety variables (esp. laboratory data) will be listed / summarised by treatment group using valid count and either percentage (qualitative data) or mean, standard deviation and (0, 25, 50, 75, 100) percentiles (quantitative data).

*Clostridium difficile* associated diarrhea (CDAD) will be counted by treatment group and analysed with methods for rates or proportions.

#### 6.6 Subgroup analyses

Subgroup analyses (of primary and secondary endpoints) are performed by sex (male-female ratio 3:2), country (not study center since there are too many (32)) and antibiotic susceptibility status (MSSA/MRSA), respectively. Further subgroup analysis will be conducted for (1) different foci of infection, (2) patients with comorbidities (pacemaker, prosthetic joints, moderate or severe liver disease, end-stage renal failure, immune suppression (at least one of the following: corticoid therapy, neutropenia, current antineoplastic therapy, immunosuppressive therapy, organ or marrow transplant)), (3) Charlson Comorbidity Index (<3 vs. >=3), (4) performance of diagnostic echocardiography (TEE/TTE), and (5) age groups (<65y vs. >= 65y). At least 10 patients in each subgroup will be necessary for analysis. The infective focus will be grouped in “central venous catheter” (central venous catheter, Shaldon catheter, Hickman/Broviac catheter, PICC line, and port system), “peripheral catheter” (peripheral venous catheter and arterial catheter), “skin and soft-tissue” (skin and soft tissue infection (without surgical wound) and surgical wound), “other”, and “focus not identified”.

##### 6.7 Additional Analysis

Detailed data from patients that died unrelated to SAB, and are thus excluded from PP analysis, will be listed.

##### 6.8 Interim analysis

An initially planned interim analysis will be converted into a final analysis, mainly due to difficulties in reaching the full sample size. Previously, an interim analysis was planned in the following way: A possibly adaptive interim analysis based on 215 included patients (at information fraction 0.5) serves to assess the risk-benefit ratio and may lead to (1) stopping (albeit non-mandatory) due to overwhelming non-inferiority, (2) stopping for futility/safety, (3) continuation as planned, or (4) recalculation of the sample size based on conditional power. No other adaptations will be allowed. Any adaptation of the study design suggested by the masked DMC will be based on the observed proportions of SAB-related late complications applying the inverse normal method.

#### 7 Data problems

Not expected. If any, circumstances and treatment will be documented in the final analysis report.

#### 8 Software

Data preparation and statistical analyses will be performed using SPSS Statistics (IBM Corp., Armonk, NY, USA) and SAS (SAS Institute Inc., Cary, NC, USA).

#### 9 References

Clinical study protocol Uni-Koeln-1400 from 20.03.2018 (Amendment III).

### 10 Appendices

#### 10.1 List of abbreviations

| | |
|---|---|
| AE | Adverse event |
| CDAD | *Clostridium difficile* associated diarrhea |
| CRC | Clinical review committee |
| CTCC | Clinical Trials Center Cologne |
| DFG | Deutsche Forschungsgemeinschaft |
| DMC | Data monitoring committee |
| eCRF | Electronic case report form |
| EOT | End of treatment |
| IMSB | Institute of Medical Statistics and Computational Biology |
| IST | Intravenous standard therapy |
| ITT | Intention to treat |
| i.v. | Intravenous |
| max | Maximum |
| min | Minimum |
| MRSA | Methicillin-resistant *Staphylococcus aureus* |
| MSSA | Methicillin-susceptible *Staphylococcus aureus* |
| p.o. | Per os (oral) |
| OST | Oral switch therapy |
| PP | Per protocol |
| Q1 | First quartile |
| Q3 | Third quartile |
| SAB | *Staphylococcus aureus* bloodstream infection |
| SAE | Serious adverse event |
| SAP | Statistical analysis plan |
| SD | Standard deviation |
| SOP | Standard operating procedure |

#### 10.2 List of variables for demography

| | |
|---|---|
| 1 | SAB related complication within 90 days |
| 2 | Mortality |
| 3 | Death attributable to SAB |
| 4 | In hospital days |
| 5 | Days on the intensive care unit |
| 6 | Complication of i.v. therapy |
| 7 | Clostridium difficile associated diarrhea |
| 8 | Diarrhea |
| 9 | 90-day mortality |
| 10 | Methicillin resistance |
| 11 | Daptomycin resistance |
| 12 | Linezolid resistance |
| 13 | Clindamycin resistance |
| 14 | Cotrimoxazol resistance |
| 15 | Remaining days of study drug therapy |
| 16 | Intravenous drug use |
| 17 | Systemic corticosteroid therapy for >1 week |
| 18 | Neutropenia |
| 19 | Anti-neoplastic chemotherapy |
| 20 | Any other immune suppressive therapy |
| 21 | Organ or marrow transplant |
| 22 | Vascular catheter in situ at onset |
| 23 | Focus of infection established |
| 24 | Pitt Bacteremia Score |
| 25 | CRP |
| 26 | Blood pressure (systole, diastole) |
| 27 | Body temperature |
| 28 | Heart rate |
| 29 | Respiratory rate |
| 30 | History of myocardial infarction (Charlson Score) |
| 31 | Congestive heart failure (Charlson Score) |
| 32 | Peripheral vascular disease (Charlson Score) |
| 33 | Cerebrovascular disease (Charlson Score) |
| 34 | Hemi-/Para-/Tetraplegia (Charlson Score) |
| 35 | Dementia (Charlson Score) |
| 36 | Liver disease (Charlson Score) |
| 37 | Renal disease (Charlson Score) |
| 38 | Chronic lung disease (Charlson Score) |
| 39 | Diabetes (Charlson Score) |
| 40 | Connective tissue disorder (Charlson Score) |
| 41 | Peptic ulcer (Charlson Score) |
| 42 | Malignancy (solid tumor) (Charlson Score) |
| 43 | Leukaemia (acute or chronic) (Charlson Score) |
| 44 | Lymphoma (Charlson Score) |
| 45 | HIV (Charlson Score) |
| 46 | Charlson Comorbidity Index |

**Figure 2: Analysis sets**

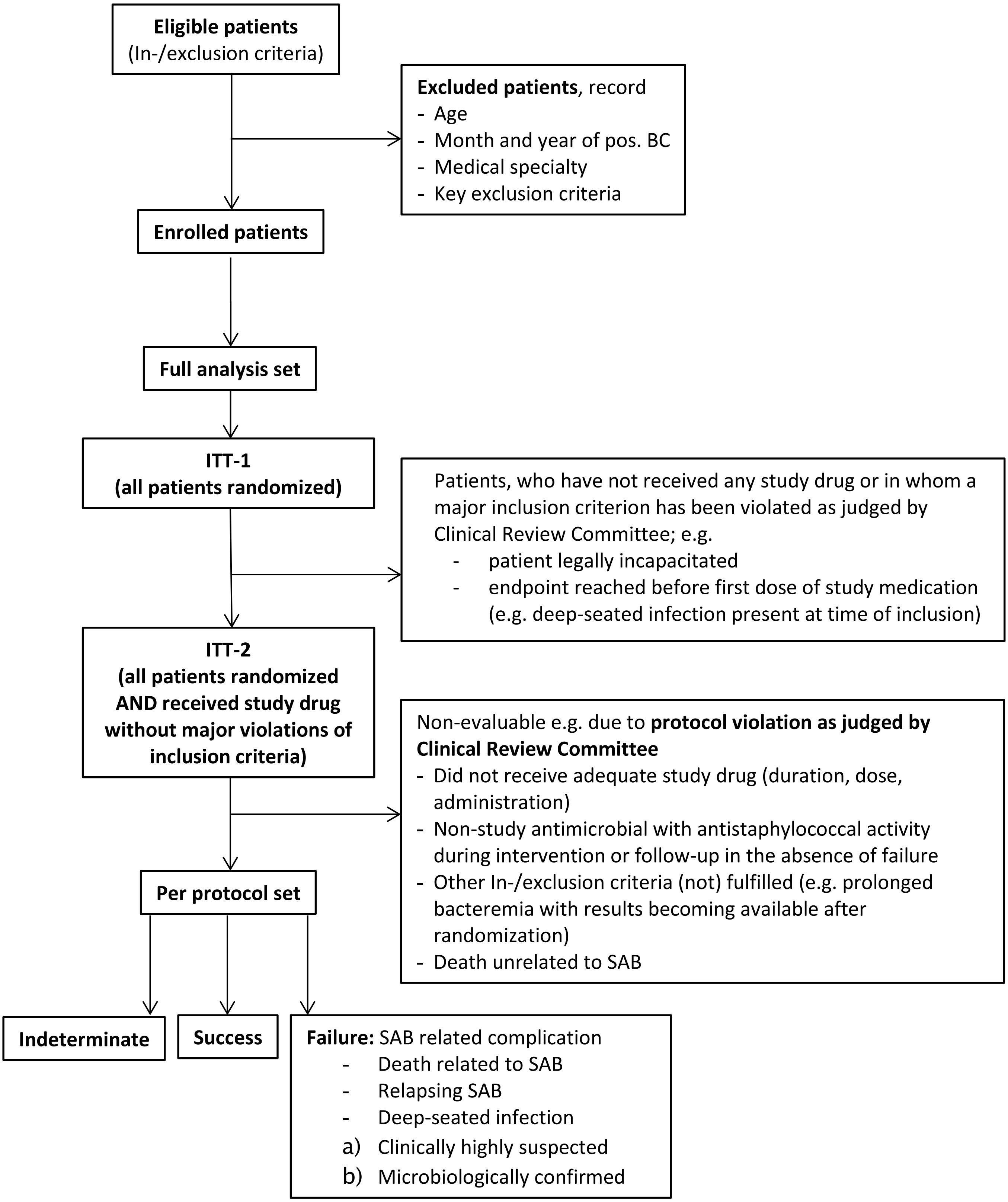
